# Testing Parametric Structure in Genetic Age-Effect Curves: A GAM-Based Framework with Application to the UK Biobank

**DOI:** 10.1101/2025.05.12.25327450

**Authors:** Yihe Yang, Xiaofeng Zhu

## Abstract

Before applying flexible nonparametric models such as a generalized additive model (GAM), it is natural to ask whether a simpler parametric form suffices. To address this question, we develop TAPS (Test for Arbitrary Parametric Structure), a framework that integrates estimation and hypothesis testing to evaluate whether a prespecified parametric form adequately captures a covariate effect in a GAM. TAPS accommodates diverse structures, including linearity, piecewise linearity with changepoints, and discontinuities with jumps, among others. It is implemented in the R package `mgcv.taps` built directly on `mgcv`, enabling seamless adoption, broad outcome support, and scalability to biobank-scale data. Using UK Biobank data, we analyze 38 continuous and 8 binary traits to investigate two scientific questions: does the effect of a polygenic risk score (PRS) vary with age beyond a linear interaction, and does retirement at age 65 modify this age-varying effect? We find that age-varying PRS effects are common and often strongly non-linear, and that retirement at 65 modifies these effects for 3 traits at a false discovery rate below 0.1.

## 1 Introduction

The generalized additive model (GAM) (Wood 2017), an extension of the generalized linear model (GLM) (Nelder & Wedderburn 1972), is widely used to model the non-linear effects of covariates on an outcome. Since its first introduction by Hastie & Tibshirani (1990), GAM has undergone substantial methodological and computational development. For instance, the R package `mgcv` exemplifies these advances, which provides a broad class of splines (Wood 2003) and multiple smoothing parameter selection methods (Wood 2011), and it has become the de facto standard software for GAM estimation and inference. Beyond methodological innovations, GAMs have also found wide applicability across scientific disciplines. For example, in epidemiology, GAMs have been used to smooth daily incidence data and to reveal trends in childhood type 1 diabetes during the COVID-19 pandemic (McKeigue et al. 2023). In ecology, GAMs have been employed to model the nonlinear effects of environmental factors in chaotic or near-chaotic ecological dynamic systems (Wood 2010). In spatial transcriptomics, GAMs are used to characterize the spatial distribution of cell types across tissues (Shang et al. 2025).

Before applying a nonparametric model such as a GAM, it is natural to ask whether a simpler parametric model can already describe the variation in the data. This is because a parametric model, if correctly specified, is often of greater scientific interest due to its interpretability and simplicity. For example, in genetics, the traditional assumption is that genetic effects of multiple variants across the genome are linear and additive (Hill et al. 2008), but growing evidence of non-linear genetic effects challenges this view (Zuk et al. 2012), sparking debate over whether linear additive models adequately capture the heritability of complex traits. In addition, in gene-environment (G×E) interaction analyses (Zhu et al. 2024, Winkler et al. 2024), it is usually assumed that a simple linear interaction model holds, meaning the genetic effect changes linearly with the environmental variable, an assumption that can be overly strong and potentially incorrect in practice.

Compared to the wide variety of estimation methods available for GAMs, the tools for hypothesis testing remain relatively limited. Most existing hypothesis test methods focus on assessing whether a smooth function is null (i.e., identically zero) (Wood 2013), rather than whether it conforms to a specific parametric structure of scientific interest. In this regard, only two main lines of work address parametric structures. For spline-based methods, hypothesis testing of specific parametric structures is largely restricted to evaluating whether a smooth term follows a polynomial form (Zhang & Lin 2003, Crainiceanu et al. 2005, Nummi et al. 2011). In the context of kernel-based methods, combined estimators have been developed to accommodate arbitrary parametric structures (Olkin & Spiegelman 1987, Fan & Ullah 1999, Mazo & Portier 2021), but extending them to GAMs with multiple smooth terms or non-Gaussian responses remains computationally challenging. To the best of our knowledge, no existing method enables hypothesis testing of arbitrary parametric structures within a GAM, especially in the presence of multiple smooth terms.

We develop the Test for Arbitrary Parametric Structure (TAPS) to assess whether a user-specified parametric structure adequately describes the relationship between a covariate and the response in a GAM. The methodological innovation of TAPS lies in constructing a mixed-effect representation in which the nonparametric component is forced to be orthogonal to any prespecified parametric form, thereby enabling rigorous assessment of whether the parametric structure adequately captures the target function within a GAM. Specifically, we develop an R package, `mgcv.taps`, which integrates seamlessly with `mgcv`, one of the most powerful GAM software packages. This integration enables TAPS to be easily adopted without additional learning costs, to support a wide range of outcome types including survival and ordinal responses (Wood et al. 2016), and to scale efficiently to biobank-scale datasets (Wood et al. 2017). We analyze UK Biobank data (Bycroft et al. 2018) to address two questions: (i) whether the effects of polygenic risk scores (PRSs) on complex traits vary with age beyond a simple linear interaction, and (ii) whether reaching retirement age at 65 coincides with a structural change in these age-varying effects. Applying TAPS to 38 continuous and 8 binary traits, we found that non-linear age-varying effects of PRSs were widespread, and that 3 traits exhibited significant trend changes at age 65 after multiple-testing correction.

## 2 Data and Scientific Questions

### 2.1 UK Biobank data

The UK Biobank is a large prospective cohort of approximately 500,000 individuals aged 40-70 years recruited across the United Kingdom between 2006 and 2010 (Sudlow et al. 2015). Participants completed extensive baseline assessments, including laboratory biomarker assays, standardized physical measurements, and touchscreen questionnaires on lifestyle, diet, and medical history, supplemented by interviews and linkage to electronic health records. Genotypes were phased with SHAPEIT3 and imputed using IMPUTE4 against a combined Haplotype Reference Consortium, UK10K, and 1000 Genomes Phase 3 reference panel, yielding approximately 96 million variants (Bycroft et al. 2018). With its scale, harmonized phenotyping, and dense genomic coverage, the UK Biobank is now a cornerstone resource for genetic epidemiology research.

### 2.2 Age-varying effect of PRS

The genetic additive model, first formalized by Fisher (1919), is the canonical framework for modeling genotype-phenotype relationships due to its interpretability, parsimony, and practical utility. It can be written as

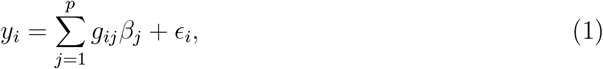

where *y*_*i*_ is the phenotype of individual *i, g*_*ij*_ is the genotype coding for variant *j, β*_*j*_ is its additive effect size, and *ϵ*_*i*_ is a residual error term. Polygenic effects refer to the collection of effect sizes *β*_1_, …, *β*_*p*_, typically with *p* large and each *β*_*j*_ small, so that the genetic contribution arises from the aggregation of many weak effects. The linear predictor 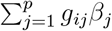 is then defined as a PRS, representing the linearly cumulative polygenic effects across variants (Zheng et al. 2024).

However, the adequacy of this genetic additive model has been debated. While many studies find that additive effects explain most genetic variance for complex traits, with non-additive effects contributing relatively little at the population level (Hill et al. 2008), other evidence points to systematic departures from additivity, including non-linear genotype-phenotype relationships, dominance, epistasis, and G×E interactions (Zuk et al. 2012). A common specification for G×E interaction is the following linear interaction model:

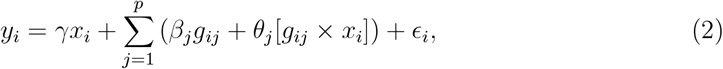

where *x*_*i*_ is an environmental variable, *γ* is the environmental effect, *β*_*j*_ represents the main effect of variant *j*, and *θ*_*j*_ represents its interaction effect with *x*_*i*_. Because individual variant effects are generally very small, single-variant interactions are very difficult to detect. An alternative approach (Aschard 2016) aggregates multiple variants into a PRS and tests for a universal interaction parameter *θ*_0_ in the model below:

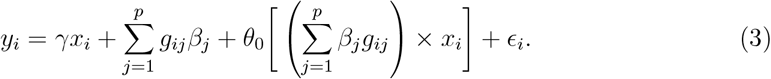

Nevertheless, genetic effects have been shown to exhibit context-specific heterogeneity, where their interactions with an individual’s environmental or physiological state can depart from linearity (Dahl et al. 2020). For example, Tian et al. (2025) demonstrated that alcohol intake exerts a causal effect on blood pressure only beyond a certain threshold. Hottenga et al. (2005) reported that the genetic effects of certain variants on systolic and diastolic blood pressure are stratified by age. Motivated by prior evidence of context-stratified genetic effects, our first scientific question is:

> *Among all individuals in the UK Biobank, does the effect of a PRS for a complex trait vary with age, and if so, is this age-varying effect linear or nonlinear?*

Mathematically, we consider the model

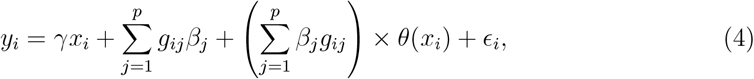

where *x*_*i*_ denotes age and *θ*(·) is a smooth function. In the literature, this model is known as the varying coefficient model (Hastie & Tibshirani 1993). Assessing whether *θ*(·) follows a simple linear form or a more complex pattern naturally leads to the hypothesis test

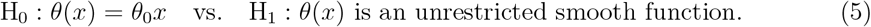

In practice, we will first test whether *θ*_0_ is significantly different from zero and then assess whether H_0_ is rejected or not, meaning that there is additional variation beyond the linear interaction. As will be introduced later, we develop a new hypothesis testing framework that allows the significance test of *θ*_0_ and the test in (5) to be carried out independently. To our knowledge, while previous work such as Jiang et al. (2021) has examined age-dependent changes by detecting single variants whose marginal genetic effects vary across age strata, our study is among the first to take a prediction-oriented perspective, asking whether the cumulative genetic effect captured by a PRS departs from a linear interaction with age.

### 2.3 Retirement-stratified age-varying effect of PRS

A growing number of studies show that retirement affects multiple aspects of late-life health. For example, exploiting the United Kingdom’s state pension age as a natural experiment, Furuya & Fletcher (2024) found that eligibility-induced retirement accelerated biological aging by about 0.87 years. In addition, Eibich (2015) reported that retirement improved self-rated health, reduced stress, and promoted healthier lifestyles in Germany. These studies suggest that retirement is a pivotal life-course event that can alter both biological and functional dimensions of aging.

Following our first question, we next investigate the following scientific question:

> *Among individuals near the retirement threshold in the UK Biobank, does the age-varying effect of a PRS exhibit a jump or a change-in-slope at age 65?*

Here, “near the retirement threshold” refers to UK Biobank participants aged 63 *<* age *<* 67, and “retirement” is defined as an indicator taking the value 1 if age ≥ 65 and 0 otherwise, based on the United Kingdom’s state pension age. Mathematically, we consider two commonly-used parametric structures in regression discontinuity design (RDD) and regression kink design (RKD) (Huntington-Klein 2021): a linear jump structure

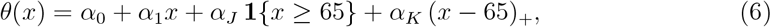

and a linear change-of-slope structure

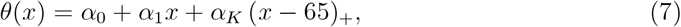

where *α*_0_, *α*_1_ describe the baseline linear trend, *α*_*J*_ is the jump parameter, and *α*_*K*_ is the change-of-slope parameter; **1**{*x* ≥ 65} is the indicator function (1 if *x* ≥ 65, 0 otherwise), and (*x* − 65)_+_ = max(*x* − 65, 0) is the truncated linear function. We test whether *θ*(*x*) can be sufficiently described by the structure (6) or (7) by using the following hypothesis tests:

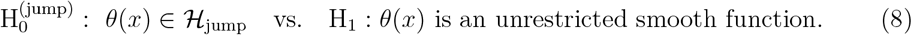

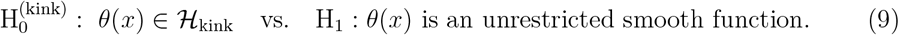

where ℋ_jump_ and ℋ_kink_ are the parametric spaces spanned by the basis functions (1, *x*, **1**{*x* ≥ 65}, (*x* − 65)_+_) and (1, *x*, (*x* − 65)_+_), respectively. We first assess whether the structural parameter *α*_*J*_ in (6) or *α*_*K*_ in (7) is statistically significant. If the parameter is significant and the null hypothesis 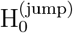 or 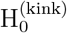 is not rejected, we infer that retirement is associated with a structural change at age 65. If the parameter is significant but 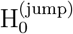 or 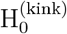 is rejected, the age-varying effect deviates from the simple jump or kink form, suggesting a more complex non-linear pattern. Conversely, if *α*_*J*_ or *α*_*K*_ is not significant, there is no evidence of a PRS-retirement interaction at age 65, regardless of whether 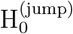 or 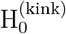 is rejected. As will be introduced later, our new hypothesis testing framework ensures that the test for the parametric components is independent of the tests in (8) and (9).

Note that this formulation differs from standard RDD and RKD, which directly test whether retirement alters a biomarker (e.g., a depression score). Here, our target is the structure of the age-varying PRS effects, and a detected structural change is not interpreted as the causal effect of retirement (Huntington-Klein 2021). We further restrict to individuals near the retirement threshold so that any detected change reflects local differences around the policy cutoff rather than long-term age trends (Huntington-Klein 2021).

## 3 Method

### 3.1 Overview of TAPS

To address the two scientific questions in Section 2, we need a framework that 1) flexibly captures non-linear relationships between the PRS and age and 2) formally tests whether this relationship conforms to prespecified parametric structures, such as linearity or structural breaks at the retirement threshold. The GAM provides a natural basis, enabling smooth, data-driven estimation of age-varying genetic effects while accommodating other covariates parametrically (Wood 2017). Building upon GAM, our proposed TAPS tests whether a smooth effect can be adequately represented by a given parametric structure.

Specifically, the model of a GAM is

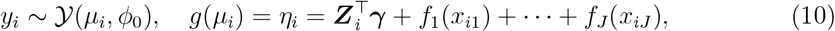

where *y*_*i*_ is the response belonging to a certain distribution Y(*µ*_*i*_, *ϕ*_0_) with mean *µ*_*i*_ and dispersion parameter *ϕ*_0_, *g*(·) is a known link function, *η*_*i*_ is a linear predictor, ***Z***_*i*_ is a vector of strict linear covariates, ***γ*** is a regression coefficient vector, and *f*_*j*_(·) is a smooth function of covariate *x*_*ij*_, *j* = 1, …, *J*. TAPS performs a hypothesis test on whether a target function in (10), such as *f*_1_(·), adheres to a parametric structure. The null and alternative hypotheses that TAPS addresses are

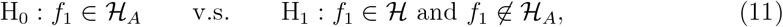

where ℋ_*A*_ is the space of functions with the target parametric structure and ℋ is a general functional space. In particular, ℋ should be sufficiently large to accommodate the parametric functions in ℋ_*A*_ (which may even be nonsmooth), but also be capable of approximating smooth functions when ℋ_*A*_ does not reflect the true structure.

To achieve this, we propose a new mixed-effects representation of *f*_1_(*x*):

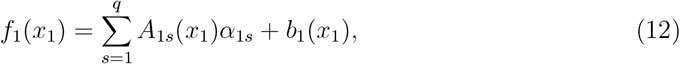

where ***A***_1_(*x*_1_) = (*A*_11_(*x*_1_), …, *A*_1*q*_(*x*_1_))^⊤^ is a vector of the basis functions of the parametric structure, ***α***_1_ = (*α*_11_, …, *α*_1*q*_)^⊤^ is a fixed vector, and *b*_1_(*x*_1_) is a non-parametric term orthogonal to 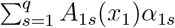. Here, we assume *b*_1_(*x*_1_) to lie in a reproducing kernel Hilbert space (RKHS) and be subject to this orthogonality constraint. According to the equivalence between a smooth function in an RKHS and a Gaussian process in *L*_2_ space (Kanagawa et al. 2018), *b*_1_(*x*) can be regarded as a Gaussian process with a covariance function

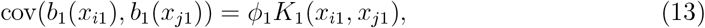

where *K*_1_(*x*_*i*_, *x*_*j*_) is a known bivariate function, and *ϕ*_1_ ≥ 0 is the variance of this Gaussian process. We call the equation (12) a mixed-effect representation of a function where 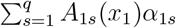 is the fixed effect and *b*_1_(*x*_1_) is the random effect, as it shares the same form as the generalized linear mixed model (GLMM) (Breslow & Clayton 1993). As a result, the hypothesis (11) reduces to

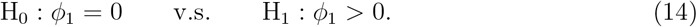

The techniques of GLMM, including the Wald test (Wood 2013) and score test (Zhang & Lin 2003), can yield the *p*-value of (14).

The orthogonality constraint is essential for identifiability: without it, the parametric component 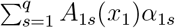 and the nonparametric component *b*_1_(*x*_1_) become confounded. Specifically, because typical spline bases (e.g., cubic splines, Gaussian correlation functions (Wood 2017)) can approximate most smooth functions with high accuracy, any parametric structure ***A***_1_(*x*_1_) that lies within the span of the spline basis can be absorbed into *b*_1_(*x*_1_), making the decomposition non-unique. This identifiability failure manifests as unstable parameter estimation and unreliable hypothesis tests in standard GAM implementations (**Supplementary Materials**). By enforcing orthogonality, TAPS ensures that 1) all parametric coefficients ***α***_1_ are uniquely identified regardless of the functional form of ***A***_1_(*x*_1_), and 2) the test of parametric significance (H_0_: ***α***_1_ = **0**) is statistically independent of the test of nonparametric deviation (H_0_: *ϕ*_1_ = 0), enabling separate assessment of whether the proposed structure is significant and whether it is sufficient.

TAPS can be simply extended to varying-coefficient models:

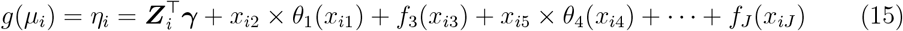

where the function *θ*_1_(*x*_1_) in an interaction term is the target of testing. Similar to the hypothesis test (11), TAPS can test

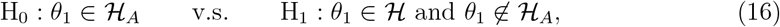

where ℋ_*A*_ is a parametric functional space and ℋ is a general functional space. Furthermore, we consider the same mixed-effects representation of *θ*_1_(*x*_1_) as (12):

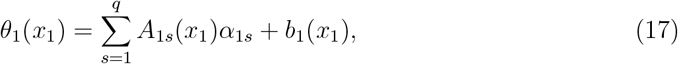

and we evaluate if *θ*_1_(*x*_1_) can be sufficiently described by the fixed effect 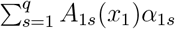 by testing whether the variance of the random effect *b*_1_(*x*_1_) is zero or not. In the context of the data and scientific questions outlined in Section 2, 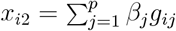 is the PRS term, and *x*_*i*1_ is age. The null space ℋ_*A*_ takes different forms depending on the question of interest: for the first scientific question, ℋ_*A*_ is the space of linear functions of age, whereas for the second question, ℋ_*A*_ is the parametric space with a jump or changepoint at *x* = 65.

Figure 1A shows a classic example used in the literature, the motorcycle acceleration data, which records head acceleration during a simulated crash (Wood et al. 2016). In this example, our goal is to test whether the time trend can be segmented into four phases, with changepoints marked by vertical dashed lines. The null hypothesis assumes that the function lies within a piecewise linear space ℋ_*A*_, whose basis functions are shown in Figure 1C. To test for potential deviations from this structure, TAPS constructed *b*_1_(*x*) from a space ℋ_*B*_ = ℋ*/* ℋ_*A*_, with basis functions adaptively learned from the data, as shown in Figure 1D. Figure 1B displays the fitted trend under the mixed representation. The first three phases aligned with the piecewise linear form, while the fourth showed a slightly non-linear effect. Figure 1E summarizes the overall TAPS workflow.

**Figure 1:**
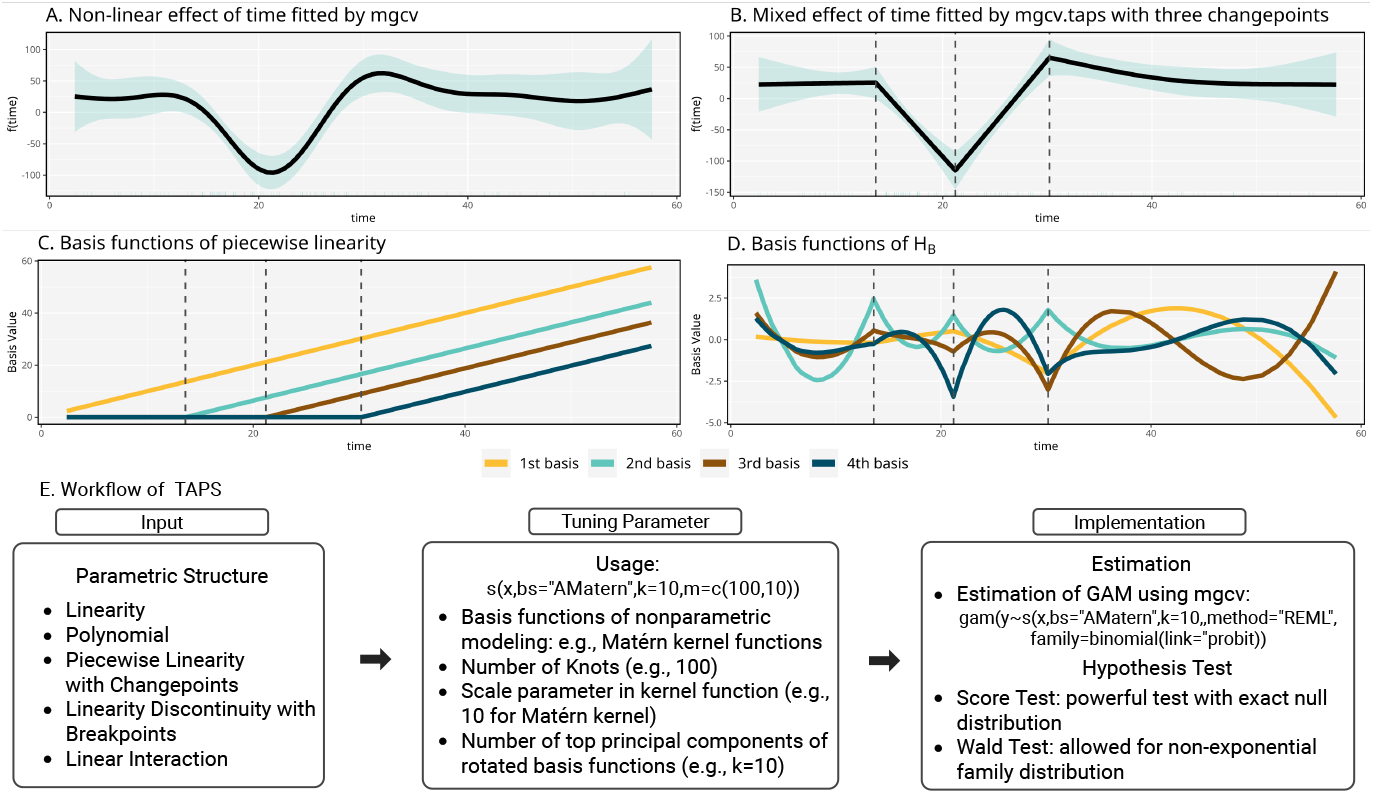
Overview of the TAPS framework. This figure uses the `mcycle` data, which records head acceleration over time in a simulated motorcycle crash experiment, to illustrate the workflow of TAPS. These data are provided by the R package `MASS` and are commonly used for benchmarking smooth curve fitting methods. **A**. The fitted smooth function of time using `mgcv` with default smooth terms. **B**. The fitted smooth function of time using `mgcv.taps` with three user-specified changepoints. **C**. Basis functions of the piecewise linear space ℋ_*A*_. **D**. Basis functions from the kernel-smoothed space ℋ_*B*_, automatically learned via principal component decomposition. **E**. Summary of the TAPS components.

### 3.2 Methodology of TAPS

Methodologically, we address two key challenges in the implementation of TAPS. First, we show how to construct the random effect *b*_1_(*x*) that is adaptively orthogonal to an arbitrary prespecified ***A***_1_(*x*), guaranteeing an identifiable mixed-effect representation. Second, we explore the methodology to conduct the hypothesis test (14) within a GAM model. We design two testing procedures: a Wald test (Wood 2013) and a score test (Zhang & Lin 2003). The score test is generally more powerful, yet currently limited to outcomes from the exponential family, whereas the Wald test supports all outcome types implemented in `mgcv`. By offering both options, users can choose the method best suited to their analysis objectives. Additional technical details are provided in the **Supplementary Materials**.

#### 3.2.1 Construction of basis functions

The construction of *b*_1_(*x*) starts from representing it using the RKHS (Wahba 1990, Wainwright 2019), under the alternative hypothesis that *b*_1_(*x*) is a general smooth function. According to the functional analysis theory, every RKHS H_*K*_ corresponds to a unique semi-positive definite kernel function *K*(·, ·) defined in the Cartesian product space [0, 1] × [0, 1]. For any function *b*_1_ ∈ ℋ_*K*_,

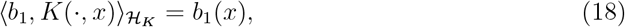

where 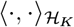 is the inner product in the space ℋ_*K*_. This property ensures that for *n* observations {*x*_1_, …, *x*_*n*_} ⊂ [0, 1], there must exist an (*n* × *n*) matrix **K**_1_ with the (*i, j*)th element being *K*(*x*_1*i*_, *x*_1*j*_) and an (*n* × 1) vector ***θ***_1_ = (*θ*_11_, …, *θ*_1*n*_)^⊤^ such that

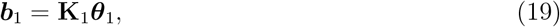

where ***b*** = (*b*(*x*_1_), …, *b*(*x*_*n*_))^⊤^. The Matérn correlation function (Ruppert et al. 2003)

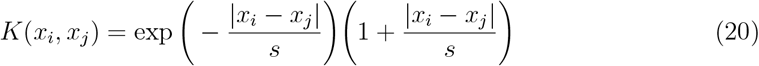

is used as the default to construct the design matrix of the random effect with the scale parameter *s* = 10. Other kernel functions have also been investigated, with results showing that TAPS is indeed not sensitive to the choice of kernel functions (Figure S2-S5).

We construct the basis functions of ℋ_*B*_ = ℋ_*K*_*/*ℋ_*A*_ by finding an ((*n* + *q*) × *n*) orthogonal matrix **Q** satisfying the following linear constraint

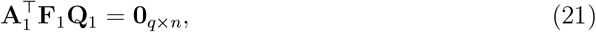

where **F**_1_ = (**A**_1_, **K**_1_) is the combination of **A**_1_ and **K**_1_. Here, the orthogonal matrix **Q**_1_ describes a special linear transformation that rotates the columns of **F**_1_, i.e., the basis functions of ℋ, into a new one perpendicular to **A**_1_. As a result, the columns of **F**_1_**Q**_1_ can be regarded as the basis functions of ℋ_*B*_ and thus it can represent ***b***_1_ = **F**_1_**Q**_1_***π***_1_ where ***π***_1_ is an unconstrained vector in ℝ^*n*^. The QR decomposition can yield a candidate for the orthogonal matrix **Q**_1_, where the details are shown in **Supplementary Materials**.

Mathematically, there is a well-known equivalence between a smooth function in an RKHS induced by a kernel function *K*(·, ·), and a Gaussian process defined on an *L*_2_ space with a covariance function the same as the kernel function *K*(·, ·) (Kanagawa et al. 2018). That is, for a smooth function *b*_1_(*x*) ∈ ℋ_*K*_, its *n* realizations mathematically can be regarded as

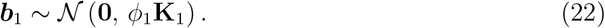

However, *b*_1_(*x*) is not a free function in ℋ_*K*_: it should be orthogonal to the fixed effect 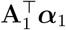, and hence there is an additional step to construct its prior distribution. Specifically, as ***b***_1_ falls within ℋ = ℋ_*A*_ ∪ ℋ_*K*_ and **F**_1_ is a basis matrix of ℋ, ***b***_1_ can also be represented as

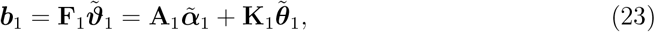

where the vector 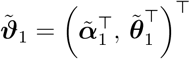. The constraint to ensure ***b***_1_ fully lies within ℋ_*B*_ is

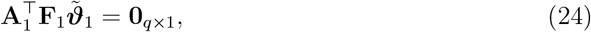

because 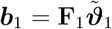 must be orthogonal to **A**_1_***α***_1_. According to Wood (2003, Page 99), any 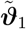 subject to the constraint (24) can be represented as 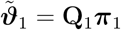, where **Q**_1_ is the basis matrix for 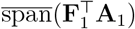 and ***π***_1_ is a free (*n* × 1) vector. As a result, 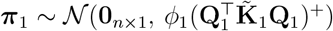 where 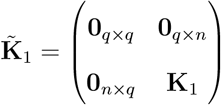 . Subsequently, since ***b***_1_ = **F**_1_**Q**_1_***π***_1_, the prior distribution of ***b***_1_ is

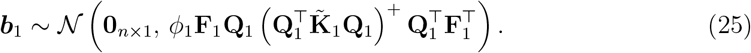

Thus, testing whether *f*_1_ = **A**_1_***α***_1_ reduces to testing whether *ϕ*_1_ = 0.

#### 3.2.2 Dimension reduction

Constructing **B**_1_ requires the QR decomposition of the ((*n* + *q*) × *q*) matrix 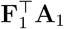, which is costly for large *n*. To alleviate this, we use two dimension-reduction strategies. First, we construct the smooth function with a low-rank RKHS representation (Kammann & Wand 2003), where the number of knots *p* = max(*n/*4, 100) is used by default. Second, we apply principal component analysis (PCA) (Wood 2003) to further reduce the dimension of the basis matrix **F**_1_**Q**_1_ and the corresponding penalty matrix 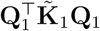. Although the penalty matrix 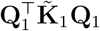 may be indefinite, the leading eigenvalues retained by PCA are typically far from zero, so a non-positive definite covariance matrix does not occur in practice.

#### 3.2.3 Score test

We extend the score test proposed by Zhang & Lin (2003) to evaluate whether the smooth deviation term *b*_1_(*x*) is equal to zero. This test is based on the equivalence between GAM and GLMM. Specifically, the score test is constructed from the following GLMM model:

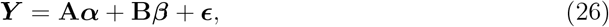

where ***Y*** = (*Y*_1_, …, *Y*_*n*_)^⊤^ with *Y*_*i*_ = *η*_*i*_ + (*y*_*i*_ − *µ*_*i*_)*g*^′^(*µ*_*i*_), **A** is the design matrix for fixed effects ***α***, and **B** is the design matrix for random effects ***β***. The noise term follows a Gaussian distribution ***ϵ*** ∼ N(**0**, *ϕ*_0_**W**^−1^), where *ϕ*_0_ is the dispersion parameter, **W** = diag(*W*_1_, …, *W*_*n*_) and *W*_*i*_ = 1*/*(*V* (*µ*_*i*_)*g*^′^(*µ*_*i*_)^2^), with *V* (*µ*) denoting the variance function and *g* the canonical link function. For the GAM (10), **A** = (**A**_1_, …, **A**_*J*_, **Z**), **B** = (**F**_1_**Q**_1_, …, **F**_*J*_ **Q**_*J*_), 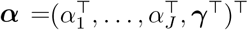, and 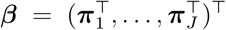. Since both **B*β*** and ***ϵ*** are random, it is reasonable to consider them as a new noise **e** = **B*β*** + ***ϵ***, whose prior distribution is N (**0**_*n*×1_, **V**(***ϕ***)) with ***ϕ*** = (*ϕ*_1_, …, *ϕ*_*J*_)^⊤^ and

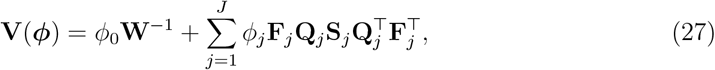

where **S**_*j*_ is the penalty matrix for the *j*th smooth term (Wood 2017). For instance, 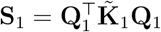 for the target smooth term *f*_1_.

We first apply the R package `mgcv` to estimate the model parameters. Specifically, the regression coefficients ***α*** and ***β*** are estimated by solving:

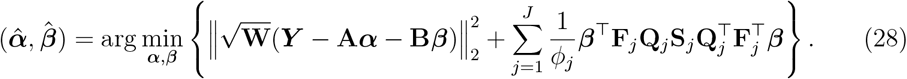

Given the current estimate 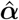, the variance components ***ϕ*** = (*ϕ*_1_, …, *ϕ*_*J*_)^⊤^ are then estimated by minimizing the restricted maximum likelihood (REML) function:

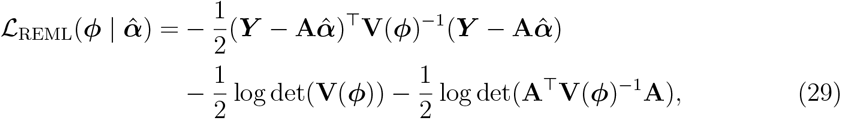

where 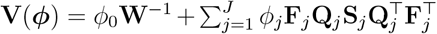. The R package `mgcv` proceeds by iteratively updating 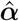 (and 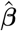) given the current 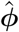, and then updating 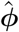 based on the updated 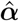 (and 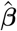), until convergence (Wood 2011).

Based on the outputs of `mgcv`, we then perform the score test. Specifically, the score equation **U**(·) of the REML function (29) is:

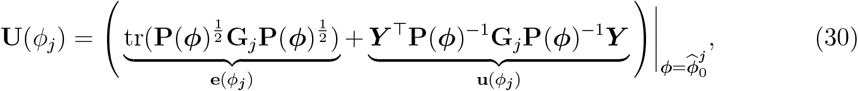

where 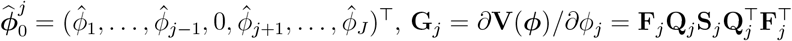, and

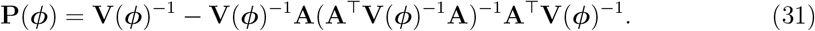

Wu et al. (2011) showed that under the null hypothesis (i.e., *ϕ*_1_ = 0), **u**(*ϕ*_1_) in (30) follows:

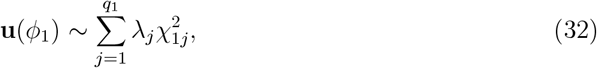

where *q*_1_ is the rank of the matrix **S**_1_, 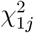 denotes a chi-squared random variable with 1 degree of freedom, and *λ*_1_, …, *λ*_*q*1_ are the eigenvalues of the matrix 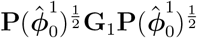 . Following sequence kernel association test (SKAT) and the related methods (Wu et al. 2011), we use numerical approximations to calculate the *p*-value under the null distribution (32). We find that a hybrid strategy outperforms single approximations: we prioritize the exact numerical integration and automatically switch to the Liu method for extreme tail probabilities (Liu et al. 2009), both implemented by the R package CompQuadForm.

When the sample size *n* is large, explicitly constructing the (*n* × *n*) precision matrix **V**(***ϕ***)^−1^ and projection matrix **P**(***ϕ***) becomes computationally infeasible. We therefore develop an operator-based strategy that avoids materializing these matrices. Specifically, we define matrix-vector product operators that compute **V**(***ϕ***)^−1^***v*** and **P**(***ϕ***)***v*** for any vector ***v*** without forming the full (*n* × *n*) matrices. For **V**(***ϕ***)^−1^, we apply the Woodbury matrix identity to express the product as a combination of diagonal operations and low-rank updates. For **P**(***ϕ***), we construct the projection operator using **V**(***ϕ***)^−1^ without explicit matrix inversion. With these improvements, the score test runs in about 10 seconds on 10 CPU cores with 100 GB memory for about 350,000 individuals.

#### 3.2.4 Wald test

We adopt the Wald test proposed by Wood (2013) to evaluate whether the smooth deviation term *b*_1_(*x*) is equal to zero. Specifically, let 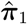 denote the estimate of ***π***_1_ under the estimated variance component 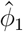. The null hypothesis is H_0_: *b*_1_(*x*) = 0. The Wald test statistic is given by 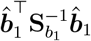, where 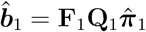 and 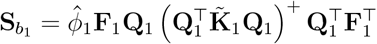. Although this statistic is theoretically assumed to follow a *χ*^2^ distribution, Wood (2013) showed that it is a mixture of *χ*^2^ distributions due to the penalization imposed on *b*_1_(*x*). The Wald test can be performed directly using the function `testStat(·) in mgcv`.

#### 3.2.5 Simulation

Owing to space constraints, the technical details of TAPS and its evaluation using simulations are presented in the **Supplementary Materials**. Specifically, we conducted extensive simulations to evaluate the Type I error control and power of both the score test and Wald test under various parametric structures and outcome distributions. Section 2 of the **Supplementary Materials** presents the complete simulation design, including data generation procedures (Section 2.1), four structural hypotheses: linearity, piecewise linearity, linearity with discontinuity, and linear interaction (Sections 2.2–2.5), and implementations for Gaussian, binary, Poisson, survival, ordinal outcomes, and time-varying model (Sections 2.6–2.11), with illustrative R codes and mathematical formulations.

Section 3 of the **Supplementary Materials** summarizes the key findings. Under the null hypothesis, the score test exhibited proper Type I error control with p-values closely following the uniform distribution, while the Wald test was slightly conservative (Figures S6A–B, S7A-B, S10A, S11–S13). Both tests demonstrated high power to detect deviations from the hypothesized structures, with power exceeding 0.9 for moderate sample sizes when deviations were substantial (Figures S6C–D, S7C-D, S8-S9, S10B). The methods performed consistently across logistic and probit links (Figure S7), remained robust to outliers when using median GAM (Figure S8), and extended successfully to survival and ordinal outcomes (Figure S9) as well as varying-coefficient models (Figure S10).

## 4 Real Data Analysis

### 4.1 Linear interaction test of age-varying PRS effect

We applied TAPS to UK Biobank data to investigate whether the age-varying effect of a PRS can be adequately represented by a linear structure for 38 continuous traits and 8 binary traits, which corresponds to the traditional G×E interaction analysis (Aschard 2016). For non-disease traits (including age, smoking initiation, frequent drinking, and medication status), we used baseline measurements. Regarding the cardiovascular diseases, cases were defined as individuals diagnosed with the condition at any time during follow-up, and controls were additionally restricted to individuals free of any cardiovascular condition (**Supplementary Table 1**). Medication status (Data Field: 6153, 6177) was adjusted for systolic blood pressure (SBP), diastolic blood pressure (DBP), apolipoprotein B (APOB), low-density lipoprotein cholesterol (LDL), and total cholesterol (TC), following standard adjustment procedures established in the literature (Graham et al. 2021, Keaton et al. 2024). Hypertension was defined as SBP ≥ 140 or DBP ≥ 90 or taking blood-pressure-lowering medication using the baseline measurements. In addition, our analysis was restricted to the intersection of participants of genetically-inferred (Privé et al. 2022) and self-reported (Data Field: 21000) European ancestry in the UK Biobank. Finally, we removed the related individuals (kinship *>* 0.0884) and excluded withdrawn participants.

**Table 1:** Test of piecewise linear age-varying effect of a PRS.

| Trait | Abbreviation | Parametric Part |  |  | Nonparametric Part |  |
| --- | --- | --- | --- | --- | --- | --- |
| | | Location | Estimate | $p$-value | Wald | Score |
| Systolic Blood Pressure | SBP | Age=57 | -0.0292 | 3.854E-50 | 0.0922 | 0.0599 |
| Diastolic Blood Pressure | DBP | Age=52 | -0.0177 | 1.820E-11 | 0.9755 | 0.9479 |
| Pulse Pressure | PP | Age=61.8 | -0.0345 | 1.57E-37 | 0.1666 | 0.0506 |

Estimate = fixed-effect estimate of change-of-slope ( $\alpha_K$ ); $p$ -value = nominal significance of the fixed effect; Wald and Score = random-effect tests for non-linearity after accounting for the fixed effect.

Many traits exhibited skewed distributions and had some outliers; therefore, we applied inverse-rank normalization (IRNT) to these traits and used the corresponding GWAS summary statistics from the Neale Lab (Lab 2018) for each setting. PRSs were estimated using SBayesRC (Zheng et al. 2024) with the default settings and predicted for individuals of European ancestry in the UK Biobank using PLINK (Purcell et al. 2007). In addition, the GAM we used is

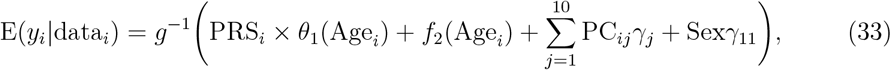

where *y*_*i*_ is a trait, *g*(·) is the link function corresponding to the distribution of *y*_*i*_, PRS_*i*_ is its PRS, PC_*ij*_ is the *j*th genetic PC, and data_*i*_ denotes all the covariates. We aimed to test whether H_0_ : *θ*_1_(Age_*i*_) = *θ*_0_Age_*i*_ for a certain parameter *θ*_0_, as described in (5).

As shown in Figure 2A, the top panel presents the Manhattan plot of − log_10_(*p*) values from the t-tests for linear age-PRS interactions, while the bottom panel reports − log_10_(*p*) values from the score tests evaluating non-linear age-varying effects orthogonal to the linear interaction term. Full results are shown in **Supplementary Table 2**. Among the 46 traits analyzed, 36 had *p*-values below the Bonferroni-corrected threshold for the linear interaction term, and 16 had *p*-values below the same threshold for the non-linear term. Notably, the most significant score test *p*-value for non-linear age-varying effects was observed for SBP (*p*-value = 8.34E-53), followed by pulse pressure (PP) (*p*-value = 4.68E-35) and gamma-glutamyl transferase (GGT) (*p*-value = 2.41E-15). Figure 2B shows computational times for both GAM fitting (using `mgcv`) and our score test, with both approaches completing within 10 seconds on standard high-performance computing (HPC) infrastructure.

**Table 2:** Results of jump and change-of-slope tests of age-varying effects of PRSs at age 65.

| Trait | Abbreviation | Parametric Part |  |  | Nonparametric Part |  |
| --- | --- | --- | --- | --- | --- | --- |
| Test of jump at age = 65 |  |  |  |  |  |  |
|  |  | Estimate | *p*-value | *q*-value | Wald | Score |
| Apolipoprotein A | APOA | -0.0295 | 0.0295 | 0.339 | 0.9318 | 0.4259 |
| Cystatin C | CysC | 0.0350 | 0.0105 | 0.241 | 0.9869 | 0.7955 |
| LDL Cholesterol | LDL | -0.0485 | 0.0274 | 0.420 | 0.4861 | 0.1887 |
| Total Cholesterol | TC | -0.0474 | 0.0036 | 0.1669 | 0.1286 | 0.0631 |
| Test of change-of-slope at age = 65 |  |  |  |  |  |  |
|  |  | Estimate | *p*-value | *q*-value | Wald | Score |
| Albumin | ALB | 0.0307 | 0.0091 | 0.1041 | 0.0706 | 0.0084 |
| Monocyte Count | MONO | -0.0634 | 0.0062 | **0.0950** | 0.4726 | 0.2283 |
| Total Proteins | TP | -0.1084 | 0.0014 | **0.0620** | 0.4716 | 0.2283 |
| Urate | UA | -0.0591 | 0.0113 | 0.1043 | 0.9885 | 0.8821 |
| Leukocyte Count | WBC | -0.0806 | 0.0030 | **0.0699** | 0.4205 | 0.2171 |

Estimate = fixed-effect estimate of jump ( $\alpha_J$ ) or change-of-slope ( $\alpha_K$ ); *p*-value = nominal significance of the fixed effect; *q*-value = FDR-adjusted *p*-value; Wald and Score = random-effect tests for non-linearity after accounting for the fixed effect. Bold *q*-values indicate significance at FDR < 0.1. The FDR *q*-value of the score test for ALB is 0.1943.

**Figure 2:**
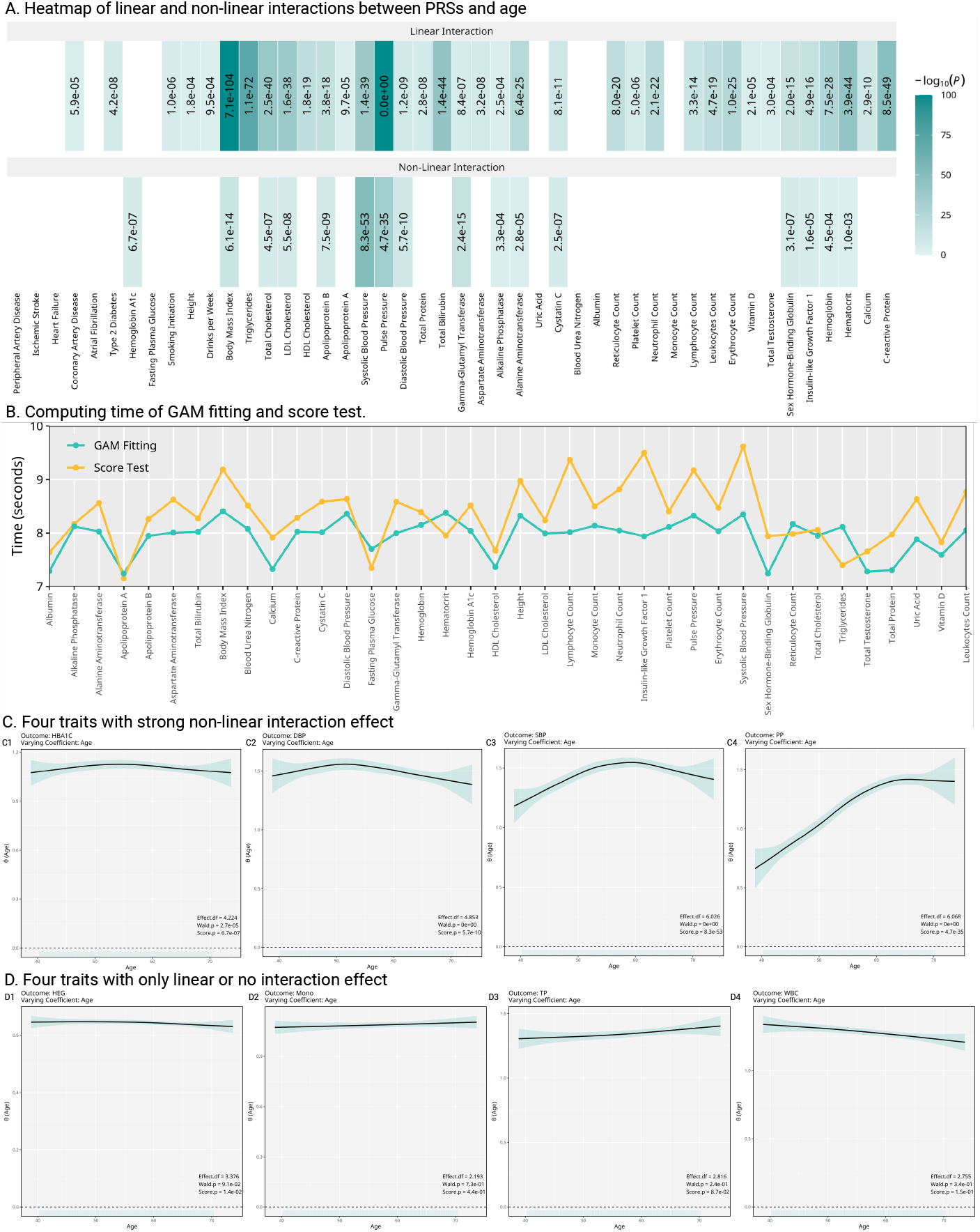
Age-varying coefficient of PRS effects. **A**. Heatmaps of interaction tests evaluating whether PRS effects vary with age. The top panel shows −log_10_(*p*)-values from t tests for linear age-PRS interactions, while the bottom panel shows −log_10_(*p*)-values from score tests for non-linear age-varying effects, orthogonal to the linear interaction terms. **B**. Line plot of computing times for GAM fitting and score testing. **C**. Estimated age-varying effect functions for selected traits with strong evidence of nonlinearity. **D**. Estimated age-varying effect functions for traits with linear or no detectable interaction. Each curve refers to the fitted age-dependent PRS effect; shaded bands indicate 95% confidence intervals. Each panel reports the edf, the score test and the Wald test *p*-values.

Figure 2C shows estimated age-dependent PRS effect curves for four representative traits with significant non-linear interaction effects: hemoglobin A1C (HBA1C), DBP, PP, and SBP. The linearity parameter for HBA1C was not significant in Figure 2A because its age-varying PRS effect exhibited a symmetric U-shaped pattern. In addition, a consistent pattern emerged across these examples: the age-varying effect appeared approximately piecewise linear, with inflection points typically occurring between ages 50 and 60. In particular, both DBP and SBP exhibited a rise-and-fall structure: the PRS effect increased during middle age and then declined later in life, with the changepoint for DBP occurring earlier than those for PP and SBP.

Figure 2D presents traits with either purely linear or no significant age-varying genetic effects. For instance, height (HEG), monocytes (MONO), total proteins (TP), and leukocytes (WBC) showed significant linear interaction effects without evidence of non-linearity. These traits served as negative controls, demonstrating that our score test was not overly liberal: it successfully distinguished between linear and non-linear patterns and yielded appropriately conservative results when no age-dependent variation was present. Note that the age-decreasing effect of the HEG PRS may be due to a slight reduction in height as age increases, thereby diminishing the effectiveness of the HEG PRS in older individuals.

#### 4.1.1 Follow-up analysis I: split-sample PRS construction

Neale Lab GWAS summary statistics are derived from UK Biobank participants of European ancestry. Using these summary statistics to construct PRSs and evaluate them in the same cohort may introduce training-testing overlap, which can bias PRS-phenotype associations (Mak et al. 2018). In this follow-up analysis, we selected 34 continuous traits that had been previously processed and quality controlled in our laboratory, which substantially overlapped with, but were not identical to, the traits analyzed in the main study. The full sample was randomly split into two independent groups (Group A and Group B), stratified by age bins and sex. For each group, a GWAS was performed using IRNT traits as outcomes, adjusting for age, age squared, sex, the first 15 genetic PCs, and leave-one-chromosome-out PRSs, implemented in PLINK (Purcell et al. 2007). To ensure no sample overlap between PRS construction and downstream analysis, PRSs derived from one group were applied to the other group in a cross-validation manner, though this may result in some loss of power. Full results can be found in **Supplementary Table 3**.

We then fitted the same TAPS models (33), using PRSs trained from the non-overlapping sample. Figure 3 summarizes the results. Applying the same *p*-value threshold as in the main analysis (0.05/46), we identified 14 significant fixed-effect signals in each group, along with 5 and 8 significant random-effect signals in Groups A and B, respectively. The overall reduction in detected signals compared with the full-sample analysis is expected, given the approximately twofold decrease in effective sample size. Importantly, the split-sample results are qualitatively consistent with our main findings, supporting the conclusion that many PRS effects vary with age and that a non-negligible proportion of these age-varying effects cannot be adequately captured by a simple linear PRS-by-age interaction.

**Figure 3:**
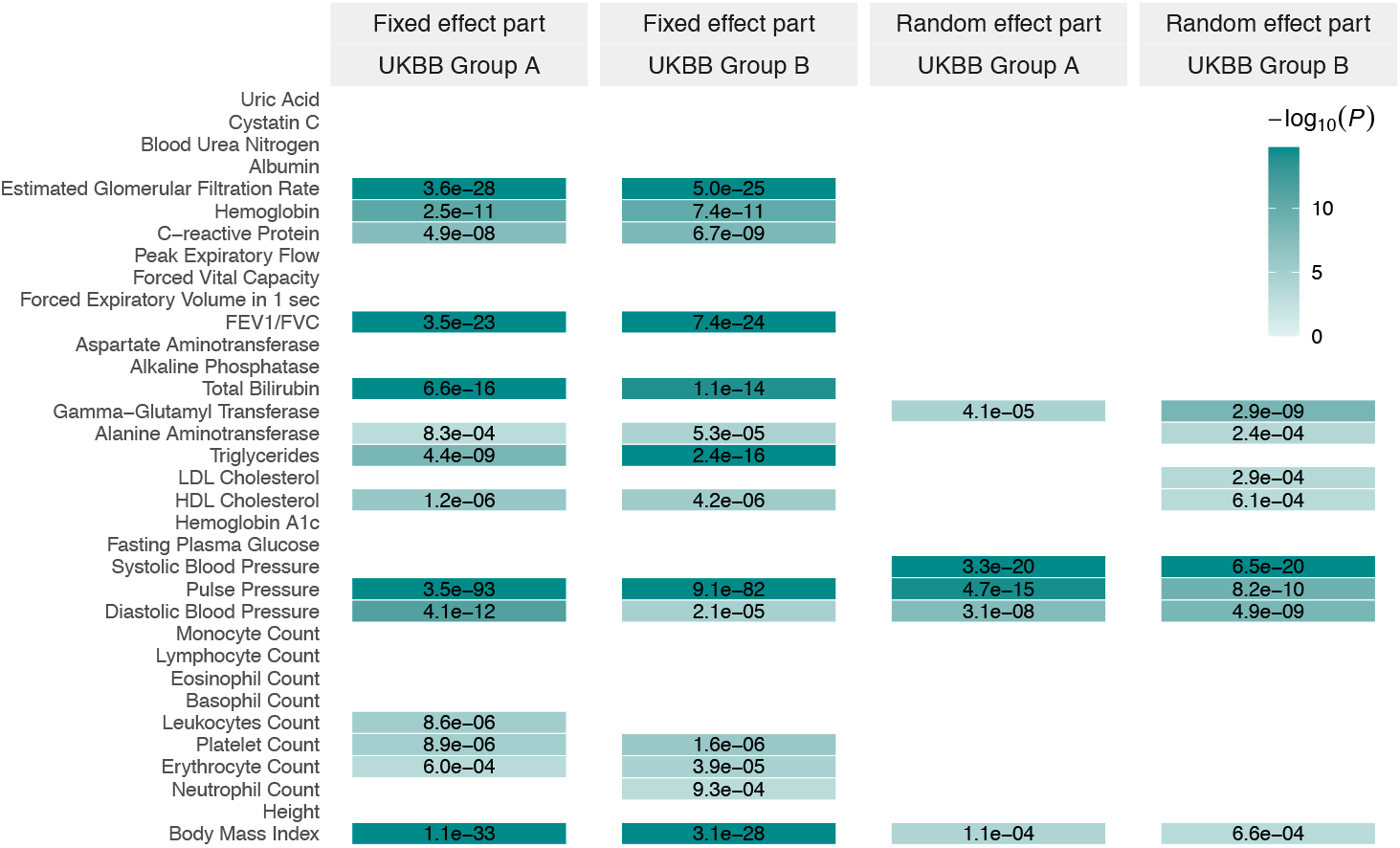
Heatmaps of −log_10_(*p*) values from split-sample analyses in UK Biobank, shown separately for fixed-effect and random-effect components in Group A and Group B. Each row corresponds to a trait, and darker colors indicate stronger statistical evidence. Only associations that remain significant after adjustment for multiple comparisons are displayed.

#### 4.1.2 Follow-up analysis II: piecewise linear vs nonlinear PRS effect

As a follow-up to the main analysis, we further examined whether the significant non-linear age-varying effects could be parsimoniously represented by a piecewise linear function with empirically determined changepoints. Specifically, we observed that many age-varying effects of PRSs exhibited an approximately piecewise linear form with one or two changepoints of slope. For example, in Figures 2C2-2C4, the three blood pressure traits seemed to follow a piecewise linear form with a single changepoint. Motivated by this pattern, we selected SBP, DBP, and PP as examples and tested whether their age-varying effects could be adequately described by a piecewise linear structure with one changepoint, located at age = 57 for SBP, age = 52 for DBP, and age = 61.8 for PP. These changepoints were selected manually, rather than through a data-driven method. The null hypothesis we tested was:

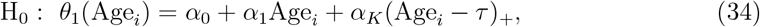

where *τ* = 57, 52, and 61.8 for SBP, DBP, and PP, respectively.

Table 1 summarizes the structure tests for a single-changepoint piecewise linear specification of the age-varying effects of SBP, DBP, and PP PRSs. For all three traits, the fixed-effect estimates of the change-of-slope (*α*_*K*_) were negative and highly significant, indicating a consistent attenuation of the genetic effect after the changepoint. In contrast, both the Wald and score tests for the variance of the random effect were non-significant (*p*-value*>*0.05) for all traits, suggesting no residual non-linearity after accounting for the fixed change-of-slope. Figure 4A provides a visual illustration of these patterns: the PRS effects increase with age up to the estimated changepoints, age 57 for SBP, age 52 for DBP, and age 61.8 for PP, and subsequently decline, with no evidence of additional structural complexity beyond the specified piecewise linear structures. Figure 4B shows that the non-overlapping split-sample curves for SBP, DBP, and PP closely match those from the full-sample analysis, supporting the robustness of these age-varying patterns.

**Figure 4:**
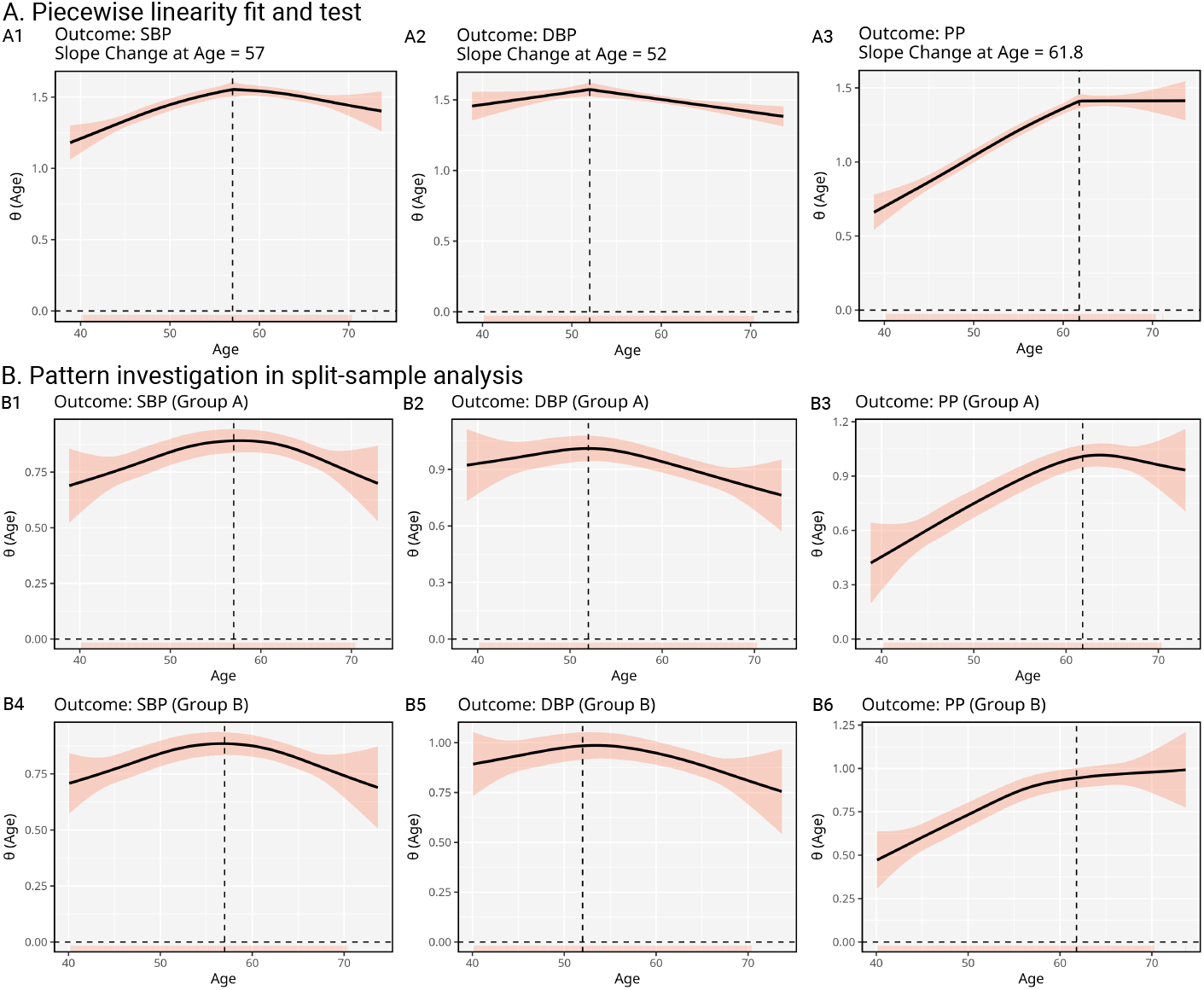
Change-of-slope analyses of age-varying PRS effects for blood pressure traits. **A**. Piecewise linear fits for SBP (A1), DBP (A2), and PP (A3) in the full sample, with estimated slope change points at ages 57, 52, and 61.8, respectively. Each curve shows the fitted age-dependent PRS effect with 95% confidence intervals (shaded bands). **B**. Split-sample validation showing consistent age-varying patterns across independent Group A (B1-B3) and Group B (B4-B6) for the same three traits.

### 4.2 Structural change tests of retirement-stratified age-varying PRS effect

We applied TAPS to UK Biobank data to statistically assess whether the age-varying effect of a PRS undergoes a structural change at the statutory retirement age of 65 years, considering both a jump and a change-of-slope specification, i.e., the hypotheses (8) and (9) described in Section 2.3. The analysis focused on participants aged 63-67 from the genetically inferred European ancestry subset of the UK Biobank. We again examined the 38 continuous traits and 8 binary traits. Continuous traits were IRNT-transformed prior to analysis, and the corresponding GWAS summary statistics from the Neale Lab were used. PRSs were estimated using SBayesRC and predicted for individuals. Moreover, we considered the same GAM model as (33). Details are provided in **Supplementary Tables 4-5**.

Table 2 shows the results of the jump and change-of-slope tests for age-varying effects of PRSs at the statutory retirement age of 65. Overall, most of the 46 traits analyzed in the retirement-stratified setting showed no evidence of a structural change in the age-varying effects of PRSs at age = 65. For traits listed in Table 2, although one score test reached nominal significance, none remained significant after false discovery rate adjustment, suggesting limited evidence for additional residual nonlinearity beyond the specified jump or change-of-slope term. In the jump-at-65 analysis, four traits (APOA, CysC, LDL, and TC) exhibited nominal significance for the jump parameter *α*_*J*_ (*p <* 0.05), but none remained significant after controlling the false discovery rate at 0.1. As illustrated in Figure 5A-5C, CysC shows an upward shift at age 65, whereas LDL and TC display downward shifts, while the remaining traits exhibit smooth age-varying patterns without clear discontinuities. In the change-of-slope analysis, five traits showed nominal evidence of a slope change at age 65; among them, MONO, TP, and WBC exhibited borderline significance after FDR adjustment (*q*-value *<* 0.1). As shown in Figure 5D-5F, these effects correspond to subtle changes in trend, with gradual increases before age 65 followed by mild attenuation thereafter.

**Figure 5:**
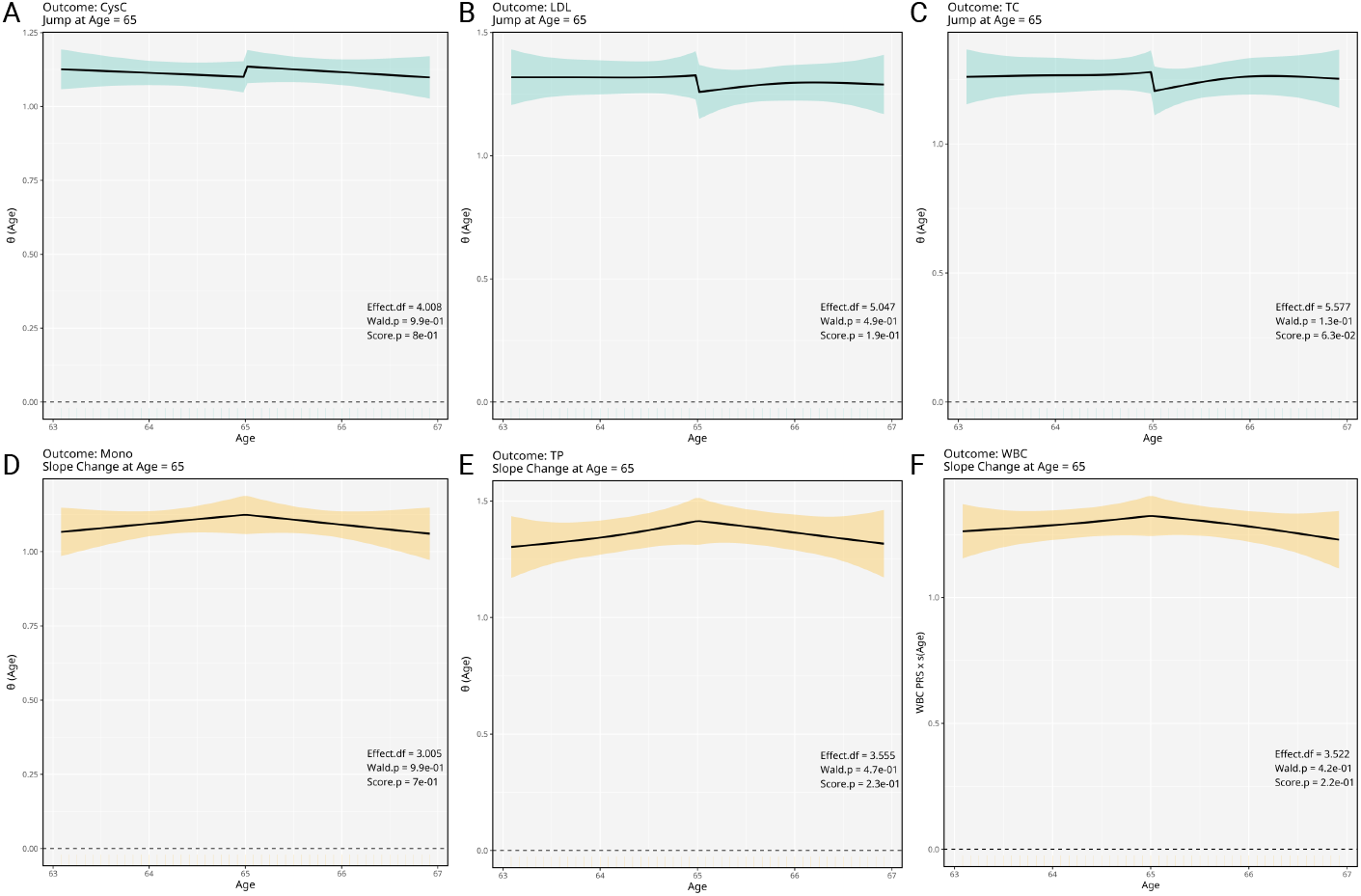
Jump- and change-of-slope analyses of age-varying effects of PRSs at the statutory retirement age of 65 years. **A**-**C**: Representative traits under the jump-at-65 model. **D**-**F**: Representative traits under the slope-change-at-65 model. Each curve represents the fitted age-dependent PRS effect; shaded bands indicate 95% confidence intervals. Each panel reports the edf, the score test *p*-value and the Wald test *p*-value.

## 5 Discussion

In this paper, we develop TAPS, a computational framework for testing whether a target function in a GAM can be fully described by a prespecified parametric structure. Existing tests are typically restricted to assessing whether a smooth term is zero, rather than whether it follows a scientifically meaningful parametric form. TAPS addresses this gap by formulating the null as membership in a given parametric structure and the alternative as a general smooth function. This formulation directly answers the question: Does the function follow the specified structure? If so, there is no need to apply a more complex and less interpretable nonparametric model.

Methodologically, TAPS ensures identifiability by decomposing the function into orthogonal parametric (fixed-effect) and nonparametric (random-effect) components, so that the significance of the parametric part can be tested independently of the test of the nonparametric deviation. TAPS provides both Wald and score tests: the Wald test applies to all outcome families supported by `mgcv`, while the score test typically offers greater power. Given its superior Type I error control and higher power, the score test is recommended as the default, with the Wald test as a practical alternative for non-exponential-family outcomes. Finally, we implement TAPS in the R package `mgcv.taps`, tightly integrated with `mgcv`, allowing users to apply it with minimal additional learning, while supporting diverse outcome types and scaling efficiently to biobank-scale data.

In the first analysis, 78% (36/46) exhibited significant age-varying effects of PRSs, indicating that age modifies the genetic architecture for most complex traits analyzed (Sung et al. 2018, Winkler et al. 2024). For 44.4% of these 36 traits, the age-varying pattern could not be adequately captured by a simple linear age-PRS interaction, suggesting substantial heterogeneity in how genetic influences evolve across the life course. Similar heterogeneity has been reported for blood pressure, BMI, and metabolic biomarkers, where genetic effects may accelerate, plateau, or even reverse direction at specific life stages (Hottenga et al. 2005, Guindo-Martínez et al. 2021, Jiang et al. 2021). Several traits were also well approximated by a parsimonious piecewise linear structure, where the linear interaction may hold within certain age ranges but with different slopes, or even opposite signs, between younger and older individuals, as observed for SBP, DBP, and PP. These results suggest that G×E interaction studies could gain power by stratifying analyses by age and other environmental factors (Domingue et al. 2020). Nevertheless, the biological underpinnings of the observed changepoints remain unclear and warrant further investigation.

In the second analysis, we found that retirement at age 65 was significantly associated with structural changes in age-varying PRS effects for only a small subset of traits: MONO, TP, and WBC. To our knowledge, this is the first evidence that retirement may interact with genetic effects to shape health-related phenotypes. For these inflammatory and protein markers, retirement may reduce chronic psychosocial stress (Eibich 2015), which has been linked to lower systemic inflammation in older adults (Steptoe et al. 2007). These conjectures may require future validation in trials or longitudinal studies. Compared with prior work (Eibich 2015, Furuya & Fletcher 2024), which examined direct effects of retirement on health indicators, our analysis is novel in testing whether retirement coincides with structural changes in PRS effects, highlighting a previously unexplored aspect of G×E interaction.

Our two analyses also highlight important limitations in the current application of PRSs. Specifically, a growing body of literature has shown that the predictive power and risk associations of PRSs differ across age strata (Khan et al. 2022). Our findings suggest that this pattern may arise because the effects of PRSs themselves are continuously age-varying rather than constant over the lifespan. Such patterns may partly reflect risk process mechanisms, in which genetic risk factors are present from birth while non-genetic factors accumulate over time, and statistical interactions between them can dilute the apparent genetic contribution in a manner analogous to frailty, without requiring age-dependent changes in the underlying genetic effects (Falconer 1967, Jiang et al. 2021). Current PRSs can only capture age-averaged or absolute accumulated polygenic effects (Chatterjee et al. 2016). We thus speculate that much of the observed age variation in PRS effects may reflect differences in the predictive power of absolute PRSs across age groups, although novel biological mechanisms cannot be excluded. In the future, constructing PRSs based on age-stratified GWAS or explicitly modeling age-varying genetic effects may enable more targeted disease risk prediction across different age groups, thereby enhancing the practical utility of PRSs (Chatterjee et al. 2016).

Several cautions regarding the use and interpretation of TAPS are worth noting. First, TAPS requires users to specify the parametric structure to be tested, including hyperparameters such as the location of a jump or change-of-slope. We should emphasize that the utility of TAPS is not designed to discover the best-fitting structure from data, but rather to statistically test whether a given structural form can sufficiently describe the variation of the data, for reasons of interpretability and generalizability. In addition, in some applications like RDD and RKD, as well as the retirement analysis, these locations correspond to cutoffs of treatment assignments and are not subject to tuning (Huntington-Klein 2021). However, we acknowledge the importance of discovering structural forms rather than requiring them to be pre-specified, and future extensions of TAPS could incorporate structural change detection methods (Bai & Perron 1998, Muggeo 2003, Killick et al. 2012, Zhang & Li 2023, Chen et al. 2023). Second, because our motivating applications arise from genetic studies, it may be tempting to view TAPS as a variance-component mixed-effects method, akin to SKAT (Wu et al. 2011), STAAR (Li et al. 2022), GREML (Yang et al. 2010, 2014), or G×EMM (Dahl et al. 2020). These approaches are designed to quantify or test the contribution of a genetic component through its random-effect variance, typically using a genetic relatedness matrix (GRM) constructed from a very large number of variants, which is often full or nearly full rank. In contrast, TAPS addresses a different inferential target: it is a GAM-based hypothesis test of whether a prespecified parametric structure provides an adequate description of a non-linear effect. Accordingly, the kernel matrix in TAPS does not represent genome-wide genetic similarity, but rather parameterizes deviations from the null parametric form within a controlled smooth space. This kernel is typically low-rank, as TAPS applies low-rank kriging and PCA for dimension reduction. The use of orthogonal constraints allows the fixed and random components to be tested separately. Third, although TAPS favors the more powerful score test, its empirical performance in this work has only been investigated under standard exponential-family outcome models. Extending the score test to settings such as non-exponential-family outcomes, variance heteroskedasticity, and quantile regression (Wood et al. 2016, Fasiolo et al. 2021) remains an important direction for future research, as ignoring these departures may lead to invalid inference.

## Data Availability

All GWAS summary statistics used in this study are publicly available, with their sources listed in Table S1. Given the large number of traits analyzed, we have implemented an interactive Shiny web interface (https://ovlwff-yihe-yang.shinyapps.io/visualization_of_taps/) to visualize all plots. The individual-level data from the UK Biobank used for analyses are available under Application ID: 81097.

https://www.nealelab.is/uk-biobank

## Acknowledgments

We thank the editors and anonymous reviewers for their constructive comments and suggestions, which have significantly improved the manuscript. The first author is grateful to Dr. Jianxin Pan for insightful discussions during the conceptualization of this work. We also extend our gratitude to Dr. Simon Wood for developing the R package `mgcv`, which facilitates user-defined smooth terms and enables seamless methodological extensions. This work was supported by grants HG011052 and HG011052-03S1 (to X.Z.) from the National Human Genome Research Institute (NHGRI), USA.

## Data and Code Availability

All GWAS summary statistics used in this study are publicly available, with their sources listed in Table S1. The individual-level data from the UK Biobank used for analyses are available under Application ID: 81097.

Given the large number of traits analyzed, we have implemented an interactive Shiny web interface (https://ovlwff-yihe-yang.shinyapps.io/visualization_of_taps/) to visualize all plots. The R package `mgcv.taps`, which implements the TAPS method, is available at https://github.com/harryyiheyang/mgcv.taps. All analysis code used in this paper is available at https://github.com/harryyiheyang/mgcv.taps.code.

## SUPPLEMENTARY MATERIAL

### Supplementary Materials

Supplementary Materials contain the supplementary methodology, simulations, and real data analyses.

### Supplementary Tables

Supplementary Tables contain multiple tables recording the results of real data analysis.

## Notes

### Competing Interest Statement

The authors have declared no competing interest.

### Summary of Updates

Updates according to reviewers' comments

