## Supplementary Materials for "Testing Parametric Structure in Genetic Age-Effect Curves: A GAM-Based Framework with Application to the UK Biobank"

#### 1 Supplementary Method

In Supplementary Method, we show the detailed methodology of TAPS.

#### 1.1 Notation

Span($\mathbf{A}$) denotes the space spanned by the columns of $\mathbf{A}$ and $\overline{\text{span}}(\mathbf{A})$ is the complementary space. Besides, $\mathrm{I}(\cdot)$ is the indicator function, $\langle \cdot, \cdot \rangle$ is the inner product operator, and $(\cdot)_+ = \max(0, \cdot)$ is the linear segmented function. In addition, $f^{(q)}$ is the $q$th derivative of $f$. All the functions are defined in the interval [0,1].

#### 1.2 Test with arbitrary parametric structure

A standard GAM takes the following representation

$$y_i \sim \mathbf{EF}(\mu_i, \phi_0), \quad g(\mu_i) = \eta_i = \boldsymbol{Z}_i^\top \boldsymbol{\gamma} + f_1(x_{i1}) + \cdots + f_J(x_{iJ}), \tag{S1}$$

where $y_i$ is a response variable belonging to an exponential family distribution $\mathbf{EF}(\mu_i, \phi_0)$ with mean $\mu_i$ and dispersion parameter $\phi_0$, $g$ is a known link function, $\eta_i$ is a linear predictor, $\boldsymbol{Z}_i$ is the $i$th row of a parametric model matrix $\mathbf{Z}$, $\boldsymbol{\gamma}$ is a regression coefficient vector, and $f_j$ is a smooth function of covariate $x_{ij}$.

Suppose that $A_1(x), \ldots, A_q(x)$ are the basis functions of a parametric structures and $\mathcal{H}_A = \text{span}(A_1, \ldots, A_q)$ is the space spanned by these bases. The common parametric structures include $\{A_1(x) = 1, A_2(x) = x\}$ for linearity, $\{A_1(x) = 1, A_2(x) = x, A_3(x)) = (x - 0.5)_+\}$ for linearity with a change-point $\nu_1 = 0.5$, and $\{A_1(x) = 1, A_2(x) = \mathrm{I}(x > 0.5), A_3(x) = (x-0.5), A_4(x) = (x-0.5)\mathrm{I}(x > 0.5)\}$ for linearity with a breakpoint $\nu_0 = 0.5$. The null and alternative hypotheses that TAPS addresses are

$$\mathrm{H}_0 : f \in \mathcal{H}_A \qquad \text{v.s.} \qquad \mathrm{H}_1 : f \in \mathcal{H} \text{ and } f \notin \mathcal{H}_A, \tag{S2}$$

where $\mathcal{H}$ is a general functional space. In application, $\mathcal{H}$ should be wide enough to fully cover $\mathcal{H}_A$ (i.e., $\mathcal{H}_A \subsetneqq \mathcal{H}$) which may involve nonsmooth functions. On the other hand, $\mathcal{H}$ should also be able to describe common smooth functions in case $\mathcal{H}_A$ is misspecified. To make this hypothesis testable, TAPS constructs a new representation of $f(x)$:

$$f(x) = \boldsymbol{A}(x)^\top \boldsymbol{\alpha} + b(x), \tag{S3}$$

where $\boldsymbol{A}(x) = (A_1(x), \ldots, A_q(x))^\top$ is the basis function vector, $\boldsymbol{\alpha} = (\alpha_1, \ldots, \alpha_q)^\top$ is a fixed vector, $b(x)$ is a Gaussian process with covariance function

$$\texttt{cov}(b(x_i), b(x_j)) = \phi K(x_i, x_j), \tag{S4}$$

$K(x_i, x_j)$ is a known bivariate function, and $\phi \geq 0$ is the variance. Here, we made use of the equivalence between Reproducing Kernel Hilbert Spaces (RKHS) and Gaussian processes: a function $b(x)$ that belongs to an RKHS $\mathcal{H}_K$ with kernel function $K(x_i, x_j)$ is mathematically equivalent to a Gaussian process $b(x)$ in the $\mathcal{L}_2$ space with correlation function $K(x_i, x_j)$. In addition, $b(x)$ is not unrestricted; it must satisfy the following orthogonality condition:

$$\int_R b(x) A_i(x) dx = 0, \quad i = 1, \ldots, p. \tag{S5}$$

This constraint ensures identifiability of the model. Without it, there would be non-unique decompositions of the effect between the fixed components $A_i(x)$ and the smooth/random function $b(x)$. From the perspective of Gaussian process modeling, this condition implies that $b(x)$ can be interpreted as a stochastic component that is orthogonal to the fixed-effect space spanned by $A_i(x)$. In other words, $b(x)$ captures only the residual variation that cannot be explained by the fixed basis functions, ensuring that it does not overlap or interfere with their contribution. Thus, for any function belonging to $\mathcal{H} = \mathcal{H}_A \cup \mathcal{H}_K$, the hypothesis (S2) reduces to

$$\mathrm{H}_0 : \phi = 0 \qquad \text{v.s.} \qquad \mathrm{H}_1 : \phi > 0. \tag{S6}$$

As a result, the techniques of GLMM such as score test [19] can be applied to yield the $p$-value of (S6).

#### 1.3 Smoothing Polynomial Splines

The relationship between the new mixed-effects representation (S3) and the well-known mixed-effects representation of the smoothing spline [7] is as follows. Specifically, Kimeldorf and Wahba [7] showed that any function belonging to the Sobolev-Hilbert space

$$\mathcal{W}_q = \{f:\ f, f', \ldots, f^{q-1} \text{ are absolutely continuous, } f^q \text{ is bounded}\} \tag{S7}$$

can be described by a mixed-effects model that shares the same representation as (3) in the main text, where the fixed parametric effect is a $(q-1)$-order polynomial function and the random effect is a $q$-fold integrated Wiener process with covariance function:

$$K(x_i, x_j) = \int_0^1 \{(x_i - u)_+^{q-1}(x_j - u)_+^{q-1}\}/((q-1)!)^2 \mathrm{d}u. \tag{S8}$$

In particular, the polynomial and the integrated Wiener process are two strictly orthogonal components as long as the boundary condition:

$$f(0) = f'(0) = \cdots = f^{q-1}(0) = 0 \tag{S9}$$

is satisfied, awarding the smoothing spline an excellent statistical interpretability. One of the contributions of TAPS is showing how to construct the random effect $b(x)$ in a way that ensures it is orthogonal to any given fixed parametric effect. See more details of polynomial smoothing spline in Wahba [11].

The mixed-effects representation of the smoothing spline can be regarded as a specific example of (S3), in which the fixed parametric effect is a $(q-1)$-order polynomial and the random effect is a $q$-fold integrated Wiener process whose covariance function is (S8). The polynomial and the integrated Wiener process have been proved to be two strictly orthogonal components as long as the so-called boundary condition [13] is satisfied, awarding the smoothing spline an excellent statistical interpretability. Inspired by it, we specify a new constraint (S16) such that the random effect $b(x)$ is strictly orthogonal to the fixed parametric effect $\boldsymbol{A}(x)^\top \boldsymbol{\alpha}$.

#### 1.4 Construction of New Mixed-Effects Representation

The construction of $b(x)$ starts from representing it using the RKHS [13], under the alternative hypothesis that $b(x)$ is a general smooth function. According to the functional analysis theory, every RKHS $\mathcal{H}_K$ corresponds to a unique semi-positive definite kernel function $K(\cdot,\cdot)$ defined in the Cartesian product space $[0,1]\times[0,1]$. For any function $b \in \mathcal{H}_K$,

$$\langle b, K(\cdot, x)\rangle_{\mathcal{H}_K} = b(x), \tag{S10}$$

where $\langle\cdot,\cdot\rangle_{\mathcal{H}_K}$ is the inner product in the space $\mathcal{H}_K$. This property ensures that for $n$ observation $\{x_1, \ldots, x_n\} \subset [0,1]$, there must exist an $(n\times n)$ matrix $\mathbf{K}$ with the $(i,j)$th element being $K(x_i, x_j)$ and an $(n \times 1)$ vector $\boldsymbol{\theta} = (\theta_1, \ldots, \theta_n)^\top$ such that

$$\boldsymbol{b} = \mathbf{K}\boldsymbol{\theta}, \tag{S11}$$

where $\boldsymbol{b} = (b(x_1), \ldots, b(x_n))^\top$. When a specific RKHS $\mathcal{H}_K$ is determined, the null and alternative hypotheses that TAPS shift to

$$\mathrm{H}_0: f \in \mathcal{H}_A \qquad \text{v.s.} \qquad \mathrm{H}_1: f \in \mathcal{H}_K \text{ and } f \notin \mathcal{H}_A, \tag{S12}$$

where the alternative hypothesis proposes that if the target function deviates from the specified parametric structure, it is instead a smooth function belonging to $\mathcal{H}_K$. In practice, most of the commonly used kernel functions can generate a general smooth function, such as the Gaussian kernel, spherical kernel, the Mat'ern family correlation function, and the kernels of the Sobolev space [10]. In addition, the expression (S11) also holds for multivariate function $\boldsymbol{f} = (f(\boldsymbol{x}_1), \ldots, f(\boldsymbol{x}_n))^\top$ where $\boldsymbol{x}_i$ is a multivariate vector. In this case, the

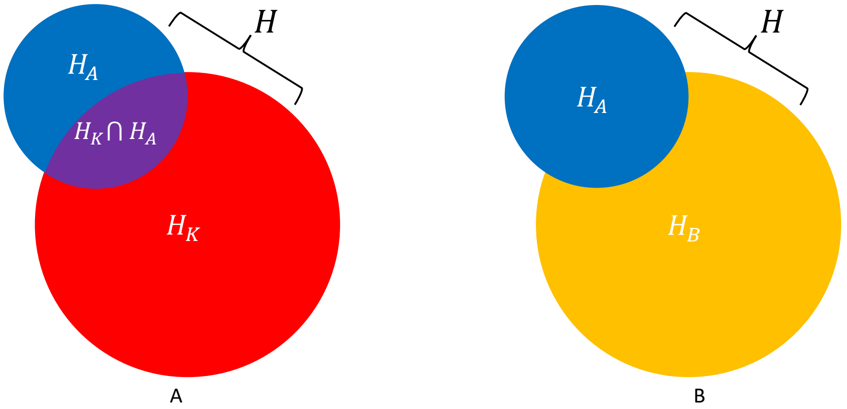

Figure S1: Illustration of principle to construct $b(x)$. A: the red circle represents the RKHS $\mathcal{H}_K$, the blue circle represents the parametric space $\mathcal{H}_A$, and the general functional space in (S2) is defined as $\mathcal{H} = \mathcal{H}_K \cup \mathcal{H}_A$. B: the yellow region, which represents $\mathcal{H}_K/\mathcal{H}_A$, is the functional space of the random effect $b(x)$.

kernel function $K(\boldsymbol{x}_i, \boldsymbol{x}_j)$ is usually chosen as the isotropic kernel function, i.e., $K(\boldsymbol{x}_i, \boldsymbol{x}_j) = K(||\boldsymbol{x}_i - \boldsymbol{x}_j||)$ with a certain norm $||\cdot||$.

Figure S1 illustrates the principle of enforcing $b(x)$ to be orthogonal to $\boldsymbol{A}(x)^\top \boldsymbol{\alpha}$. In panel A, the red circle denotes the RKHS $\mathcal{H}_K$ and the blue circle the parametric space $\mathcal{H}_A$, with the overall function space defined as $\mathcal{H} = \mathcal{H}_K \cup \mathcal{H}_A$. In panel B, the yellow subspace $\mathcal{H}_B$ is the orthogonal complement of $\mathcal{H}_A$ within $\mathcal{H}_K$. Thus, $b(x)$ lies in $\mathcal{H}_B$ and can be expressed using its basis functions, reducing the problem to finding a basis for $\mathcal{H}_B$. We construct the basis functions of $\mathcal{H}_B$ by finding an $((n+q)\times n)$ orthogonal matrix $\mathbf{Q}$ satisfying the following linear constraint

$$\mathbf{A}^\top \mathbf{F}\mathbf{Q} = \mathbf{0}_{q\times n}, \tag{S13}$$

where $\mathbf{F} = (\mathbf{A}, \mathbf{K})$ is the combination of $\mathbf{A}$ and $\mathbf{K}$. Here, the orthogonal matrix $\mathbf{Q}$ describes a special linear transformation that resorts the columns of $\mathbf{F}$, i.e., the basic functions of $\mathcal{H}$, into a new one perpendicular to $\mathbf{A}$. As a result, the columns of $\mathbf{FQ}$ can be regarded as the basis functions of $\mathcal{H}_B$ and thus it can represent $\boldsymbol{b} = \mathbf{FQ}\boldsymbol{\pi}$ where $\boldsymbol{\pi}$ is an unconstrained vector in $\mathbb{R}^n$. The QR decomposition can yield a candidate of the orthogonal matrix $\mathbf{Q}$. Consider the following QR decomposition:

$$\mathbf{F}^\top \mathbf{A} = (\mathbf{Q}_1, \mathbf{Q}_2)\begin{pmatrix}\mathbf{R}_1\\ \mathbf{0}_{n\times q}\end{pmatrix}, \tag{S14}$$

where $\mathbf{Q}_1$ is an $((n+q)\times q)$ orthogonal matrix, $\mathbf{Q}_2$ is an $((n+q)\times n)$ orthogonal matrix, and $\mathbf{R}_1$ is an $(q\times q)$ upper triangular matrix. Here $\mathbf{Q}_1$ is an orthogonal basis matrix of $\text{span}(\mathbf{F}^\top\mathbf{A})$ and $\mathbf{Q}_2$ is an orthogonal basis matrix of $\overline{\text{span}}(\mathbf{F}^\top\mathbf{A})$, thus we can choose $\mathbf{Q} = \mathbf{Q}_2$. Note that other numerical tools such as the singular values decomposition (SVD) can also yield an orthogonal basis matrix of $\overline{\text{span}}(\mathbf{F}^\top\mathbf{A})$, which means $\mathbf{Q}$ is not unique. In contrast, the column space of $\mathbf{FQ}$, i.e., the space spanned by the left singular vectors of $\mathbf{FQ}$, is unique.

#### 1.5 Prior Distriubtion of New Mixed-Effects Representation

The prior distribution of $b(x)$ can be constructed as follows. Specifically, since $\boldsymbol{b}$ also falls within $\mathcal{H} = \mathcal{H}_A \cup \mathcal{H}_K$, it can be alternatively represented as

$$\boldsymbol{b} = \mathbf{F}\tilde{\boldsymbol{\vartheta}} = \mathbf{A}\tilde{\boldsymbol{\alpha}} + \mathbf{K}\tilde{\boldsymbol{\theta}}, \tag{S15}$$

where the vector $\tilde{\boldsymbol{\vartheta}} = (\tilde{\boldsymbol{\alpha}}^\top, \tilde{\boldsymbol{\theta}}^\top)^\top$. The constraint to ensure $\boldsymbol{b}$ fully fall within $\mathcal{H}_B$ is

$$\mathbf{A}^\top \mathbf{F}\tilde{\boldsymbol{\vartheta}} = \mathbf{0}_{q\times 1}. \tag{S16}$$

This constraint is similar to the boundary condition of the polynomial smoothing spline [13], but it has a clearer meaning: the random effect $\boldsymbol{b}$ represented by $\mathbf{F}\tilde{\boldsymbol{\vartheta}}$ is perpendicular to the fixed parametric effect $\mathbf{A}\boldsymbol{\alpha}$. In addition, this constraint guarantees that $\tilde{\boldsymbol{\vartheta}}$ must be a vector in $\overline{\text{span}}(\mathbf{F}^\top\mathbf{A})$. Therefore, given the $\mathbf{Q}$ whose columns are the basis of $\overline{\text{span}}(\mathbf{F}^\top\mathbf{A})$, there must exist the $(n \times 1)$ vector $\boldsymbol{\pi}$ such that $\tilde{\boldsymbol{\vartheta}} = \mathbf{Q}\boldsymbol{\pi}$. In other words, any $\boldsymbol{b} = \mathbf{FQ}\boldsymbol{\pi}$ where $\mathbf{Q}$ is the basis matrix of $\overline{\text{span}}(\mathbf{F}^\top\mathbf{A})$ and $\boldsymbol{\pi}$ is an unconstrained vector in $\mathbb{R}^n$, can be represented as $\boldsymbol{b} = \mathbf{F}\tilde{\boldsymbol{\vartheta}}$ where $\tilde{\boldsymbol{\vartheta}}$ is subject to the linear constraint (S16).

On the other hand, a smooth function $b(x) \in \mathcal{H}_K$ is statistically equivalent to a Gaussian process with zero mean and covariance function $\texttt{cov}(b(x_i), b(x_j)) = \phi K(x_i, x_j)$ in $L_2$ space [6]. Such a Bayesian interpretation stems from the kriging method used in geostatistical analysis [2]. Theorem 12.11 in Wainwright [13] ensures $\mathbf{K}$ is a semipositive definite matrix so that the above Gaussian distribution is valid. Based on this random effect representation of smooth functions in RKHS, the augmented vector $\tilde{\boldsymbol{\vartheta}}$ should follow $\mathcal{N}(\mathbf{0}_{(n+q)\times 1}, \phi\tilde{\mathbf{K}}^+)$ where $\tilde{\mathbf{K}} = \text{diag}(\mathbf{0}_{q\times q}, \mathbf{K})$ is an $((n+q) \times (n+q))$ block-diagonal matrix. The block corresponding to $\tilde{\boldsymbol{\alpha}}$ is a zero matrix because $\mathbf{A}\tilde{\boldsymbol{\alpha}}$ is a fixed effect and hence this is no randomness of $\tilde{\boldsymbol{\alpha}}$. Furthermore, since $\tilde{\boldsymbol{\vartheta}} = \mathbf{Q}\boldsymbol{\pi}$, the prior distribution of $\boldsymbol{\pi}$ is then

$$\boldsymbol{\pi} \sim \mathcal{N}(\mathbf{0}_{n\times 1}, \phi(\mathbf{Q}^\top\tilde{\mathbf{K}}\mathbf{Q})^+). \tag{S17}$$

Subsequently, since $\boldsymbol{b} = \mathbf{FQ}\boldsymbol{\pi}$, the prior distribution of $\boldsymbol{b}$ is

$$\boldsymbol{b} \sim \mathcal{N}(\mathbf{0}_{n\times 1}, \phi\mathbf{FQ}(\mathbf{Q}^\top\tilde{\mathbf{K}}\mathbf{Q})^+\mathbf{Q}^\top\mathbf{F}^\top). \tag{S18}$$

Thus, testing whether $\boldsymbol{f} = \mathbf{A}\boldsymbol{\alpha}$ reduces to testing whether $\phi = 0$.

#### 1.6 Dimension Reduction of New Mixed-Effects Representation

The construction of $\boldsymbol{b}$ involves the QR-decomposition of the $((n+q) \times q)$ matrix $\mathbf{F}^\top\mathbf{A}$, which is costly if $n$ is large. Besides, the dimension of the vector $\boldsymbol{\pi}$ is $n$, which is easy to cause the curse of dimensionality in practice. To resolve such numerical problems, we apply two dimension-reducing techniques to simplify the representation of $\boldsymbol{b}$.

First, we employ the low-rank RKHS [5] to represent a general smooth function. Based on a set of pre-given knots $\{\kappa_1, \ldots, \kappa_p\} \subset \{x_1, \ldots, x_n\}$, the low-rank RKHS has the same representation as the standard one, i.e.,

$$\boldsymbol{b} = \mathbf{K}\boldsymbol{\theta},$$

but the kernel matrix $\mathbf{K}$ here is a $(n \times p)$ matrix with the $(i, j)$th entry $K(x_i, \kappa_j)$. In practice, the knots are usually set as the $1/p, \ldots, p/p$ quantiles of $\boldsymbol{x}$. Besides, the prior distribution of the $(p \times 1)$ vector $\boldsymbol{\theta}$ is $\mathcal{N}(\mathbf{0}_{p\times 1}, \phi\boldsymbol{\Omega}^+)$ where $\boldsymbol{\Omega}$ is a $(p \times p)$ matrix with the $(i, j)$th entry $K(\kappa_i, \kappa_j)$. Ruppert, Wand, and Carroll [10] showed that the low-rank RKHS is almost the same accuracy as the standard but can give considerable computational payoffs in implementation. We take the same strategy to find the orthogonal matrix $\mathbf{Q}$ such that $\mathbf{A}^\top\mathbf{FQ} = \mathbf{0}$ where $\mathbf{F} = (\mathbf{A}, \mathbf{K})$ is an $(n \times (p+q))$ matrix. The random effect now is given by $\boldsymbol{b} = \mathbf{FQ}\boldsymbol{\pi}$ where $\boldsymbol{\pi}$ has a prior distribution $\mathcal{N}(\mathbf{0}_{p\times 1}, \phi(\mathbf{Q}^\top\tilde{\boldsymbol{\Omega}}\mathbf{Q})^+)$ with $\tilde{\boldsymbol{\Omega}} = \text{diag}(\mathbf{0}_{q\times q}, \boldsymbol{\Omega})$.

In practice, the number of knots $p$ should be large enough to guarantee that the bias of the representation is sufficiently small. Ruppert, Wand, and Carroll [10] suggested selecting $p = \min(35, n/4)$ and verified it is sufficient to fit most common smooth functions. However, setting any other moderately large value will still lead to a serious curse of dimensionality when there are multiple functions to be estimated in a model. To remove this obstacle, we further reduce the dimension of $\mathbf{FQ}$ using the strategy of Wood [16]. Consider the following singular values decomposition (SVD):

$$\mathbf{FQ} = \mathbf{UDV}^\top, \tag{S19}$$

where $\mathbf{D}$ is an $(p \times p)$ diagonal matrix composed of the singular values of $\mathbf{FQ}$, $\mathbf{U}$ is an $(n \times p)$ orthogonal matrix, and $\mathbf{V}$ is an $(p \times p)$ orthogonal matrix. Usually, only the first few singular values are large, which means the most information of $\mathbf{FQ}$ is carried by the first few left singular vectors. By selecting an appropriate cutoff $k$,

$$\mathbf{FQ} \approx \mathbf{U}_k\mathbf{D}_k\mathbf{V}_k^\top \tag{S20}$$

where $\mathbf{U}_k$ and $\mathbf{V}_k$ consist of the first $k$ columns of $\mathbf{U}$ and $\mathbf{V}$, respectively, and $\mathbf{D}_k$ is the first $(k \times k)$ submatrix of $\mathbf{D}$. The function $\boldsymbol{f}$ is then approximated by

$$\boldsymbol{f} \approx \mathbf{A}\boldsymbol{\alpha} + \mathbf{B}\boldsymbol{\beta}, \tag{S21}$$

where $\mathbf{B} = \mathbf{U}_k$ and $\boldsymbol{\beta} = \mathbf{D}_k \mathbf{V}_k^\top \boldsymbol{\pi}$. In this representation, $\boldsymbol{\beta}$ can be regarded as a Gaussian variable with a prior distribution $\mathcal{N}(\mathbf{0}_{k\times 1}, \phi\boldsymbol{\Theta}^+)$ where

$$\boldsymbol{\Theta} = \mathbf{D}_k^{-1}\mathbf{V}_k^\top \mathbf{Q}^\top \tilde{\boldsymbol{\Omega}} \mathbf{Q} \mathbf{V}_k \mathbf{D}_k^{-1}. \tag{S22}$$

Thereby $\boldsymbol{b} \approx \mathbf{B}\boldsymbol{\beta}$ whose prior distribution is $\mathcal{N}(\mathbf{0}_{n\times 1}, \phi\mathbf{B}\boldsymbol{\Theta}^+\mathbf{B}^\top)$. With the mixed-effects representation (S21), the so-called general functional space $\mathcal{H}$ is approximated by $\mathcal{H} = \mathcal{H}_A \oplus \mathcal{H}_B$ where $\mathcal{H}_A = \mathrm{span}(\mathbf{A})$ and $\mathcal{H}_B \approx \mathrm{span}(\mathbf{B})$.

The bias of this representation is $\boldsymbol{\delta} = \mathbf{F}\mathbf{Q}\boldsymbol{\pi} - \mathbf{B}\boldsymbol{\beta}$ which declines as $k$ increases. Wood [16] showed it can accurately fit a common function by setting $k = 10$. Besides, unlike the regression spline that requires selecting $k$ and the locations of knots carefully, our representation is affected by neither $k$ nor the locations of knots, as long as $k$ and $p$ are appropriately large. In terms of regulating the wiggliness of the final function estimator, the variance component $\phi$ is more essential than $k$ and the locations of knots.

#### 1.7 Patterns of New Mixed-Effects Representation

We demonstrate with respect to different $\mathbf{A}$, what the main patterns of the columns of $\mathbf{B}$ are. Without losing generality, we set the covariate to be $\boldsymbol{x} = (1/400, 2/400, \ldots, 1)^\top$ and the knots to be $\boldsymbol{\kappa} = (1/200, 2/200, \ldots, 1)^\top$.

##### 1.7.1 Isotropic kernel function

We first introduce the isotropic kernel function. The isotropic kernel function refers to some special kind of kernel function that actually varies only with the distance between its two input variables, i.e., $K(x, y) = K(|x - y|)$. Among all isotropic kernel functions, the kernel functions with stationary property are called the Gaussian process kernel functions, correlation functions and so on. Common stationary kernel functions include the exponential kernel function, Gaussian kernel function, trigonometric kernel function, spherical kernel function, Mat{'e}rn family kernel functions, etc. The Mat{'e}rn family kernel function may be the most commonly used isotropic function, whose general expression is

$$K(x, y) = \frac{|x - y|^\nu \mathcal{B}_\nu(|x - y|)}{2^{\nu - 1}\Gamma(\nu)}, \tag{S23}$$

where $\mathcal{B}_\nu(\cdot)$ is the modified $\nu$-order Bessel function. For general order $\nu > 0$, $\mathcal{B}_\nu(\cdot)$ has no general expression. Fortunately, for $\nu = m + \frac{1}{2}$, $m = 1, 2, \ldots$, the function has an explicit expression. For details, see table S1 and table S2.

On the other hand, since the distance $|x - y|$ has a different range of values depending on the units of $x$ and $y$. Therefore, we will also introduce a scale parameter $\rho$ to make the isotropic kernel function more adaptable. For example, for the exponential kernel function, we define the modified exponential kernel function

$$K_\rho(x, y) = \exp\left(-\frac{|x - y|}{\rho}\right),$$

where $\rho > 0$ is the so-called scale parameter, which can generally be simply chosen as $\rho = c_0\widehat{\mathrm{std}}(x)$, and $c_0 > 0$. For the other isotropic kernel functions, we also do a similar adjustment to the exponential kernel function.

Table S1: Some isotropic (correlation) functions with corresponding spectral densities

| | isotropic functions | spectral density functions |
|---|---|---|
| Exponential kernel | $\exp(-\lvert x-y\rvert)$ | $\frac{1}{\pi(1+x^2)}$ |
| Gaussian kernel | $\exp(-\lvert x-y\rvert^2/2)$ | $\frac{1}{\sqrt{2\pi}}\exp(-\frac{x^2}{2})$ |
| Trigonometric kernel | $(1-\lvert x-y\rvert)_+$ | $\frac{1-\cos(x)}{\pi x^2}$ |
| Spherical kernel | $(1-\frac{3}{2}\lvert x-y\rvert+\frac{1}{2}\lvert x-y\rvert^3)\mathbf{1}_{\lvert x-y\rvert<1}$ | no explicit density function |

Table S2: Matérn isotropic function with parameters $\nu$ =0.5, 1.5, 2.5, 3.5

| | isotropy function |
|---|---|
| $\nu=0.5$ | $\exp(-\lvert x-y\rvert)$ |
| $\nu=1.5$ | $\exp(-\lvert x-y\rvert)(1+\lvert x-y\rvert)$ |
| $\nu=2.5$ | $\exp(-\lvert x-y\rvert)(1+\lvert x-y\rvert+\frac{1}{3}\lvert x-y\rvert^2)$ |
| $\nu=3.5$ | $\exp(-\lvert x-y\rvert)(1+\lvert x-y\rvert+\frac{2}{5}\lvert x-y\rvert^2+\frac{1}{15}\lvert x-y\rvert^3)$ |

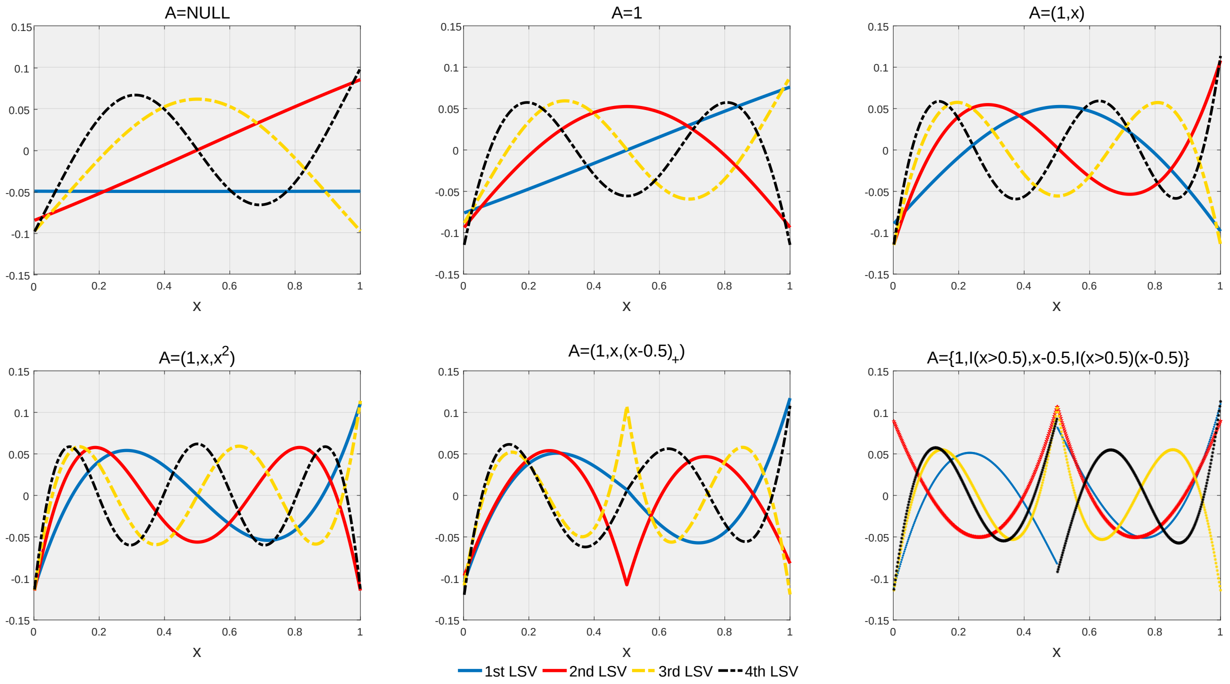

Figure S2: Matérn family kernel function with $\nu = 1.5$ and $\rho = 5$. The six panels show the patterns of each column of the orthogonal matrix $\mathbf{B}$ obtained when "fixed effects"$\mathbf{A}$ are taken for different cases. LSV refers to the left singular vector.

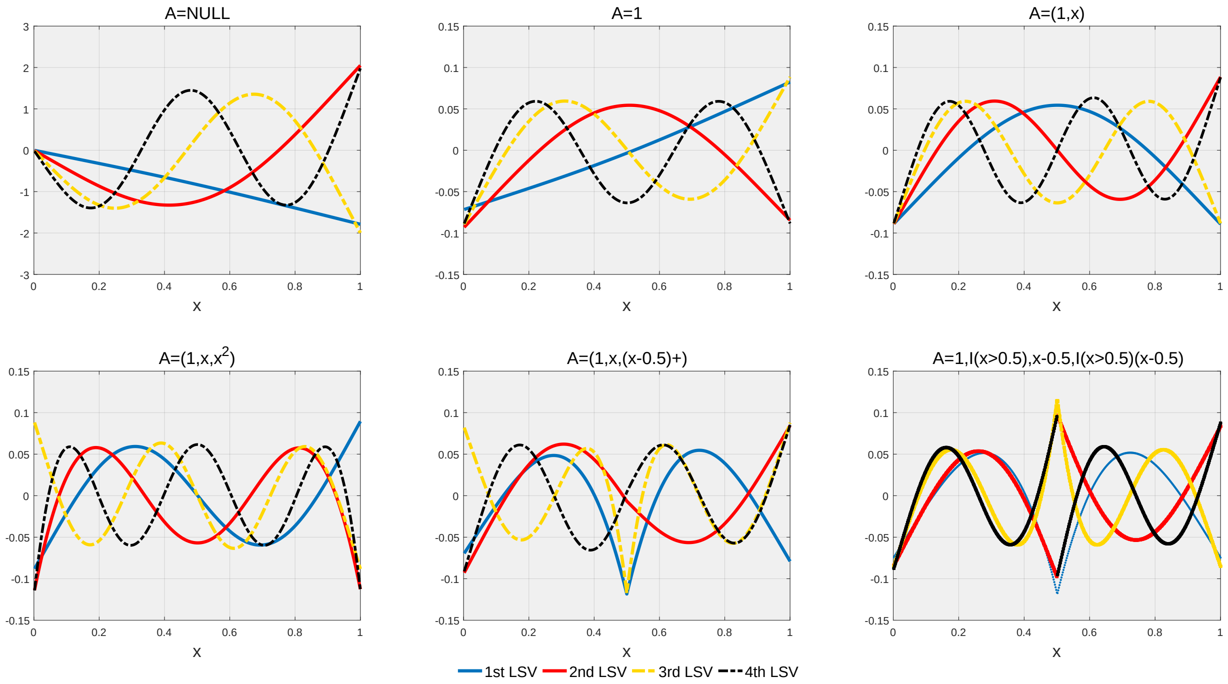

Figure S3: Wahba cubic smoothing spline. The six panel show the patterns of each column of the orthogonal matrix $\mathbf{B}$ obtained when "fixed effects"$\mathbf{A}$ are taken for different cases. LSV refers to the left singular vector.

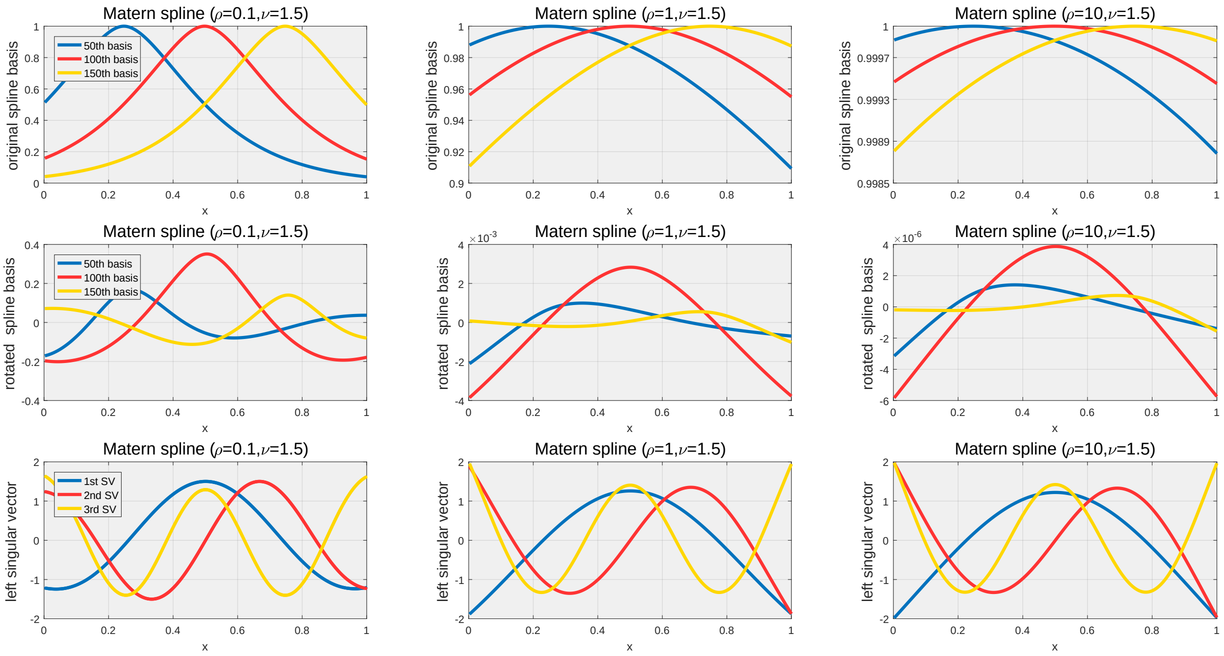

Figure S4: The first row of three plots shows the Matérn kernel basis functions for different parameters $\rho$, $\nu = 1.5$. The second row of three plots then shows these basis functions obtained by the rule $\mathbf{A}^\top\mathbf{KQ} = \mathbf{0}$ where $\mathbf{A} = (\mathbf{1}, \mathbf{x})$ to obtain the rotated basis function matrix $\mathbf{KQ}$ of basis functions. The third row of three plots then shows the principal components of the rotated basis function matrix $\mathbf{KQ}$ extracted by PCA.

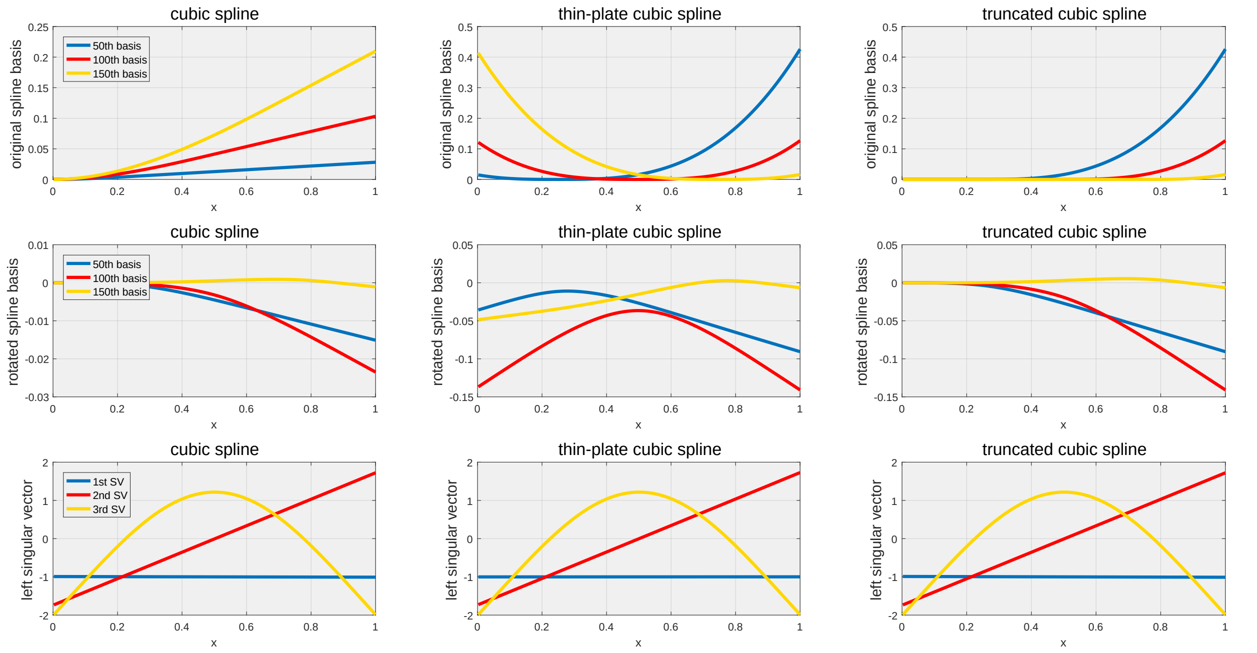

Figure S5: The three subplots in the first row show the images of the original spline basis functions for the Wahba cubic spline, the thin-sliced cubic spline, and the truncated cubic spline. The three subplots in the second row show the images of the functions of the rotated spline basis $\mathbf{K}_1\mathbf{Q}$. The three subplots in the third row show the first three columns of $\sqrt{n}\mathbf{B}$ constructed by the simultaneous diagonalization method. The first, second columns of them must be $\mathbf{1}$ and $2\sqrt{3}(\mathbf{x}-0.5)$.

##### 1.7.2 Smoothing Spline

Smoothing polynomial spline is a RKHS method corresponding to a so-called Sobolev-Hilbert space $\mathcal{W}$ [7]. Consider the following two kernel functions defined on $[0,1]\times[0,1]$:

$$K_0(x_i,x_j)=\sum_{l=0}^{q-1}\frac{x_i^l x_j^l}{[l!]^2},\quad K_1(x_i,x_j)=\int_0^1\frac{(x_i-u)_+^{q-1}(x_j-u)_+^{q-1}}{[(q-1)!]^2}\mathrm{d}u, \tag{S24}$$

where $q\geq 1$ is a given integer. The Sobolev-Hilbert space $\mathcal{W}=\mathcal{H}_0\oplus\mathcal{H}_1$, where $\mathcal{H}_0$ and $\mathcal{H}_1$ are two perpendicular spaces defined by kernel functions $K_0(x_i,x_j)$ and $K_1(x_i,x_j)$, respectively. This Sobolev-Hilbert space has an attractive feature. That is, in the presence of boundary condition: $f^{(l)}(0)=0$ for all $l\leq q-1$, any function $f\in\mathcal{W}$ admits the mixed-effects representation:

$$\boldsymbol{f}=\mathbf{A}\boldsymbol{\alpha}+\mathbf{K}_1\boldsymbol{\theta}, \tag{S25}$$

where $\mathbf{A}=(\mathbf{1},\boldsymbol{x},\ldots,\boldsymbol{x}^{q-1}/(q-1)!)$, $\mathbf{K}_1$ is a $(n\times n)$ matrix with the $(i,j)$th entry $K_1(x_i,x_j)$, $\boldsymbol{\alpha}=(\alpha_0,\ldots,\alpha_{q-1})^\top$ is a fixed vector, and $\boldsymbol{\theta}=(\theta_1,\ldots,\theta_n)^\top$ is a random vector that has prior distribution $\mathcal{N}(\mathbf{0},\phi\mathbf{K}_1^+)$ with $\phi>0$. The boundary condition $\mathbf{A}^\top\boldsymbol{\theta}=\mathbf{0}$ means $\boldsymbol{\theta}$ must belong to $\overline{\mathrm{span}}(\mathbf{A})$, enabling us to reparametrize $\boldsymbol{\theta}=\mathbf{Q}\boldsymbol{\pi}$ with arbitrary orthogonal matrix $(n\times(n-q))$ $\mathbf{Q}$ whose columns are the basis of $\overline{\mathrm{span}}(\mathbf{A})$. Thereby, we represent $\boldsymbol{f}=\mathbf{K}\tilde{\boldsymbol{\theta}}$ where $\mathbf{K}=(\mathbf{A}:\mathbf{K}_1\mathbf{Q})$ and $\tilde{\boldsymbol{\theta}}=(\boldsymbol{\alpha}^\top,\boldsymbol{\pi}^\top)^\top$. The "prior distribution" of $\tilde{\boldsymbol{\theta}}$ is

$$\tilde{\boldsymbol{\theta}}\sim\mathcal{N}(\mathbf{0},\phi\mathrm{diag}(\mathbf{0},\mathbf{Q}^\top\mathbf{K}_1\mathbf{Q})^+) \tag{S26}$$

Wahba and Wendelberger [12] introduced how to conduct low-rank smoothing spline. Suppose $\{\kappa_1,\ldots,\kappa_p\}\subset\{x_1,\ldots,x_n\}$ are $p$ knots. The low-rank smoothing spline has the same representation $\boldsymbol{f}=\mathbf{A}\boldsymbol{\alpha}+\mathbf{K}_1\boldsymbol{\theta}$ where the $(i,j)$th entry of $\mathbf{K}_1$ is $K_1(x_i,\kappa_j)$. The "prior distribution" of $\boldsymbol{\theta}$ is $\mathcal{N}(\mathbf{0},\boldsymbol{\Omega})$ where the $(i,j)$th entry of $\boldsymbol{\Omega}$

is $K_1(\kappa_i, \kappa_j)$, and the boundary condition now is $\tilde{\mathbf{A}}^\top \boldsymbol{\theta} = \mathbf{0}$ where $\tilde{\mathbf{A}} = (\mathbf{1}, \boldsymbol{\kappa}, \ldots, \boldsymbol{\kappa}^{q-1}/(q-1)!)$ is a $(p \times 1)$ matrix. Using a similar scheme, we can obtain the final represnetation $\boldsymbol{f} = \mathbf{K}\tilde{\boldsymbol{\theta}}$ where $\mathbf{K} = (\mathbf{A} : \mathbf{K}_1\mathbf{Q})$ where $\tilde{\boldsymbol{\theta}} \sim \mathcal{N}(\mathbf{0}, \phi \text{diag}(\mathbf{0}, \mathbf{Q}^\top \boldsymbol{\Omega}_1 \mathbf{Q})^+)$.

In our study, we found the following three cubic spline basis functions to be equivalent:

- Wahba cubic spline basis function: $K_1(x|\kappa) = \int_0^{\min(x,\kappa)} (x-u)(\kappa-u)\mathrm{d}u$;
- Thin-plate cubic spline basis function: $K_1(x|\kappa) = |x-\kappa|^3$;
- Truncated cubic spline basis function: $K_1(x|\kappa) = (x-\kappa)_+^3$.

Later we will give numerical evidence that the shape of the orthogonal matrix $\mathbf{B}$ constructed based on the three cubi spline functions is the same. For higher polynomial splines, the basis functions of the three splines have the following representation:

- Wahba $(2q+1)$-polynomial spline basis functions: $K_1(x|\kappa) = \int_0^{\min(x,\kappa)} (x-u)^q(\kappa-u)^q\mathrm{d}u$;
- Thin-plate $(2q+1)$-polynomial spline basis function: $K_1(x|\kappa) = |x-\kappa|^{2q+1}$;
- Truncate $(2q+1)$-polynomial spline basis functions: $K_1(x|\kappa) = (x-\kappa)_+^{2q+1}$.

###### 1.7.3 Patterns of Kernel-Based and Smoothing-Spline-Based Basis Functions

We first investigate the patterns of columns of $\mathbf{B}$. Figure S2-S3 display the patterns of columns of $\mathbf{B}$ generated by Mat'ern family kernel function and Wahba cubic kernel function. It is obvious that all the kernels share very similar patterns of columns of $\mathbf{B}$, no matter the fixed effects are null, intercept, linearity, quadratic polynomial, piecewise linearity with changepoint $\nu = 0.5$, linearity discontinuity with breakpoint $\nu_0 = 0.5$. Besides, when fixed effect is piecewise linearity, the patterns of columns of $\mathbf{B}$ are also singular at the changepoint. Similarly, when "fixed effects" is linearity discontinuity, the patterns are discontinuous at the breakpoint.

We then investigate the patterns of columns of $\mathbf{B}$ generated by Mat'ern family kernel with different scale parameter $\rho$, and the patterns of columns of $\mathbf{B}$ generated by different representations of the smoothing spline. Figure S4 show the results of the Mat'ern family kernel: the patterns of columns of $\mathbf{B}$ are unchanged if $\rho$ is larger than a certain value, and further increasing $\rho$ only makes the value of $K(\cdot,\cdot)$ more close to 1. On the other hand, although the three representation of the cubic smoothing splines are different, the final matrice $\mathbf{B}$ are exactly the same. It confirms that the different representation of the cubic smoothing splines are equivalent.

##### 1.8 Translation from GAM to GLMM

We describe how to perform the hypothesis test (S6) in GAM where there are multiple functions. Recall the GAM in the form of (S1):

$$\boldsymbol{y} \sim \mathbf{EF}(\boldsymbol{\mu}, \phi_0), \quad \boldsymbol{\eta} = \mathbf{Z}\boldsymbol{\gamma} + \boldsymbol{f}_1 + \cdots + \boldsymbol{f}_J,$$

where $\boldsymbol{y}, \boldsymbol{\mu}, \boldsymbol{\eta}, \{\boldsymbol{f}_j\}$ are the vectors of $\{y_i\}, \{\mu_i\}, \{\eta_i\}, \{f_j(x_{ij})\}$. The dispersion parameter $\phi_0$ is assumed to be known in this paper, which can be estimated by using the quasi-likelihood formula [14] in practice.

With the proposed mixed-effects representation, we turn the nonparametric GAM into a parametric GLMM. Specifically, we decompose each smooth function into $\boldsymbol{f}_j = \mathbf{A}_j\boldsymbol{\alpha}_j + \mathbf{F}_j\mathbf{Q}_j\boldsymbol{\pi}_j$ where $\mathbf{A}_j$ is the $(n \times q_j)$ basis matrix of the given parametric structure, $\mathbf{K}_j$ is the $(n \times p_j)$ kernel matrix, and $\mathbf{F}_j = (\mathbf{A}_j, \mathbf{K}_j)$. Here $p_j$ is the number of employed knots for $\boldsymbol{f}_j$. For the purpose of identifiability, all $\mathbf{A}_j$ shall exclude the intercept $\mathbf{1}$ and $\mathbf{Q}_j$ is chosen to satisfy $(\mathbf{1}, \mathbf{A}_j)^\top(\mathbf{1}, \mathbf{F}_j)\mathbf{Q}_j = \mathbf{0}$. If we do not have prior knowledge about the potential structure of $\boldsymbol{f}_j$, we suggest considering the simplest linear structure so that setting $\mathbf{A}_j = \boldsymbol{x}_j$. The SVD is further applied to reduce the dimension of $\mathbf{F}_j\mathbf{Q}_j$ in which the first $k_j$ left singular vectors are employed to form $\mathbf{B}_j$. The final representation is $\boldsymbol{f}_j \approx \mathbf{A}_j\boldsymbol{\alpha}_j + \mathbf{B}_j\boldsymbol{\beta}_j$. The prior distribution of $\boldsymbol{\beta}_j$ is $\mathcal{N}(\mathbf{0}_{k_j \times 1}, \phi_j\boldsymbol{\Theta}_j^+)$ where the $(k_j \times k_j)$ prior covariance matrix $\boldsymbol{\Theta}_j$ has the same form of (S22). Thus, the hypothesis test on whether $\boldsymbol{f}_j$ is characterized by the fixed parametric effect $\mathbf{A}_j\boldsymbol{\alpha}_j$ reduces to the hypothesis test on whether the variance component $\phi_j$ is zero.

Asymptotically, a GLMM is equivalent to a linear model with correlated and Gaussian distributed error [9]. Let $\boldsymbol{Y} = (Y_1, \ldots, Y_n)^\top$ with $Y_i = \eta_i + g'(\mu_i)(y_i - \mu_i)$, which is called the pseudo response. The linear model equivalent to the GLMM is

$$\boldsymbol{Y} = \mathbf{A}\boldsymbol{\alpha} + \mathbf{B}\boldsymbol{\beta} + \boldsymbol{\epsilon}, \tag{S27}$$

where $\mathbf{A} = (\mathbf{A}_1, \ldots, \mathbf{A}_J, \mathbf{Z})$, $\mathbf{B} = (\mathbf{B}_1, \ldots, \mathbf{B}_J)$, $\boldsymbol{\alpha} = (\boldsymbol{\alpha}_1^\top, \ldots, \boldsymbol{\alpha}_J^\top, \boldsymbol{\gamma}^\top)^\top$, $\boldsymbol{\beta} = (\boldsymbol{\beta}_1^\top, \ldots, \boldsymbol{\beta}_J)^\top$. The noise term $\boldsymbol{\epsilon} \sim \mathcal{N}(\mathbf{0}_{n\times 1}, \phi_0 \mathbf{W}^{-1})$, $\mathbf{W} = \operatorname{diag}(W_1, \ldots, W_n)$ with $W_i = 1/(V(\mu_i)g'(\mu_i)^2)$, and $V(\mu)$ is the variance function. Since both $\mathbf{B}\boldsymbol{\beta}$ and $\boldsymbol{\epsilon}$ are random, it is reasonable to consider them as a new noise $\boldsymbol{\varepsilon} = \mathbf{B}\boldsymbol{\beta} + \boldsymbol{\epsilon}$, whose prior distribution is $\mathcal{N}(\mathbf{0}_{n\times 1}, \mathbf{V}(\boldsymbol{\phi}))$ with $\boldsymbol{\phi} = (\phi_1, \ldots, \phi_J)^\top$ and

$$\mathbf{V}(\boldsymbol{\phi}) = \phi_0 \mathbf{W}^{-1} + \sum_{j=1}^{J} \phi_j \mathbf{B}_j \boldsymbol{\Theta}_j^{+} \mathbf{B}_j^\top. \tag{S28}$$

Consequently, the REML function

$$\mathcal{L}_{\text{REML}}(\boldsymbol{\phi}|\widehat{\boldsymbol{\alpha}}) = \mathcal{L}_{\text{ML}}(\boldsymbol{\phi}) - \frac{1}{2}\log\det\{\mathbf{A}^\top \mathbf{V}(\boldsymbol{\phi})^{-1}\mathbf{A}\}, \tag{S29}$$

where the negative log-likelihood function of $\mathcal{L}_{\text{ML}}(\boldsymbol{\phi})$ is given by

$$\mathcal{L}_{\text{ML}}(\boldsymbol{\phi}) = -\frac{1}{2}(\boldsymbol{Y} - \mathbf{A}\boldsymbol{\alpha})^\top \mathbf{V}(\boldsymbol{\phi})^{-1}(\boldsymbol{Y} - \mathbf{A}\boldsymbol{\alpha}) - \frac{1}{2}\log\det(\mathbf{V}(\boldsymbol{\phi})). \tag{S30}$$

#### 1.9 Estimation of GAM

Recall that we represent $\boldsymbol{f}_j \approx \mathbf{A}_j\boldsymbol{\alpha}_j + \mathbf{B}_j\boldsymbol{\beta}_j$ using the mixed-effects formulation described in Section 1.6 in this supplementary materials. Let $\mathbf{A} = (\mathbf{A}_1, \ldots, \mathbf{A}_J, \mathbf{Z})$, $\mathbf{B} = (\mathbf{B}_1, \ldots, \mathbf{B}_J)$, $\boldsymbol{\alpha} = (\boldsymbol{\alpha}_1^\top, \ldots, \boldsymbol{\alpha}_J^\top, \boldsymbol{\gamma}^\top)^\top$, and $\boldsymbol{\beta} = (\boldsymbol{\beta}_1^\top, \ldots, \boldsymbol{\beta}_J)^\top$. We estimate $\boldsymbol{\tau} = (\boldsymbol{\alpha}^\top, \boldsymbol{\beta}^\top)^\top$ by minimizing the penalized likelihood

$$\widehat{\boldsymbol{\tau}} = \arg\min_{\boldsymbol{\tau}} \left\{ \mathcal{L}_{\text{ML}}(\boldsymbol{\tau} \mid \boldsymbol{y}) + \sum_{j=1}^{J} \frac{1}{2\phi_j} \boldsymbol{\beta}_j^\top \boldsymbol{\Theta}_j \boldsymbol{\beta}_j \right\}, \tag{S31}$$

where $\boldsymbol{\Theta}_j$ is the penalty matrix associated with the smooth term $f_j(\cdot)$ and $\phi_j$ is its smoothing parameter. The optimization is carried out using iterative reweighted least squares (IRLS). TAPS is implemented in the `mgcv.taps` R package, which is fully integrated with `mgcv` and inherits its automatic selection of smoothing parameters and model fitting routines.

#### 1.10 Score Test of Variance Component

Without loss of generality, we consider $f_1$ as the target function to be tested. The corresponding hypothesis test now is

$$\text{H}_0 : \phi_1 = 0 \qquad \text{v.s.} \qquad \text{H}_1 : \phi_1 > 0. \tag{S32}$$

Besides, since we focus on the hypothesis test in this paper, we suppose that we have obtained the consistent estimators $\widehat{\boldsymbol{\alpha}}$ and $\widehat{\boldsymbol{\beta}}$ and have selected the variance component vector $\widehat{\boldsymbol{\phi}} = (\hat{\phi}_1, \ldots, \hat{\phi}_J)^\top$. The pseudo response is then given by $\widehat{\boldsymbol{Y}} = \widehat{\boldsymbol{\eta}} + (\mathbf{y} - \widehat{\boldsymbol{\mu}})/V(\widehat{\boldsymbol{\mu}})$ where $\widehat{\boldsymbol{\eta}} = \mathbf{A}\widehat{\boldsymbol{\alpha}} + \mathbf{B}\widehat{\boldsymbol{\beta}}$ and $\widehat{\boldsymbol{\mu}} = g^{-1}(\widehat{\boldsymbol{\eta}})$.

The $p$-value of (S32) is calculated using the score test, which was originally developed for testing the polynomial structure in a univariate nonparametric model [19]. The score test is based on the score equation $\mathbf{U}(\cdot)$ of the REML function below:

$$\mathbf{U}(\phi_j) = \left( \underbrace{\frac{1}{2}\operatorname{tr}(\mathbf{P}(\boldsymbol{\phi})^{\frac{1}{2}}\mathbf{G}_j\mathbf{P}(\boldsymbol{\phi})^{\frac{1}{2}})}_{\mathbf{e}(\phi_j)} + \underbrace{\frac{1}{2}\boldsymbol{Y}^\top \mathbf{P}(\boldsymbol{\phi})^{-1}\mathbf{G}_j\mathbf{P}(\boldsymbol{\phi})^{-1}\boldsymbol{Y}\}}_{\mathbf{u}(\phi_j)} \right)\Bigg|_{\boldsymbol{\phi}=\widehat{\boldsymbol{\phi}}_0^j}, \tag{S33}$$

where $\hat{\boldsymbol{\phi}}_0^j = (\hat{\phi}_1, \ldots, \hat{\phi}_{j-1}, 0, \hat{\phi}_{j+1}, \ldots, \hat{\phi}_J)^\top$, $\mathbf{G}_j = \partial \mathbf{V}(\boldsymbol{\phi})/\partial \phi_j = \mathbf{F}_j \mathbf{Q}_j \boldsymbol{\Omega}_j \mathbf{Q}_j^\top \mathbf{F}_j^\top$, and

$$\mathbf{P}(\boldsymbol{\phi}) = \mathbf{V}(\boldsymbol{\phi})^{-1} - \mathbf{V}(\boldsymbol{\phi})^{-1}\mathbf{A}(\mathbf{A}^\top \mathbf{V}(\boldsymbol{\phi})^{-1}\mathbf{A})^{-1}\mathbf{A}^\top \mathbf{V}(\boldsymbol{\phi})^{-1}. \tag{S34}$$

Wu et al. [17] showed that under the null hypothesis, $\mathbf{u}(\phi_1)$ in (S33) asymptotically follows a weighted sum of independent chi-squared random variables:

$$\mathbf{u}(\phi_1) \sim \sum_{j=1}^{q_1} \lambda_j \chi_{1j}^2, \tag{S35}$$

where $q_1$ is the rank of the matrix $\mathbf{S}_1$, $\chi_{1j}^2$ denotes a chi-squared random variable with 1 degree of freedom, and $\lambda_1, \ldots, \lambda_{q_1}$ are the eigenvalues of the matrix $\mathbf{P}(\hat{\boldsymbol{\phi}}_0^1)^{\frac{1}{2}}\mathbf{G}_1\mathbf{P}(\hat{\boldsymbol{\phi}}_0^1)^{\frac{1}{2}}$. Following sequence kernel association test (SKAT) and the related methods [17], we use numerical approximations to calculate the $p$-value regarding the null distribution (S35).

When the sample size $n$ is large, direct construction of the $(n \times n)$ precision matrix $\mathbf{V}(\boldsymbol{\phi})^{-1}$ and the $(n \times n)$ projection matrix $\mathbf{P}(\boldsymbol{\phi})$ is computationally unfeasible. To address this, we develop an operator-based strategy to compute the score statistic $u(\phi_1)$, its expectation $e(\phi_1)$, and its variance component $\mathbf{H}(\phi_1, \phi_1)$, without explicitly forming these matrices. In particular, we construct matrix–vector product operators that evaluate the action of $\mathbf{V}(\boldsymbol{\phi})^{-1}$ and $\mathbf{P}(\boldsymbol{\phi})$ on arbitrary vectors, and use them to implement both residual projection and trace computation efficiently. For the trace terms, we exploit the low-rank structure of the form $\mathbf{G}_1 = \mathbf{F}_1\mathbf{Q}_1\mathbf{S}_1\mathbf{Q}_1^\top\mathbf{F}_1^\top$. These computational tricks make the score test scalable and memory-efficient for large datasets.

It is worth noting that, while we use the score test proposed by Zhang and Lin [19], we do not analyze the identical problem they did. The hypothesis test on polynomial structure in the univariate nonparametric model with ordinal random effects was explored by them, whereas the hypothesis test on arbitrary structure in GAM was studied by us. In our opinion, what Zhang and Lin [19] provided is a technical tool that can be used in the GLMM to perform the variance component test, which has been incorporated into TAPS. TAPS is unique in that it shows how to construct a mixed-effects representation where the fixed parametric effect can follow an arbitrary parametric structure, as well as how to utilize that representation to convert a GAM into a testable GLMM.

#### 1.11 Wald Test of Smoothing Parameters

We directly adopt the Wald test procedure proposed by Wood [15] to evaluate whether the smooth deviation term $b(x)$ is equal to zero. This test is constructed under the framework of penalized regression splines in a GAM, where smooth terms are treated as random effects with fixed smoothing parameters. Let $\hat{\boldsymbol{\pi}}$ denote the estimate of the smooth coefficient vector $\boldsymbol{\pi}$ under the penalized likelihood. The null hypothesis is $\mathrm{H}_0 : b(x) = 0$, that is, the deviation from the specified parametric structure vanishes.

The Wald test statistic is computed as

$$T_{b_1} = \hat{\boldsymbol{b}}_1^\top \mathbf{S}_{b_1}^{+} \hat{\boldsymbol{b}}_1, \tag{S36}$$

where $\hat{\boldsymbol{b}}_1 = \mathbf{B}_1\hat{\boldsymbol{\beta}}_1$ is the fitted smooth deviation term, and

$$\mathbf{S}_{b_1} = \hat{\phi}_1 \mathbf{B}_1 \boldsymbol{\Theta}_1^{+} \mathbf{B}_1^\top$$

is its corresponding covariance matrix under the estimated smoothing variance $\hat{\phi}_1$. This test statistic $T_{b_1}$ has the intuitive form of a Mahalanobis norm and may follow a $\chi^2$ distribution under the null. The construction of $T_b$ can be further simplified. Let $\mathbf{B}_1 = \mathbf{Q}_1\mathbf{R}_1$ be the QR decomposition of $\mathbf{B}_1$. Then the test statistic becomes

$$\begin{aligned} T_{b_1} &= \hat{\phi}_1^{-1}\hat{\boldsymbol{\beta}}_1^\top \mathbf{B}_1^\top \left(\mathbf{B}_1\boldsymbol{\Theta}_1^{+}\mathbf{B}_1^\top\right)^{+} \mathbf{B}_1\hat{\boldsymbol{\beta}}_1 \\ &= \hat{\phi}_1^{-1}\hat{\boldsymbol{\beta}}_1^\top \mathbf{R}_1^\top \mathbf{Q}_1^\top \left(\mathbf{Q}_1\mathbf{R}_1\boldsymbol{\Theta}_1^{+}\mathbf{R}_1^\top\mathbf{Q}_1^\top\right)^{+} \mathbf{Q}_1\mathbf{R}_1\hat{\boldsymbol{\beta}}_1 \\ &= \hat{\phi}_1^{-1}\hat{\boldsymbol{\beta}}_1^\top \mathbf{R}_1^\top \left(\mathbf{R}_1\boldsymbol{\Theta}_1^{+}\mathbf{R}_1^\top\right)^{+} \mathbf{R}_1\hat{\boldsymbol{\beta}}_1. \end{aligned} \tag{S37}$$

Thus, the computational bottleneck of the Wald test reduces to the QR decomposition of the design matrix $\mathbf{B}_1$. Once the QR factors are obtained, the remaining steps involve only low-dimensional matrix operations.

However, due to the penalization imposed on $b_1(x)$, which effectively shrinks some of the smooth components, the degrees of freedom associated with $\hat{\boldsymbol{b}}_1$ are generally not integers. Wood [15] showed that $T_{b_1}$ in fact follows a nonstandard distribution, which can be well approximated by a scaled chi-squared distribution using the moment matching method. The key idea is to match the first two moments of $T_{b_1}$ to those of a $\kappa\chi^2_\nu$ distribution, where $\kappa$ is a scaling constant and $\nu$ is a fractional degree of freedom. Wood [15] further modified the basic moment-matching approach to improve accuracy and stability, particularly in the presence of small effective degrees of freedom. Instead of relying solely on raw moments, he proposed a two-component approximation of $T_{b_1}$ as a sum of a chi-squared variable with integer degrees of freedom and a second component approximated using a low-rank spectral decomposition.

### 2 Supplementary Simulation (with codes)

#### 2.1 General Setting

In the simulation study, we consider the following GAM:

$$g(\mathrm{E}(y_i)) = \eta_i = \alpha_0 + f_1(x_{i1}) + f_2(x_{i2}) + f_3(x_{i3}) + f_4(x_{i4}), \tag{S38}$$

where $y_i$ is the response variable following a Gaussian, Poisson, or binary distribution; $\alpha_0$ is an intercept; and $f_1$ is the target function to be tested. Here, $f_2$ is set to zero,

$$f_3(x_3) = 0.4\sin(t_3) + 0.8\cos(t_3) + 1.2\sin(t_3)^2 + 1.6\cos(t_3)^3 + 2\sin(t_3)^3, \tag{S39}$$

where $t_3 = 2\pi x_3$ and

$$f_4(x_4) = 3\sin(3t_4) + 6\exp(-36t_4^2), \tag{S40}$$

where $t_4 = 2(x_4 - 0.5)$. We evaluate four structural scenarios for $f_1$:

- Linearity: $f_1(x_1) = \alpha_1 x_1$, representing a simple linear structure.
- Piecewise Linearity: $f_1(x_1) = \alpha_1 x_1 + \alpha_2(x_1 - 0.5)\mathrm{I}(x_1 > 0.5)$, a piecewise linear function with a change of slope at $\nu_1 = 0.5$.
- Linearity Discontinuity: $f_1(x_1) = \alpha_1\mathrm{I}(x_1 > 0.5) + \alpha_2(x_1 - 0.5) + \alpha_3(x_1 - 0.5)\mathrm{I}(x_1 > 0.5)$, a discontinuous linear structure with a jump at $\nu_0 = 0.5$.
- Linear Interaction: $f_{12}(x_1, x_2) = \alpha_1 x_1 + \alpha_2 x_2 + \alpha_3 x_1 x_2$. TAPS can test whether a multivariate function follows a specific parametric structure like linear interaction.

The functions $f_3$ and $f_4$ remain fixed, while $f_2$ is set to zero in the first three settings.

Besides, we use the Gaussian-Beta copula method to generate correlated and Gaussian-like covariates on interval $[0, 1]$: generate $\mathbf{z}_i = (z_{i1}, z_{i2}, z_{i3}, z_{i4})^\top \sim \mathcal{N}(\mathbf{0}_{4\times 1}, \mathbf{cs}(0.25))$ and then yield $x_{ij} = F_\beta^{-1}(\Phi(z_{ij}), 1.5, 1.5)$ where $\mathbf{cs}(\rho) = (1-\rho)\mathbf{I}_4 + \rho\mathbf{1}_4\mathbf{1}_4^\top$, $\Phi(\cdot)$ is the CDF of standard normal distribution and $F_\beta(\cdot, a, b)$ is the CDF of beta distribution with parameters $a$ and $b$. The density function of the beta distribution resembles that of the Gaussian distribution when $a$ and $b$ are equal and larger than 1. The knots $\{\kappa_{j1}, \ldots, \kappa_{jp}\}$ are chosen as the $1/p, \ldots, p/p$ quantiles of $\boldsymbol{x}_j$, $j \in \{1, 2, 3, 4\}$ and $p \equiv \min(n/4, 300)$ for all $j$. The Mat{'e}rn family kernel [10]

$$K(x_i, x_j) = \exp\left(-\frac{|x_i - x_j|}{s}\right) \times \left(1 + \frac{|x_i - x_j|}{s}\right)$$

is used to construct the design matrix of the random effect, with order $\nu = 1.5$ and scale parameter $s = 10$.

The pattern of $f_3$ and $f_4$ are as follows:

```
library(MASS)
library(dplyr)
library(ggplot2)
library(egg)
n=1000
X=mvrnorm(n,rep(0,4),matrix(0.25,4,4)+0.75*diag(4))
x1=qbeta(pnorm(X[,1]),1.5,1.5)
x2=qbeta(pnorm(X[,2]),1.5,1.5)
x3=qbeta(pnorm(X[,3]),1.5,1.5)
x4=qbeta(pnorm(X[,4]),1.5,1.5)
t3=2*pi*x3
f3=0.4*sin(t3)+0.8*cos(t3)+1.2*sin(t3)^2+1.6*cos(t3)^3+2*sin(t3)^3
t4=2*(x4-0.5)
f4=4*sin(3*t4)+6*exp(-36*t4^2)
```

```r
df <- data.frame(x3 = x3, f3 = f3, x4 = x4, f4 = f4)
ggplot(df, aes(x = x3, y = f3)) +
  geom_point(size = 0.7, alpha = 0.6, color = "#1f77b4") +
  labs(x = expression(x[3]), y = expression(f[3](x[3])))
```

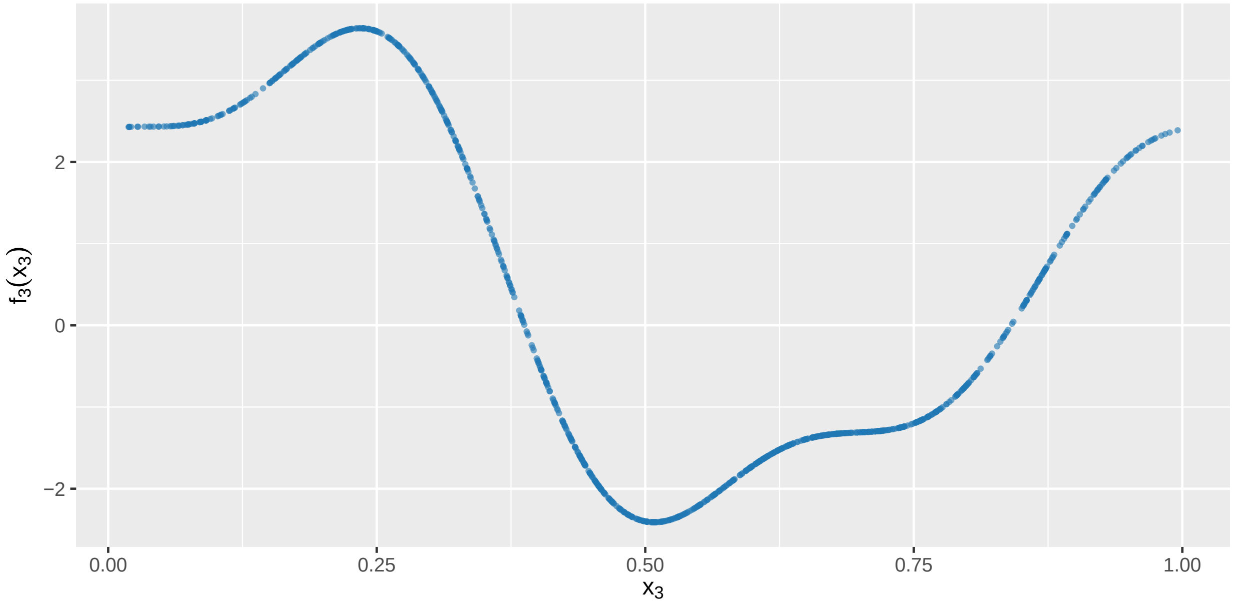

```r
ggplot(df, aes(x = x4, y = f4)) +
  geom_point(size = 0.7, alpha = 0.6, color = "#d62728") +
  labs(x = expression(x[4]), y = expression(f[4](x[4])))
```

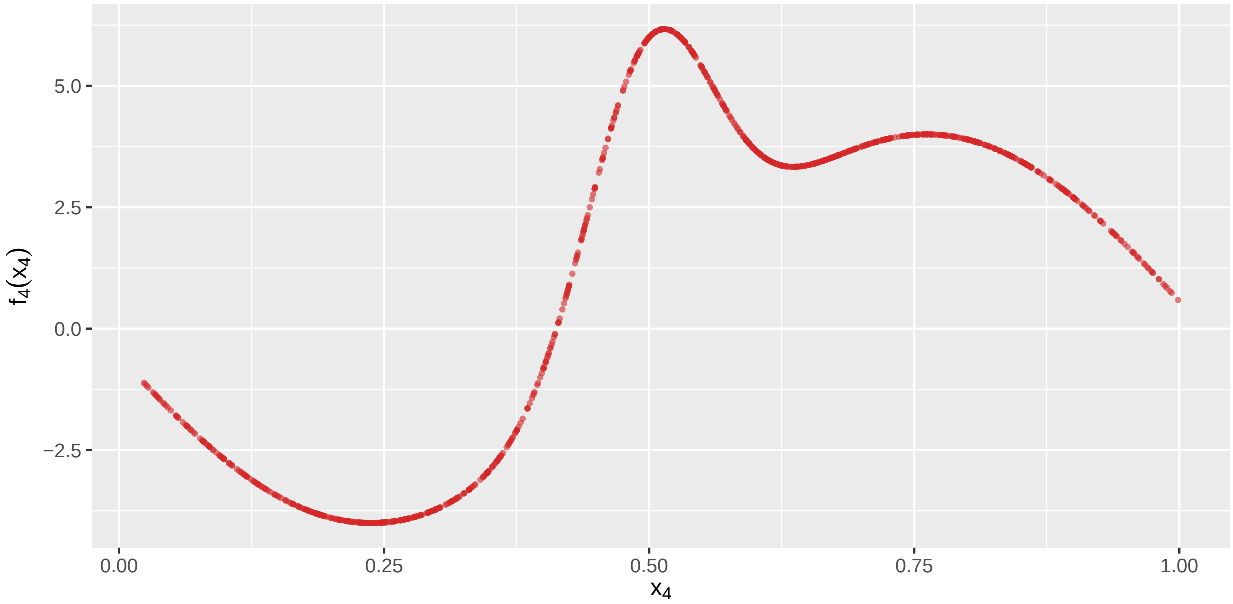

#### 2.2 Setting of Linearity Test

We test whether $f_1(x_1) = \alpha_1 x_1$ with a certain coefficient $\alpha_1$, i.e., $f_1(x_1)$ follows linearity, which may be the simplest and also the most important case. In the GAM (S38), we consider the following target function:

$$f_1(x_1) = 4x_1 + a\exp(-16(x - 0.5)^2),$$

where $a$ is a scale size. A larger $a$ will make $f_1(x_1)$ more different from a linear function. We set $a = 0, 1, \ldots, 5$ and the null hypothesis holds when $a = 0$. For convenience, we introduce a so-called deviation parameter $d$, which corresponds to $a$ via the relationship:

$$a = d.$$

The patterns of $f_1(x_1)$ with different $d$ is as follows:

```
a_vals <- 0:4

df_f1 <- lapply(a_vals, function(a) {
  data.frame(
    x1 = x1,
    f1 = 4 * x1 + a * exp(-16 * (x1 - 0.5)^2),
    d = a
  )
}) %>% bind_rows()

df_f1$d=ordered(df_f1$d,levels=as.character(c(0:4)))

ggplot(df_f1, aes(x = x1, y = f1, color = d)) +
  geom_line(size = 1) +
  scale_color_brewer(palette = "RdYlBu") +
  labs(x = expression(x[1]), y = expression(f[1](x[1])),
       title = expression(paste("Deviation from linearity in ", f[1](x[1]),
                                " under varying ", d)))
```

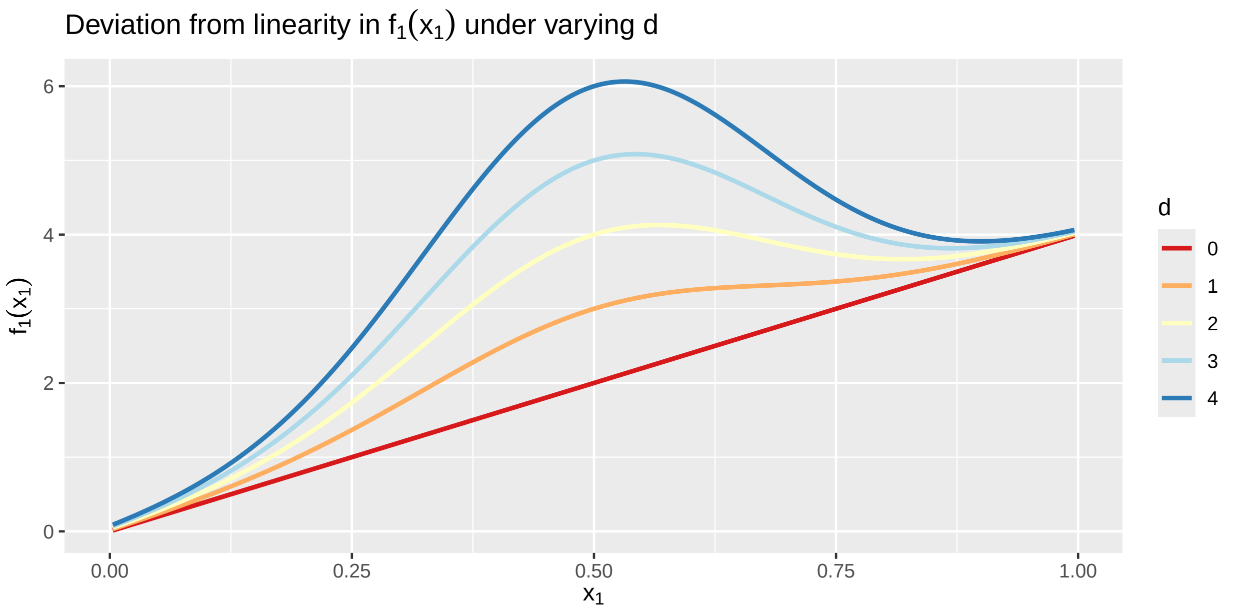

#### 2.3 Settings of Piecewise Linearity Test

We test whether

$$f_1(x_1) = \alpha_1 x_1 + \alpha_2 (x_1 - 0.5)_+$$

with certian coefficients $\alpha_1, \alpha_2$, i.e., $f_1(x_1)$ has a piecewise linear structure with two changepoints $\nu_1 = 0.3, \nu_2 = 0.7$. The difficulty in this scenario is distinguishing the changepoints in a piece-wise linear function from the changepoints in a general smooth function; e.g., the vertice of a quadratic function can also be seen as a change point, but linearity with change points cannot describe a quadratic function. Motivated by this, we consider the following function:

$$f_1(x_1) = 6x_1 - 18s(x_1, 0.5, a),$$

where

$$s(x, \nu, a) = \left\{\frac{(x-\nu+a)^2}{4a}\right\}\mathrm{I}\{x \in [\nu - a, \nu + a)\} + (x - \nu)\mathrm{I}(x \geq \nu + a).$$

Intuitively, $s(x, \nu, a)$ is the smoothed version of the segmented function $(x - \nu)_+$, which smooths the singular point $x = \nu$ using a quadratic spline in the neighborhood $[\nu - h, \nu + h]$. The smoothing size $a$ is set as $a = 0, 0.1, 0.2, 0.3, 0.4$ and $f_1(x_1)$ is an exact piecewise linear function only when $a = 0$. For convenience, we introduce a so-called deviation parameter $d$, which corresponds to $a$ via the relationship:

$$a = 0.1d.$$

The smoothed piecewise linear function has already been implemented in the mgcv.taps package, and users can directly call it to generate the corresponding basis functions. The patterns of $f_1(x_1)$ under different deviations are shown as follows:

```r
library(mgcv.taps)
library(ggplot2)
library(dplyr)

x1 <- seq(0, 1, length.out = 500)
d_vals <- 0:4
a_vals <- 0.1 * d_vals

df_f1 <- lapply(seq_along(d_vals), function(i) {
  d <- d_vals[i]
  a <- a_vals[i]

  X1_design <- if (a != 0) {
    smoothed_piecewise_linearity(x1, list(0.5, a))[,-1]
  } else {
    piecewise_linearity(x1, 0.5)[,-1]
  }

  f1 <- c(X1_design %*% c(6, -18))
  data.frame(x1 = x1, f1 = f1, d = d)
}) %>% bind_rows()

ggplot(df_f1, aes(x = x1, y = f1, color = factor(d))) +
  geom_line(size = 1) +
  scale_color_brewer(palette = "RdYlBu", name = "d") +
  labs(x = expression(x[1]), y = expression(f[1](x[1])),
       title = expression(paste("Deviation from piecewise linearity in ",
                                f[1](x[1]), " under varying ", d)))
```

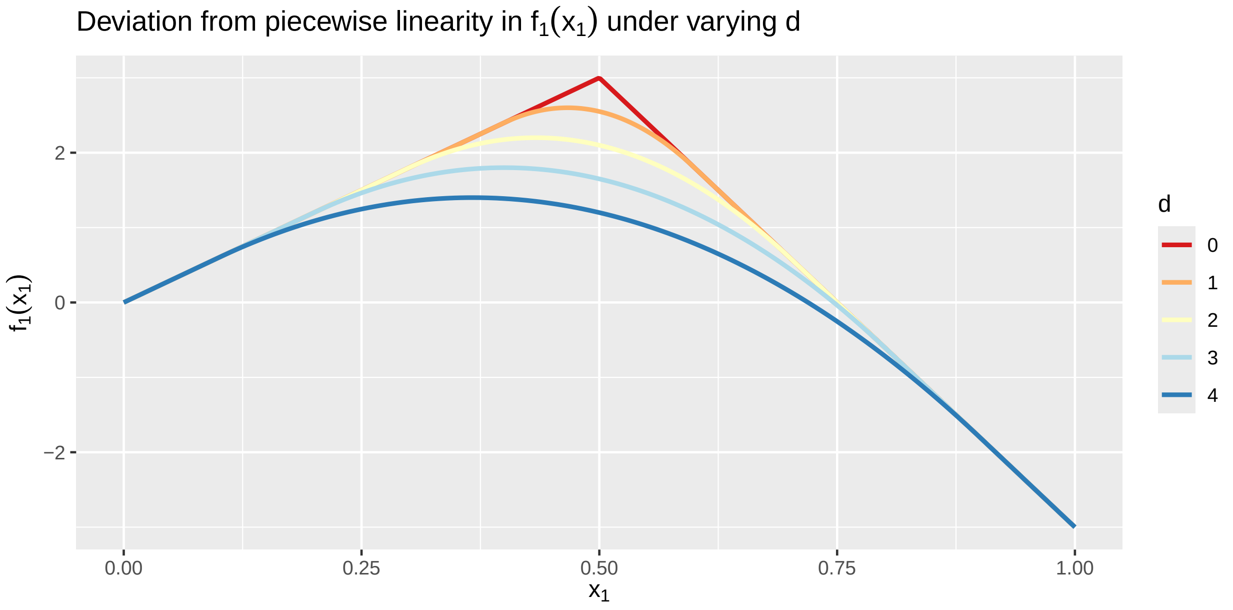

#### 2.4 Settings of Linearity Discontinuity Test

We test whether

$$f_1(x_1) = \alpha_1 \mathrm{I}(x_1 > 0.5) + \alpha_2(x_1 - 0.5) + \alpha_3\{(x - 0.5)\mathrm{I}(x > 0.5)\}$$

with certain coefficients $\alpha_1, \alpha_2, \alpha_3$, which is a linear discontinuous function with a breakpoint $\nu_0 = 0.5$. The difficulty is distinguishing the discontinuity from a very steep but continuous change in a neighborhood of the breakpoint. Hence, we consider the following target function:

$$f_1(x_1) = 4x_1 + \frac{2}{a}(x - 0.5 + a)_+ - \frac{2}{c}(x - 0.5 - a)_+,$$

where the shift size $a = 0, 0.02, \ldots, 0.08$. (The limit of $f_1(x_1)$ when $a \to 0$ is $f_1(x_1) = 4x_1 + 4\mathrm{I}(x > 0.5)$.) Intuitively, this function links the two discontinuous parts of a function in the space $\mathrm{span}(\mathrm{I}(x_1 > 0.5), (x_1 - 0.5), \{(x - 0.5)\mathrm{I}(x > 0.5)\})$ with a linear spline in the neighborhood $[\nu_0 - a, \nu_0 + a]$. For convenience, we introduce a so-called deviation parameter $d$, which corresponds to $a$ via the relationship:

$$a = 0.02d.$$

The smoothed linear discontinuous function has already been implemented in the mgcv.taps package, and users can directly call it to generate the corresponding basis functions. The patterns of $f_1(x_1)$ under different deviations are shown as follows:

```r
library(mgcv.taps)
library(ggplot2)
library(dplyr)

x1 <- seq(0, 1, length.out = 500)
d_vals <- 0:4
a_vals <- 0.02 * d_vals

df_f1 <- lapply(seq_along(d_vals), function(i) {
  d <- d_vals[i]
  a <- a_vals[i]
```

```
  X1_design <- if (a != 0) {
    smoothed_linearity_discontinuity(x1,list(0.5,a))[,-1]
  } else {
linearity_discontinuity(x1,0.5)[,-1]
  }

  f1 <- c(X1_design%*%c(4,4,0))
  data.frame(x1 = x1, f1 = f1, d = d)
}) %>% bind_rows()

ggplot(df_f1, aes(x = x1, y = f1, color = factor(d))) +
  geom_line(size = 1) +
  scale_color_brewer(palette = "RdYlBu", name = "d") +
  labs(x = expression(x[1]), y = expression(f[1](x[1])),
       title = expression(paste("Deviation from linearity discontinuity in ",
                                f[1](x[1]), " under varying ", d)))
```

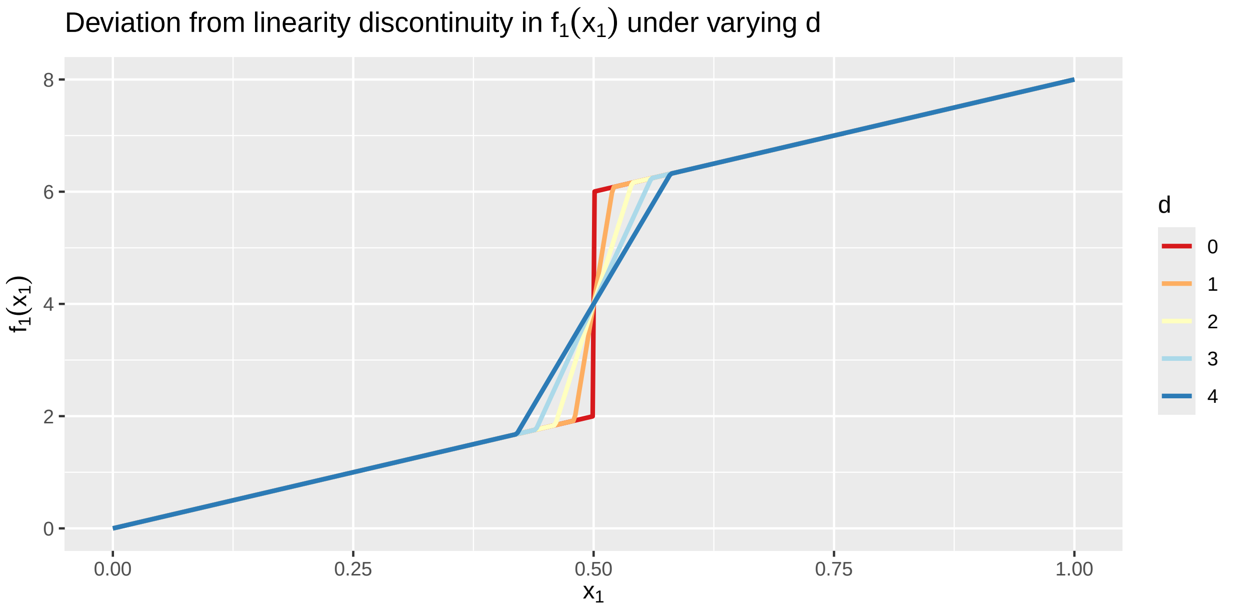

#### 2.5 Settings of Linear Interaction Test

In bivariate regression problems, a key question in epidemiology and social sciences is whether the joint effect of two variables can be adequately captured by a linear interaction model. Specifically, we consider testing whether the target function satisfies the form

$$f_{12}(x_1, x_2) = \alpha_1 x_1 + \alpha_2 x_2 + \alpha_3 x_1 x_2,$$

which defines a linear interaction structure. In many applications, such a parametric form facilitates interpretation and policy evaluation, especially when identifying synergistic effects between exposures.

To evaluate the ability of TAPS to detect deviations from this structure, we design a family of smoothed interaction functions:

$$f_{12}(x_1, x_2) = x_1 + x_2 + x_1 x_2 + a \cdot \exp(-|x_1 - x_2|),$$

where the final term introduces a smooth deviation from the linear interaction. The deviation is controlled by a parameter $a = 0.1 \cdot d$, with $d = 0, 1, 2, 3, 4$. When $d = 0$, the function reduces exactly to a linear interaction model; as $d$ increases, the deviation becomes more pronounced. The exponential term introduces a local effect around the diagonal $x_1 = x_2$, mimicking plausible interaction mechanisms such as threshold-based synergy.

We visualize the function $f_{12}(x_1, x_2)$ over a 2D domain to show how the surface evolves as the deviation parameter increases:

```
grid <- expand.grid(x1 = x1, x2 = x2)

d_vals <- 0:4
a_vals <- 0.1 * d_vals

df_f12 <- lapply(seq_along(d_vals), function(i) {
  d <- d_vals[i]
  a <- a_vals[i]
  with(grid, {
    f12 <- x1 + x2 + x1 * x2 + a * exp(-abs(x1 - x2))
    data.frame(x1 = x1, x2 = x2, f12 = f12, d = d)
  })
}) %>% bind_rows()

ggplot(df_f12, aes(x = x1, y = x2, z = f12)) +
  geom_contour_filled(bins = 20) +
  facet_wrap(~ d, labeller = label_bquote(d == .(d))) +
  scale_fill_viridis_d() +
  guides(fill="none")+
  labs(title = expression(paste("Contour plot of ", f[12](x[1], x[2]),
                                " under varying ", d)),
       x = expression(x[1]), y = expression(x[2]), fill = expression(f[12]))
```

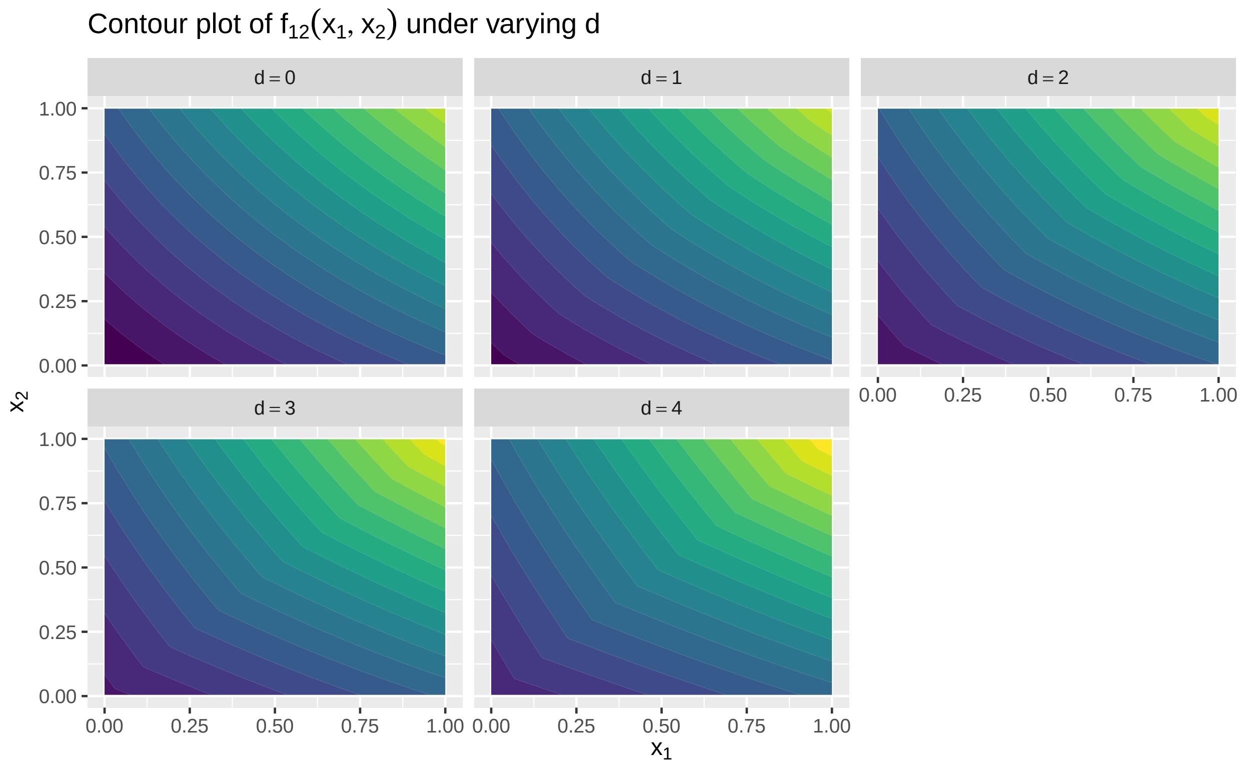

The figure above shows how the function shape transitions from a simple planar surface (linear interaction) at $d = 0$ to increasingly curved and peaked surfaces as $d$ grows.

#### 2.6 Setting of Gaussian Outcome

We use the linearity test to demonstrate how data are generated for different outcomes. First, for a Gaussian outcome, the following code shows one realization of data generation, model fitting, and hypothesis testing:

```
library(mgcv)
X=MASS::mvrnorm(n,rep(0,4),matrix(0.25,4,4)+0.75*diag(4))
x1=qbeta(pnorm(X[,1]),1.5,1.5)
x2=qbeta(pnorm(X[,2]),1.5,1.5)
x3=qbeta(pnorm(X[,3]),1.5,1.5)
x4=qbeta(pnorm(X[,4]),1.5,1.5)
f1=smoothed_linearity(x1,a=0)
t3=2*pi*x3
f3=0.4*sin(t3)+0.8*cos(t3)+1.2*sin(t3)^2+1.6*cos(t3)^3+2*sin(t3)^3
t4=2*(x4-0.5)
f4=3*sin(3*t4)+6*exp(-36*t4^2)
f2=0*x4
eta=f1+f3+f4
y=1+eta+rnorm(n,0,1)*sd(eta/2)
## The guarantee the signal-to-noise ratio is 0.67.
fit=gam(y~s(x1,bs="AMatern",k=10,m=c(100,10))+s(x2,bs="cr",k=10)
        +s(x3,bs="cr",k=10)+s(x4,bs="cr",k=15),method="REML")
summary(fit)
```

```
##
## Family: gaussian
## Link function: identity
##
## Formula:
## y ~ s(x1, bs = "AMatern", k = 10, m = c(100, 10)) + s(x2, bs = "cr",
##     k = 10) + s(x3, bs = "cr", k = 10) + s(x4, bs = "cr", k = 15)
##
## Parametric coefficients:
##             Estimate Std. Error t value Pr(>|t|)
## (Intercept)  2.90006    0.05629   51.52   <2e-16 ***
## ---
## Signif. codes:  0 '***' 0.001 '**' 0.01 '*' 0.05 '.' 0.1 ' ' 1
##
## Approximate significance of smooth terms:
##           edf Ref.df      F  p-value
## s(x1)  1.001  1.002  22.39 2.76e-06 ***
## s(x2)  1.766  2.213   1.95    0.149
## s(x3)  8.347  8.870 155.03  < 2e-16 ***
## s(x4) 12.361 13.472 183.03  < 2e-16 ***
## ---
## Signif. codes:  0 '***' 0.001 '**' 0.01 '*' 0.05 '.' 0.1 ' ' 1
##
## R-sq.(adj) =  0.798   Deviance explained = 80.3%
## -REML = 2036.7  Scale est. = 3.1686    n = 1000
```

```
plot(fit,page=1)
```

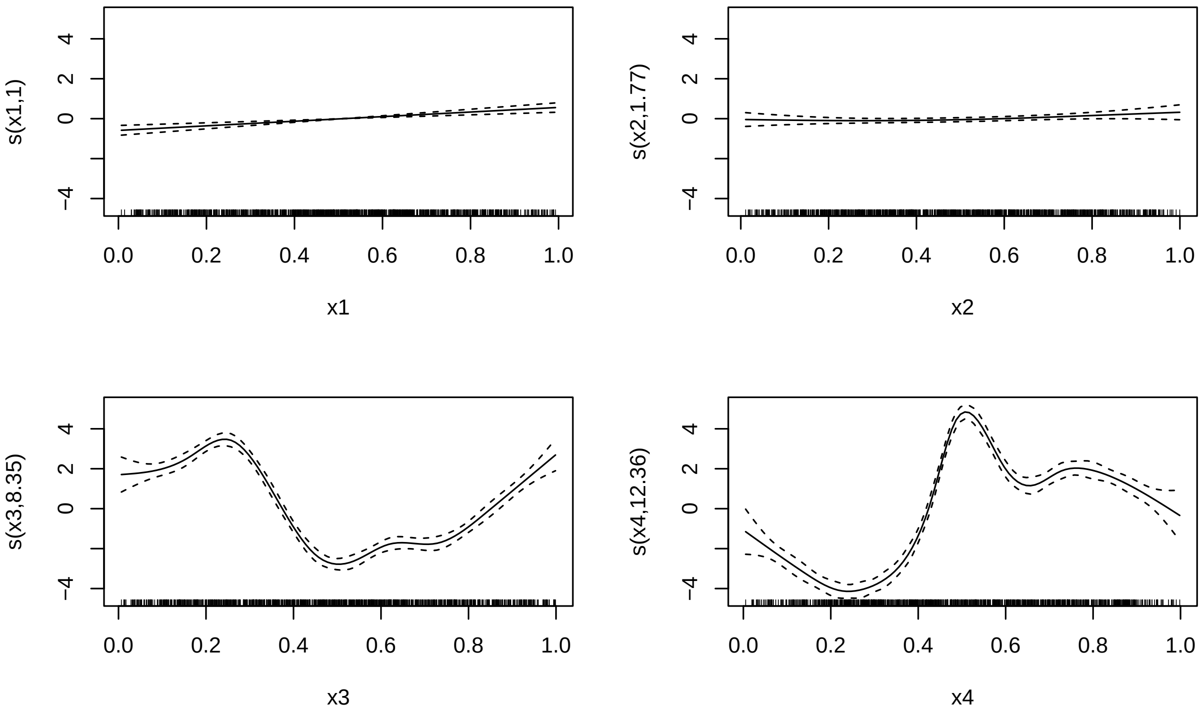

```
taps_wald_test(fit,test.component=1)
```

```
##    mixed.term fix.df fix.chisq   fix.pvalue fix.indices   smooth.df
##        <char>  <num>     <num>        <num>      <char>       <num>
## 1:      s(x1)      1  22.44257 2.165213e-06    reported 0.002070869
##    smooth.chisq smooth.pvalue
##           <num>         <num>
## 1: 7.158558e-06     0.9978659
```

```
taps_score_test(fit,test.component=1)
```

```
##    smooth.term smooth.pvalue method
##         <char>         <num> <char>
## 1:       s(x1)     0.9211583 davies
```

It is important to note that in the above code, `AMatern` refers to the only mixed-effects representation currently supported in mgcv.taps. Here, "A" denotes the fixed-effect component, while "Matern" refers to the Mat'ern kernel:

$$K(x_i, x_j) = \exp\left( - \frac{|x_i - x_j|}{s} \right) \times \left( 1 + \frac{|x_i - x_j|}{s} \right).$$

The argument `k` specifies the dimension of the basis matrix $\mathbf{B}$, formed by the top left singular vectors (i.e., the number of top principal components). The `m` parameter passes two values: the number of knots $p$ and the Mat'ern kernel's scale parameter $s = 10$.

Additionally, `taps_wald_test` and `taps_score_test` are the corresponding Wald and score test functions. The argument `test.component = i` specifies which smooth term in the `mgcv` fit is being tested for degeneracy into a fixed effect. It is important to note that in `taps_score_test`, the reported degree of freedom (smooth.df) corresponds to the parameter $\nu$ in the scaled $\kappa\chi^2_\nu$ test, and not to the effective degrees of freedom (edf) typically reported by `mgcv` following Wood's convention. In contrast, `taps_wald_test` reports the standard notion of edf, consistent with the values shown in the `summary.gam` output of `mgcv`.

We also consider the quantile GAM handled by the R package `qgam`. We genrate random outliers in the model:

```
library(qgam)
f1=smoothed_linearity(x1,a=1)
# we add some non-linear effect into f1
eta=f1+f3+f4
y=1+eta+rnorm(n,0,1)*sd(eta/2)
outlier=eta*0
outlier[sample(n,round(0.01*n))]=sign(rnorm(round(0.01*n),0,5))*10
# we generate 1% outliers in the model
y=y+outlier
dat=data.frame(y=y,x1=x1,x2=x2,x3=x3,x4=x4)
fit1=gam(y~s(x1,bs="AMatern",k=10,m=100)+s(x2,bs="cr",k=10)+
           s(x3,bs="cr",k=10)+s(x4,bs="cr",k=15),method="REML")
fit2=qgam(y~s(x1,bs="AMatern",k=10,m=100)+s(x2,bs="cr",k=10)+
            s(x3,bs="cr",k=10)+s(x4,bs="cr",k=15),qu=0.5,data=dat)
```

```
## Estimating learning rate. Each dot corresponds to a loss evaluation.
## qu = 0.5........done
```

```
summary(fit1)
```

```
##
## Family: gaussian
## Link function: identity
##
## Formula:
## y ~ s(x1, bs = "AMatern", k = 10, m = 100) + s(x2, bs = "cr",
##     k = 10) + s(x3, bs = "cr", k = 10) + s(x4, bs = "cr", k = 15)
##
## Parametric coefficients:
##             Estimate Std. Error t value Pr(>|t|)
## (Intercept)  3.42123    0.06466   52.91   <2e-16 ***
## ---
## Signif. codes:  0 '***' 0.001 '**' 0.01 '*' 0.05 '.' 0.1 ' ' 1
##
## Approximate significance of smooth terms:
##          edf Ref.df       F p-value
## s(x1)  4.133  5.086  10.595  <2e-16 ***
## s(x2)  1.002  1.004   2.988  0.0838 .
## s(x3)  8.279  8.843 109.964  <2e-16 ***
## s(x4) 11.974 13.232 150.642  <2e-16 ***
## ---
## Signif. codes:  0 '***' 0.001 '**' 0.01 '*' 0.05 '.' 0.1 ' ' 1
##
## R-sq.(adj) =  0.755   Deviance explained = 76.1%
## -REML = 2175.3  Scale est. = 4.1804    n = 1000
```

```
summary(fit2)
```

```
##
## Family: elf
## Link function: identity
##
## Formula:
```

```
## y ~ s(x1, bs = "AMatern", k = 10, m = 100) + s(x2, bs = "cr",
##     k = 10) + s(x3, bs = "cr", k = 10) + s(x4, bs = "cr", k = 15)
##
## Parametric coefficients:
##             Estimate Std. Error z value Pr(>|z|)
## (Intercept)  3.38541    0.05841   57.96   <2e-16 ***
## ---
## Signif. codes:  0 '***' 0.001 '**' 0.01 '*' 0.05 '.' 0.1 ' ' 1
##
## Approximate significance of smooth terms:
##          edf Ref.df  Chi.sq p-value
## s(x1)  3.990  4.926   53.15  <2e-16 ***
## s(x2)  1.813  2.275    7.59  0.0351 *
## s(x3)  8.327  8.862 1164.94  <2e-16 ***
## s(x4) 12.153 13.348 2416.87  <2e-16 ***
## ---
## Signif. codes:  0 '***' 0.001 '**' 0.01 '*' 0.05 '.' 0.1 ' ' 1
##
## R-sq.(adj) =  0.752   Deviance explained = 66.9%
## -REML = 2126.3  Scale est. = 1         n = 1000
```

```
plot(fit1,page=1)
```

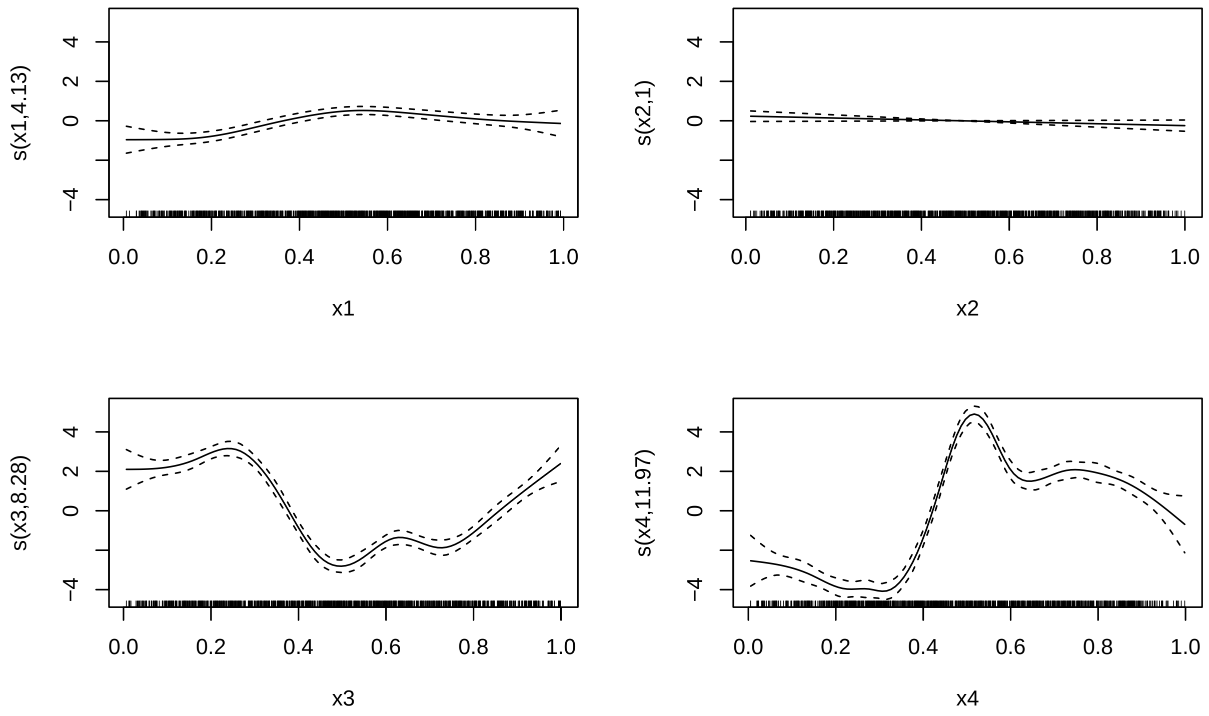

```
plot(fit2,page=1)
```

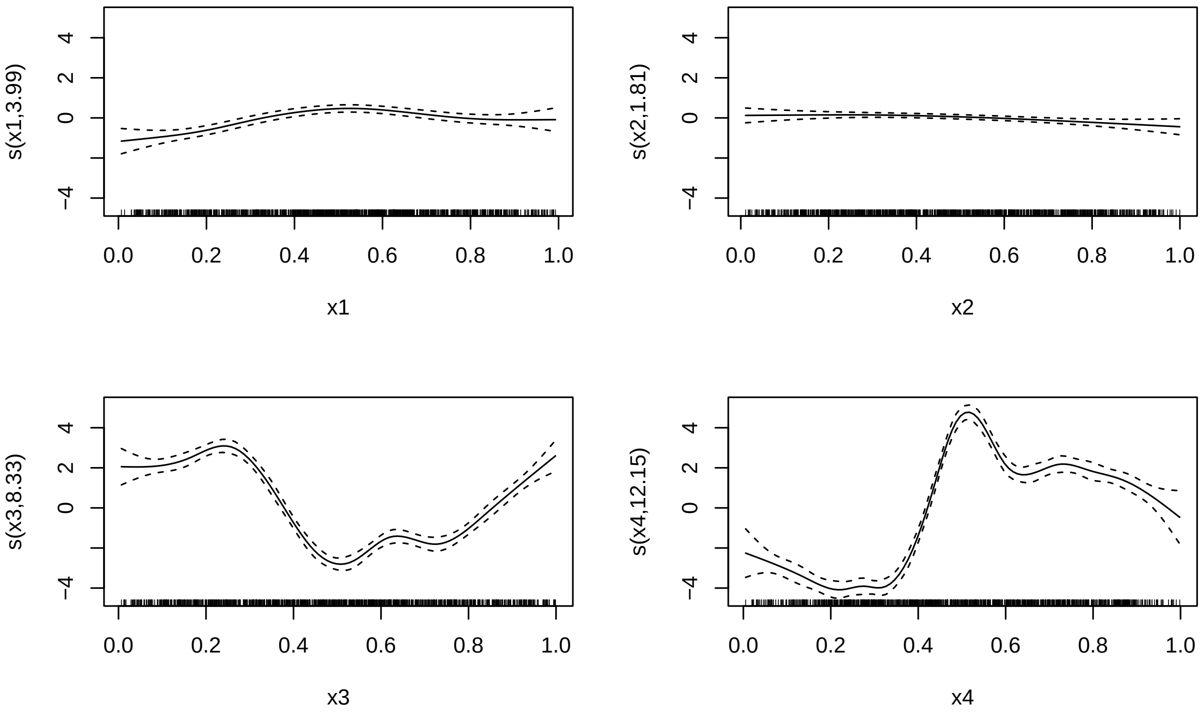

```r
taps_wald_test(fit1,test.component=1)
```

```
##    mixed.term fix.df fix.chisq   fix.pvalue fix.indices smooth.df smooth.chisq
##        <char>  <num>     <num>        <num>      <char>     <num>        <num>
## 1:      s(x1)      1  20.71402 5.332412e-06    reported  4.085722     33.39418
##    smooth.pvalue
##            <num>
## 1:    2.0013e-06
```

```r
taps_wald_test(fit2,test.component=1)
```

```
##    mixed.term fix.df fix.chisq   fix.pvalue fix.indices smooth.df smooth.chisq
##        <char>  <num>     <num>        <num>      <char>     <num>        <num>
## 1:      s(x1)      1  14.30262 0.0001556482    reported  3.925781     39.82405
##    smooth.pvalue
##            <num>
## 1:             0
```

```r
# score test is not available for qgam object
```

#### 2.7 Setting of Binary Outcome

For binary outcome, we first generate the data using the following codes:

```r
f1=smoothed_linearity(x1,4)
#### Here we consider the case where d=4
t3=2*pi*x3
f3=0.4*sin(t3)+0.8*cos(t3)+1.2*sin(t3)^2+1.6*cos(t3)^3+2*sin(t3)^3
t4=2*(x4-0.5)
f4=3*sin(3*t4)+6*exp(-36*t4^2)
f2=0*x4
```

```r
eta=f1+f3+f4
y=rbinom(n,1,exp(eta-2)/(1+exp(eta-2)))
# Here, we adjust the mean of eta such that it is around 0, which generate
# nearly balanced outcome
fit=gam(y~s(x1,bs="AMatern",k=10,m=100)+s(x2,bs="cr",k=10)
        +s(x3,bs="cr",k=10)+s(x4,bs="cr",k=15),method="REML",family=binomial())
summary(fit)
```

```
##
## Family: binomial
## Link function: logit
##
## Formula:
## y ~ s(x1, bs = "AMatern", k = 10, m = 100) + s(x2, bs = "cr",
##     k = 10) + s(x3, bs = "cr", k = 10) + s(x4, bs = "cr", k = 15)
##
## Parametric coefficients:
##             Estimate Std. Error z value Pr(>|z|)
## (Intercept)   2.1249     0.1835   11.58   <2e-16 ***
## ---
## Signif. codes:  0 '***' 0.001 '**' 0.01 '*' 0.05 '.' 0.1 ' ' 1
##
## Approximate significance of smooth terms:
##          edf Ref.df  Chi.sq p-value
## s(x1)  4.822  5.836  87.654  <2e-16 ***
## s(x2)  1.000  1.001   2.041   0.153
## s(x3)  6.988  8.045 145.610  <2e-16 ***
## s(x4) 10.180 11.670 206.625  <2e-16 ***
## ---
## Signif. codes:  0 '***' 0.001 '**' 0.01 '*' 0.05 '.' 0.1 ' ' 1
##
## R-sq.(adj) =  0.656   Deviance explained = 62.4%
## -REML = 276.05  Scale est. = 1         n = 1000
```

```r
plot(fit,page=1)
```

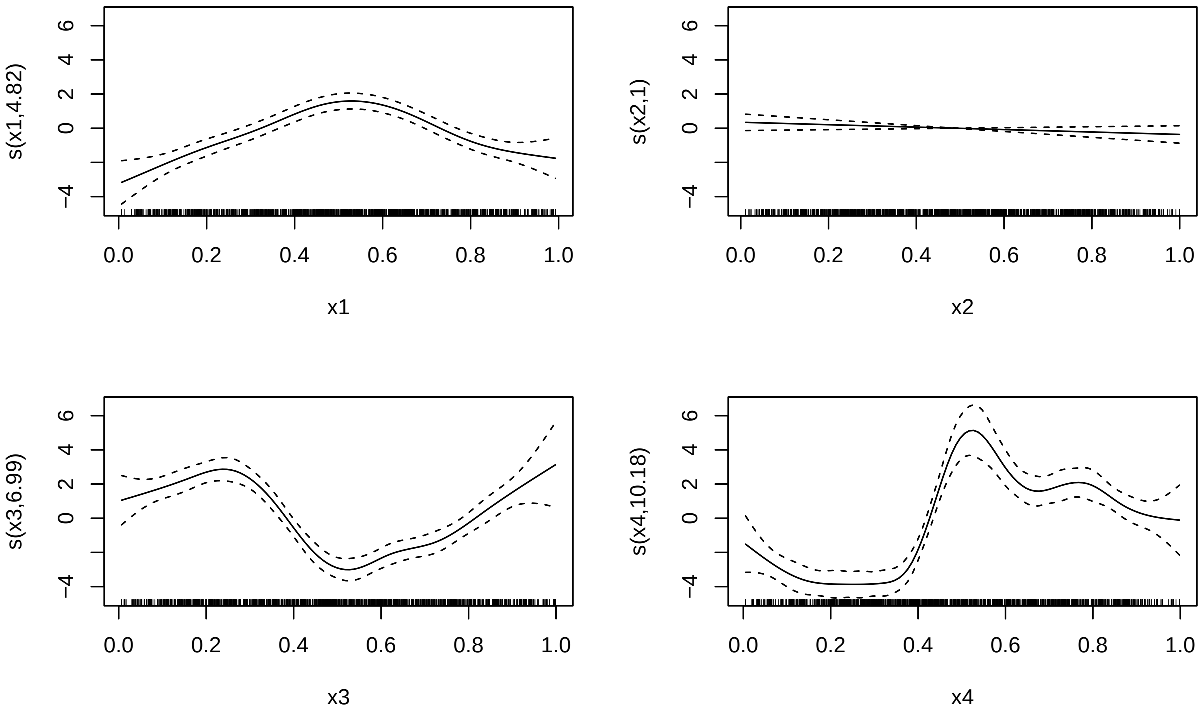

```r
taps_wald_test(fit,test.component=1)
```

```
##    mixed.term fix.df fix.chisq fix.pvalue fix.indices smooth.df smooth.chisq
##        <char>  <num>     <num>      <num>      <char>     <num>        <num>
## 1:      s(x1)      1  4.613863 0.03171452    reported  4.835561     85.00377
##    smooth.pvalue
##            <num>
## 1:             0
```

```r
taps_score_test(fit,test.component=1)
```

```
##    smooth.term smooth.pvalue method
##         <char>         <num> <char>
## 1:       s(x1)  1.144928e-19 davies
```

We also investigate if the link function will infuence the performances of the Wald test and score test:

```r
y=rbinom(n,1,pnorm((eta-2)/sqrt(3.2)))
# We adjust a factor sqrt(3.2), making the variance of liability score
# explaining similar from probit regression to logistics regression
fit=gam(y~s(x1,bs="AMatern",k=10,m=100)+s(x2,bs="cr",k=10)+s(x3,bs="cr",k=10)+
          s(x4,bs="cr",k=15),method="REML",family=binomial(link="probit"))
summary(fit)
```

```
##
## Family: binomial
## Link function: probit
##
## Formula:
## y ~ s(x1, bs = "AMatern", k = 10, m = 100) + s(x2, bs = "cr",
##     k = 10) + s(x3, bs = "cr", k = 10) + s(x4, bs = "cr", k = 15)
```

```
##
## Parametric coefficients:
##             Estimate Std. Error z value Pr(>|z|)
## (Intercept)  1.13524    0.08684   13.07   <2e-16 ***
## ---
## Signif. codes:  0 '***' 0.001 '**' 0.01 '*' 0.05 '.' 0.1 ' ' 1
##
## Approximate significance of smooth terms:
##          edf Ref.df  Chi.sq p-value
## s(x1) 5.273  6.300 104.339  <2e-16 ***
## s(x2) 1.000  1.001   0.024   0.879
## s(x3) 6.526  7.647 155.296  <2e-16 ***
## s(x4) 9.693 11.298 253.960  <2e-16 ***
## ---
## Signif. codes:  0 '***' 0.001 '**' 0.01 '*' 0.05 '.' 0.1 ' ' 1
##
## R-sq.(adj) =  0.646   Deviance explained =   62%
## -REML = 277.38  Scale est. = 1         n = 1000
```

```
plot(fit,page=1)
```

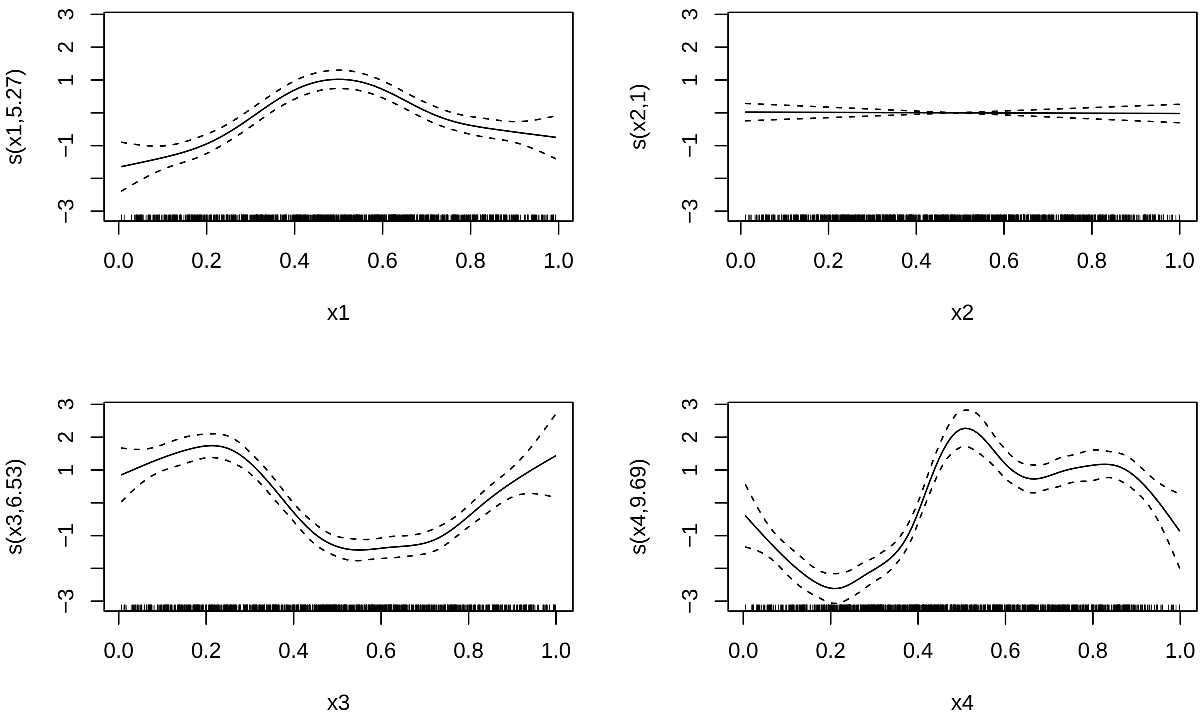

```
taps_wald_test(fit,test.component=1)
```

```
##    mixed.term fix.df fix.chisq  fix.pvalue fix.indices smooth.df smooth.chisq
##        <char>  <num>     <num>       <num>      <char>     <num>        <num>
## 1:      s(x1)      1  10.12673 0.001461345    reported  5.299945     97.66825
##    smooth.pvalue
##            <num>
## 1:             0
```

```r
taps_score_test(fit,test.component=1)
```

```
##    smooth.term smooth.pvalue method
##         <char>         <num> <char>
## 1:       s(x1)  1.805742e-20 davies
```

It is easy to see, by applying median GAM (quantile = 0.5), the power of the Wald test improves (p-value reduces from 4.12E-6 to 1.66E-7).

#### 2.8 Setting of Poisson Outcome

For Poisson data, we generate data and fit the model as follows:

```r
eta=f1+f3+f4
eta=eta/sd(eta)
## Also, we adjust the scale of eta to make it explain a reasonable
## proportion of variance of the outcome
y=rpois(n,exp(eta))
fit=gam(y~s(x1,bs="AMatern",k=10,m=100)+s(x2,bs="cr",k=10)+
          s(x3,bs="cr",k=10)+s(x4,bs="cr",k=15),method="REML",family=poisson())
summary(fit)
```

```
##
## Family: poisson
## Link function: log
##
## Formula:
## y ~ s(x1, bs = "AMatern", k = 10, m = 100) + s(x2, bs = "cr",
##     k = 10) + s(x3, bs = "cr", k = 10) + s(x4, bs = "cr", k = 15)
##
## Parametric coefficients:
##             Estimate Std. Error z value Pr(>|z|)
## (Intercept)  1.02389    0.02207   46.39   <2e-16 ***
## ---
## Signif. codes:  0 '***' 0.001 '**' 0.01 '*' 0.05 '.' 0.1 ' ' 1
##
## Approximate significance of smooth terms:
##          edf Ref.df   Chi.sq p-value
## s(x1)  6.200  7.182  448.199  <2e-16 ***
## s(x2)  1.007  1.014    0.078   0.796
## s(x3)  8.134  8.755 1241.407  <2e-16 ***
## s(x4) 11.924 13.141 1780.373  <2e-16 ***
## ---
## Signif. codes:  0 '***' 0.001 '**' 0.01 '*' 0.05 '.' 0.1 ' ' 1
##
## R-sq.(adj) =  0.846   Deviance explained = 79.2%
## -REML = 1930.6  Scale est. = 1         n = 1000
```

```r
plot(fit,page=1)
```

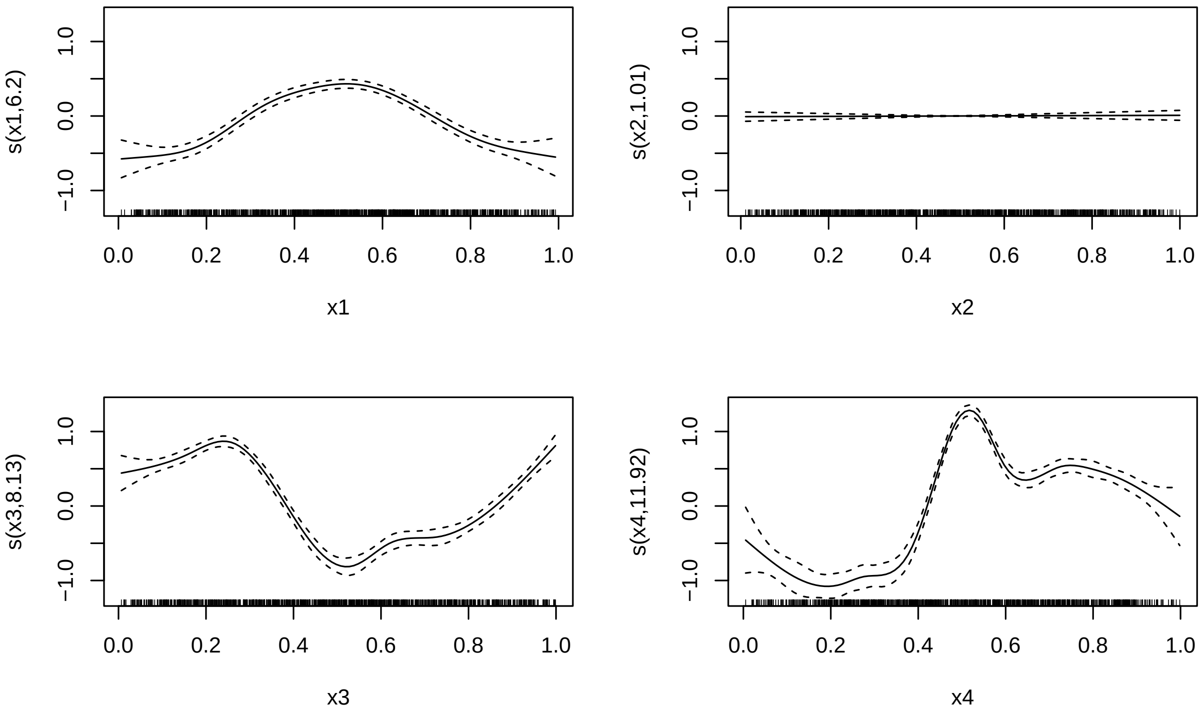

```r
taps_wald_test(fit,test.component=1)
```

```
##    mixed.term fix.df fix.chisq fix.pvalue fix.indices smooth.df smooth.chisq
##        <char>  <num>     <num>      <num>      <char>     <num>        <num>
## 1:      s(x1)      1  1.029418  0.3102954    reported  6.182176     447.0005
##    smooth.pvalue
##            <num>
## 1:             0
```

```r
taps_score_test(fit,test.component=1)
```

```
##    smooth.term smooth.pvalue method
##         <char>         <num> <char>
## 1:       s(x1)  3.348328e-97 davies
```

#### 2.9 Setting of Survival Time Outcome

We also consider the Cox-hazard regression, where the data is generated and the model is fitted as follows:

```r
library(survival)
eta=f1+f3+f4
lambda <- 0.1
shape <- 2
scale <- 1 / (lambda*exp(eta+rnorm(n,0,sqrt(2))))
time <- rweibull(n, shape = shape, scale = scale)
censor_time <- rexp(n, rate = 0.1)
# we consider the censor proportion is 10%
observed_time <- pmin(time, censor_time)
status <- as.numeric(time <= censor_time)
data <- data.frame(
```

```r
time = observed_time,
status = status,
x1 = x1,
x2 = x2,
x3 = x3,
x4 = x4
)
fit <- gam(Surv(time, status) ~ s(x1,bs="AMatern",k=10,m=100)+
             s(x2, bs="cr", k=10)+s(x3, bs="cr", k=10)+s(x4, bs="cr", k=15),
         data = data,
         method = "REML",
         family = cox.ph())
# summary() does not work for survival outcome
taps_wald_test(fit,test.component=1)
```

```
##    mixed.term fix.df fix.chisq   fix.pvalue fix.indices smooth.df smooth.chisq
##        <char>  <num>     <num>        <num>      <char>     <num>        <num>
## 1:      s(x1)      1  13.88879 0.0001939516    reported  6.548708      312.554
##    smooth.pvalue
##            <num>
## 1:             0
```

```r
# score test is not available for survival outcome
```

#### 2.10 Setting of Ordinal Category Outcome

Finally, we investigate the ordinal category outcome with number of category $R = 3$:

```r
eta = (f1 + f3 + f4)/sqrt(3.2)
u = eta -mean(eta) + rnorm(n, 0, sqrt(2))
alpha = c(-Inf, -3, 3, Inf)
R = length(alpha) - 1  # R=3
y = numeric(n)
for (ii in 1:R) {
  y[u > alpha[ii] & u <= alpha[ii+1]] <- ii
}
fit=gam(y~s(x1,bs="AMatern",k=10,m=100)+s(x2,bs="cr",k=10)+
          s(x3,bs="cr",k=10)+s(x4,bs="cr",k=15),method="REML",family=ocat(R=R))
summary(fit)
```

```
##
## Family: Ordered Categorical(-1,6.71)
## Link function: identity
##
## Formula:
## y ~ s(x1, bs = "AMatern", k = 10, m = 100) + s(x2, bs = "cr",
##     k = 10) + s(x3, bs = "cr", k = 10) + s(x4, bs = "cr", k = 15)
##
## Parametric coefficients:
##             Estimate Std. Error z value Pr(>|z|)
## (Intercept)  2.96689    0.08913   33.29   <2e-16 ***
## ---
## Signif. codes:  0 '***' 0.001 '**' 0.01 '*' 0.05 '.' 0.1 ' ' 1
##
## Approximate significance of smooth terms:
```

```
##           edf Ref.df  Chi.sq p-value
## s(x1)  5.077  6.116  99.292  <2e-16 ***
## s(x2)  1.000  1.000   4.457  0.0348 *
## s(x3)  6.508  7.601 192.420  <2e-16 ***
## s(x4) 10.728 12.298 767.805  <2e-16 ***
## ---
## Signif. codes:  0 '***' 0.001 '**' 0.01 '*' 0.05 '.' 0.1 ' ' 1
##
## Deviance explained = 59.9%
## -REML = 471.23  Scale est. = 1         n = 1000
```

```
plot(fit,page=1)
```

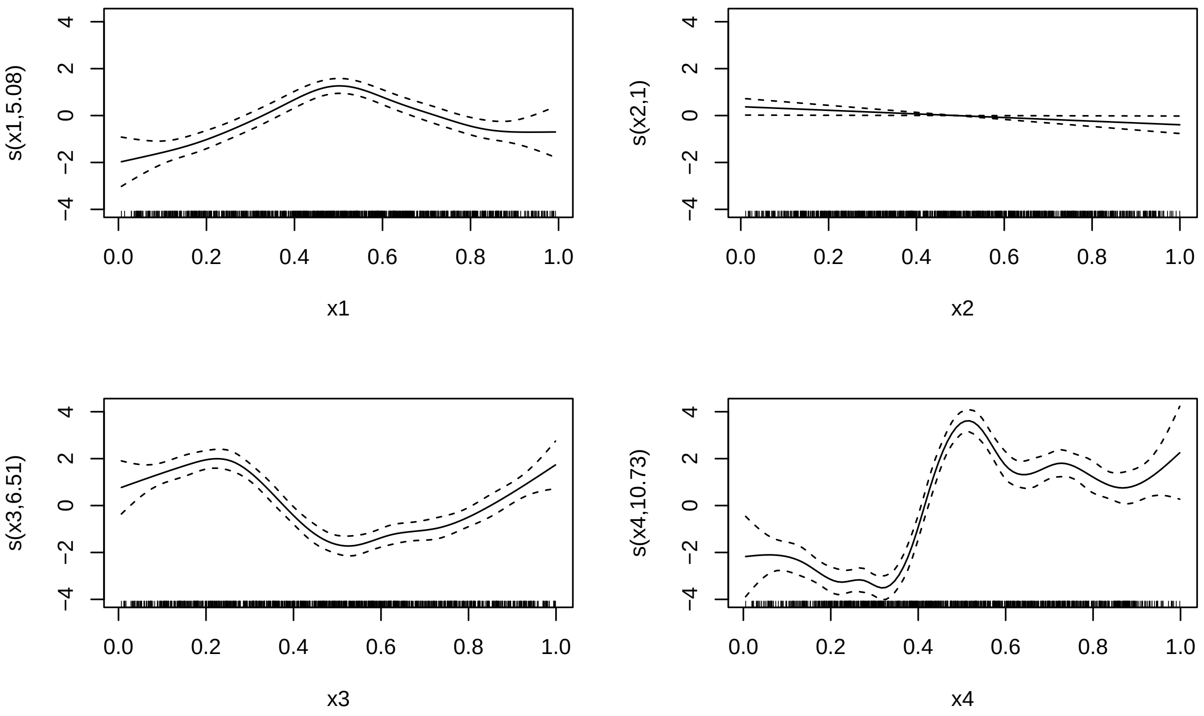

```
taps_wald_test(fit)
```

```
##    mixed.term fix.df fix.chisq  fix.pvalue fix.indices smooth.df smooth.chisq
##        <char>  <num>     <num>       <num>      <char>     <num>        <num>
## 1:      s(x1)      1  6.792287 0.009155254    reported  5.116376     91.93117
##    smooth.pvalue
##            <num>
## 1:             0
```

```
# score test does not support ordinal category outcome
```

#### 2.11 Setting of Varying Coefficient Model

The implementation of varying coefficient model is almost the same as the standard GAM in mgcv. Here, we just show the quasi codes.

```
X=MASS::mvrnorm(n,rep(0,4),matrix(0.25,4,4)+0.75*diag(4))
Z=MASS::mvrnorm(n,rep(0,4),matrix(0.25,4,4)+0.75*diag(4))
```

```
x1=qbeta(pnorm(X[,1]),1.5,1.5)
x2=qbeta(pnorm(X[,2]),1.5,1.5)
x3=qbeta(pnorm(X[,3]),1.5,1.5)
x4=qbeta(pnorm(X[,4]),1.5,1.5)
z1=qbeta(pnorm(Z[,1]),1.5,1.5)
z2=qbeta(pnorm(Z[,2]),1.5,1.5)
z3=qbeta(pnorm(Z[,3]),1.5,1.5)
z4=qbeta(pnorm(Z[,4]),1.5,1.5)
```

The first difference is that will generate a covariate z in the model. Then each function is given by

```
f1=smoothed_linearity(x1,a)
t2=2*pi*x2
f2=0.4*sin(t2)+0.8*cos(t2)+1.2*sin(t2)^2+1.6*cos(t2)^3+2*sin(t2)^3
t3=2*(x3-0.5)
f3=3*sin(3*t3)+6*exp(-36*t3^2)
f4=0*x4
eta=f1*z1+f2*z2+f3*z3
y=1+eta+rnorm(n,0,1)*sd(eta/2)
```

Please note that the generating code of $\eta$: `eta=f1*z1+f2*z2+f3*z3`, which is the varying coefficient model $f_1(x_1) \cdot z_1 + f_2(x_2) \cdot z_2 + f_3(x_3) \cdot z_3$. Finally, the implementation of varying coefficient is:

```
fit=gam(y~s(x1,by=z1,bs="AMatern",k=10,m=100)+
        s(x2,by=z2,bs="cr",k=10)+s(x3,by=z3,bs="cr",k=15)
        +s(x4,by=z4,bs="cr",k=10),method="REML",family="gaussain")
```

The codes of implementing the Wald and score tests are exactly the same as ones in GAM:

```
test1=taps_wald_test(fit,test.component=1)
test2=taps_score_test(fit,test.component=1)
```

For other type outcomes, one can just change the family in the function `gam()`.

#### 2.12 The Need of Orthogonal Construction

A natural question is whether the orthogonal construction adds unnecessary complexity compared to a standard mixed-effects model that includes both parametric terms A(x) and a nonparametric smooth term s(x) without enforcing orthogonality. However, such non-orthogonal models suffer from a fundamental identifiability problem: because parametric basis functions (e.g., polynomials, piecewise linear terms) can be represented within the span of typical nonparametric bases (e.g., cubic splines), the GAM cannot uniquely decompose the function into parametric and nonparametric components. This confounding manifests as rank deficiency, leading to undefined parameter estimates and making hypothesis testing impossible. We demonstrate this identifiability failure using simulation examples below, showing that TAPS orthogonal construction is not merely a technical refinement but a necessary solution to enable testing of arbitrary parametric structures.

We first consider a case where the null hypothesis $H_0$ holds and the parametric strucutre to be tested is linearity:

```
n=1000 # a moderate sample size
X=MASS::mvrnorm(n,rep(0,4),matrix(0.25,4,4)+0.75*diag(4))
x1=qbeta(pnorm(X[,1]),1.5,1.5)
x2=qbeta(pnorm(X[,2]),1.5,1.5)
x3=qbeta(pnorm(X[,3]),1.5,1.5)
x4=qbeta(pnorm(X[,4]),1.5,1.5)
a=0 # a=0 corresponding no departure from linearity
```

```r
f1=smoothed_linearity(x1,a)
t2=2*pi*x2
f2=0.4*sin(t2)+0.8*cos(t2)+1.2*sin(t2)^2+1.6*cos(t2)^3+2*sin(t2)^3
t3=2*(x3-0.5)
f3=3*sin(3*t3)+6*exp(-36*t3^2)
f4=0*x4
eta=f1+f2+f3
y=1+eta+rnorm(n,0,1)*sd(eta/2)
X1=cbind(1,x1)
fit=gam(y~x1+s(x1,bs="gp",k=10)+s(x2,bs="cr",k=10)+s(x3,bs="cr",k=15)
        +s(x4,bs="cr",k=10),method="REML")
# we first try gp, which uses exactly the same matern family correlation kernel function
summary(fit)
```

```
##
## Family: gaussian
## Link function: identity
##
## Formula:
## y ~ x1 + s(x1, bs = "gp", k = 10) + s(x2, bs = "cr", k = 10) +
##     s(x3, bs = "cr", k = 15) + s(x4, bs = "cr", k = 10)
##
## Parametric coefficients:
##             Estimate Std. Error t value Pr(>|t|)
## (Intercept)   2.3942     0.1402  17.083  < 2e-16 ***
## x1            0.9175     0.2567   3.574 0.000368 ***
## ---
## Signif. codes:  0 '***' 0.001 '**' 0.01 '*' 0.05 '.' 0.1 ' ' 1
##
## Approximate significance of smooth terms:
##            edf   Ref.df       F p-value
## s(x1)  0.04266  0.08415   0.003   0.988
## s(x2)  8.14463  8.77727 157.489  <2e-16 ***
## s(x3) 12.58048 13.61521 214.052  <2e-16 ***
## s(x4)  1.00589  1.01175   0.054   0.828
## ---
## Signif. codes:  0 '***' 0.001 '**' 0.01 '*' 0.05 '.' 0.1 ' ' 1
##
## Rank: 42/43
## R-sq.(adj) =  0.804   Deviance explained = 80.9%
## -REML = 1973.9  Scale est. = 2.8049    n = 1000
```

```r
taps_score_test(fit,test.component=1)
```

```
##    smooth.term smooth.pvalue method
##         <char>         <num> <char>
## 1:       s(x1)     0.3235869 davies
```

We first try the 'gp' spline, which refers to the Gaussian process. As one can observe, the parametric term `x1` in the GAM fit shows `NaN`, indicating that the linear component is completely confounded with the linear representation within the Gaussian process smooth term, resulting in an unidentifiable model where neither the parametric coefficient nor its standard error can be estimated.

```r
fit=gam(y~x1+s(x1,bs="cr",k=10)+s(x2,bs="cr",k=10)+s(x3,bs="cr",k=15)
        +s(x4,bs="cr",k=10),method="REML")
```

```r
# we first try cr, which refers to cubic regression spline.
# Its construction is effective in mgcv.
summary(fit)
```

```
##
## Family: gaussian
## Link function: identity
##
## Formula:
## y ~ x1 + s(x1, bs = "cr", k = 10) + s(x2, bs = "cr", k = 10) +
##     s(x3, bs = "cr", k = 15) + s(x4, bs = "cr", k = 10)
##
## Parametric coefficients:
##             Estimate Std. Error t value Pr(>|t|)
## (Intercept)  2.85802    0.05296   53.97   <2e-16 ***
## x1           0.00000    0.00000     NaN      NaN
## ---
## Signif. codes:  0 '***' 0.001 '**' 0.01 '*' 0.05 '.' 0.1 ' ' 1
##
## Approximate significance of smooth terms:
##          edf Ref.df       F  p-value
## s(x1)  1.117  1.224  12.937 0.000121 ***
## s(x2)  8.144  8.777 157.385  < 2e-16 ***
## s(x3) 12.580 13.615 214.068  < 2e-16 ***
## s(x4)  1.002  1.004   0.053 0.821137
## ---
## Signif. codes:  0 '***' 0.001 '**' 0.01 '*' 0.05 '.' 0.1 ' ' 1
##
## Rank: 42/43
## R-sq.(adj) =  0.804   Deviance explained = 80.9%
## -REML =   1974  Scale est. = 2.8047    n = 1000
```

```r
print(taps_score_test(fit,test.component=1))
```

```
##    smooth.term smooth.pvalue method
##         <char>         <num> <char>
## 1:       s(x1)     0.2919813 davies
```

For an alternative spline, cubic regression spline (`cr`), the results are the same.

Moreover, the p-value produced by `taps_score_test` is not trustworthy in this case, because this function was specifically designed for orthogonalized structures and requires the input model to satisfy the orthogonality constraint. In such non-orthogonal settings, one should instead examine the results from mgcv's summary directly: even when the null hypothesis holds (i.e., the true function follows the specified parametric structure), the "smooth term" shows highly significant p-values, which is a spurious result caused by the identifiability failure where the smooth term incorrectly absorbs variation that should be attributed to the parametric component.

Next, we consider another parametric structure: the linearity with one breakpoint:

```r
n=1000
X=MASS::mvrnorm(n,rep(0,4),matrix(0.25,4,4)+0.75*diag(4))
x1=qbeta(pnorm(X[,1]),1.5,1.5)
x2=qbeta(pnorm(X[,2]),1.5,1.5)
x3=qbeta(pnorm(X[,3]),1.5,1.5)
x4=qbeta(pnorm(X[,4]),1.5,1.5)
```

```r
a=0 # a=0 corresponding no departure from linearity with one breakpoint
if(a!=0){
X1_design=smoothed_linearity_discontinuity(x1,list(0.5,a))[,-1]
f1=c(X1_design%*%c(4,4,0))
}else{
X1_design=linearity_discontinuity(x1,0.5)[,-1]
f1=c(X1_design%*%c(4,4,0))
}
t2=2*pi*x2
f2=0.4*sin(t2)+0.8*cos(t2)+1.2*sin(t2)^2+1.6*cos(t2)^3+2*sin(t2)^3
t3=2*(x3-0.5)
f3=3*sin(3*t3)+6*exp(-36*t3^2)
f4=0*x4
eta=f1+f2+f3
y=1+eta+rnorm(n,0,1)*sd(eta/2)
fit=gam(y~X1_design+s(x1,bs="cr",k=10)+s(x2,bs="cr",k=10)+s(x3,bs="cr",k=15)+
          s(x4,bs="cr",k=10),method="REML")
summary(fit)
```

```
##
## Family: gaussian
## Link function: identity
##
## Formula:
## y ~ X1_design + s(x1, bs = "cr", k = 10) + s(x2, bs = "cr", k = 10) +
##     s(x3, bs = "cr", k = 15) + s(x4, bs = "cr", k = 10)
##
## Parametric coefficients:
##                      Estimate Std. Error t value Pr(>|t|)
## (Intercept)            5.1551     0.3924  13.138   <2e-16 ***
## X1_designx             0.0000     0.0000     NaN      NaN
## X1_design0.5_jump      3.7017     0.3429  10.794   <2e-16 ***
## X1_design0.5_slope    -4.5850     2.9872  -1.535    0.125
## ---
## Signif. codes:  0 '***' 0.001 '**' 0.01 '*' 0.05 '.' 0.1 ' ' 1
##
## Approximate significance of smooth terms:
##           edf Ref.df      F  p-value
## s(x1)   1.845  2.451  23.80 3.25e-07 ***
## s(x2)   7.545  8.431  79.45  < 2e-16 ***
## s(x3) 11.381 12.853 116.16  < 2e-16 ***
## s(x4)   1.002  1.003   0.00    0.997
## ---
## Signif. codes:  0 '***' 0.001 '**' 0.01 '*' 0.05 '.' 0.1 ' ' 1
##
## Rank: 44/45
## R-sq.(adj) =  0.799   Deviance explained = 80.4%
## -REML = 2290.7  Scale est. = 5.4027    n = 1000
```

```r
taps_score_test(fit,test.component=1)
```

```
##    smooth.term smooth.pvalue method
##         <char>         <num> <char>
## 1:       s(x1)     0.2173216 davies
```

We observe that the results exhibit even more severe identifiability issues compared to the linear structure case. Because the target parametric structure contains a jump discontinuity at x=0.5, which is inherently non-smooth, the cubic regression spline (`cr`) cannot fully represent this discontinuity within its smooth basis. Consequently, the parametric component estimates become unstable and difficult to interpret, as the model struggles to separate the discontinuous parametric structure from the smooth nonparametric term. This represents a more severe breakdown than the linear structure case: whereas smooth parametric structures like linearity can be well-approximated by most spline bases, non-smooth structures such as jumps or changepoints fundamentally conflict with the smooth basis representation, leading to systematic confounding and unreliable inference in non-orthogonal models.

In summary, non-orthogonal models suffer from fundamental identifiability failures where parametric components become confounded with nonparametric smooth terms, manifesting as undefined estimates (`NaN`) for smooth structures and spurious complexity (inflated `edf`) for non-smooth structures. TAPS orthogonal construction is therefore not a technical refinement but a necessary solution to enable rigorous hypothesis testing of arbitrary parametric structures within the GAM framework.

### 3 Simulation Results

In this section, we include additional figures from both the simulation studies and real data analyses to further illustrate the performance and practical applications of our method. We used the following figure as an example to illustrate how do we evaluate the results.

#### 3.1 Summary of Simulation Results

We first evaluate whether the null distributions of the Wald and score test statistics are correctly specified. If so, the associated $p$-values should follow a uniform distribution on the interval $[0, 1]$. When the sorted $p$-values systematically lie above the diagonal in a quantile-quantile (QQ) plot, it indicates inflated Type II error rates (i.e., the test is overly conservative). Conversely, when the sorted $p$-values lie below the diagonal, it suggests inflated Type I error rates (i.e., the test is overly liberal). Figure S6A-S6B shows the QQ plots of $p$-values under the null across the four testing scenarios. We observe that the empirical quantiles of the score test closely follow the uniform distribution, regardless of the sample size or whether the response distribution is continuous or discrete. These results confirm that the score test controls both Type I and Type II error rates well. In contrast, the $p$-values from the Wald test tend to deviate upward from the uniform quantiles, indicating that the Wald test is more conservative and prone to inflated Type II error. This conservativeness arises from the penalization of nonparametric terms in `mgcv`, which affects the estimation of their effective degrees of freedom (edf). Since the null distribution of the Wald statistic depends on numerical approximation [15], it may become inaccurate when the nonparametric component has near-zero edf, thus leading to inflated Type II errors.

We then investigate the power of the two testing procedures. To simulate deviations from the null hypothesis, we introduce a deviation parameter $d$, where larger values of $d$ correspond to stronger departures of $f_1$ from the parametric structure under the null. As shown in Figure S6C-S6D. The score test consistently achieves high power in detecting deviations from the null across all scenarios under Gaussian and Poisson outcomes, with slightly lower power under the binary distribution. As the deviation parameter $d$ increases, power quickly approaches 1, especially when the sample size exceeds 1500. The Wald test is generally less powerful than the score test but still shows a clear upward trend and converges to high power with moderate sample sizes. Across the four scenarios, the linearity and interaction tests are particularly sensitive to deviations, with both tests attaining high power rapidly. In contrast, the piecewise linearity and discontinuity scenarios require larger sample sizes or stronger deviations (larger $d$) to achieve comparable power. Overall, both tests become increasingly effective as the deviation from the null grows and the available data become more informative.

Despite its strong performance, the score test is limited to outcomes from the exponential family, whereas the Wald test, though slightly less powerful, is not subject to this restriction. Especially, the Wald test can be applied to quantile GAMs [4], such as the median GAM (quantile = 0.5). We further examine the performance of the Wald test when the outcome is a survival time or an ordinal categorical variable, as well as its robustness to outliers in GAM and median GAM (Figure S7-S9). Similar simulation studies for evaluating the performance of the varying-coefficient model are presented in Figure S10.

To evaluate the calibration of different score test approximation methods, we conducted simulations under the null hypothesis where both the true data-generating process and the hypothesized parametric structure were linear. We varied sample sizes from 500 to 4,500 and performed 10,000 replications at each sample size to ensure sufficient precision for examining tail behavior. For each replication, we applied six score test methods, Davies, Imhof, Liu, Satterthwaite, Hall-Buckley-Eagleson (HBE), and Wood, and examined whether their $p$-values followed the expected uniform distribution under the null. The details of the first three methods can be found in Duchesne and De Micheaux [3], while the details of the last three methods can be found in Bodenham and Adams [1]. Figure S11-S13 presents QQ plots comparing the empirical and theoretical quantiles of $p$-values across three outcome distributions (Gaussian, Poisson, and binary).

Several consistent patterns emerged across all outcome types. First, the Davies and Imhof methods produced virtually identical results, as expected given their theoretical equivalence in computing exact distributions of weighted chi-squared statistics. Second, the Liu and HBE approximations were nearly indistinguishable from each other and exhibited no visually discernible deviation from the exact methods, indicating excellent calibration. Third, the Satterthwaite approximation showed noticeable inflation in the tail region, suggesting mildly inflated Type I error rates compared to the exact Davies method. Finally, the Wood method exhibited conservative behavior, with observed $p$-values lying above the diagonal, indicating reduced power and inflated Type II error rates. These findings demonstrate that while most approximation methods maintain proper error control under the null, practitioners should be aware of the Satterthwaite method's slight anticonservatism in tail regions and the Wood method's overly conservative behavior, particularly when stringent significance thresholds are applied.

#### 3.2 Exhibition of Simulation Figures

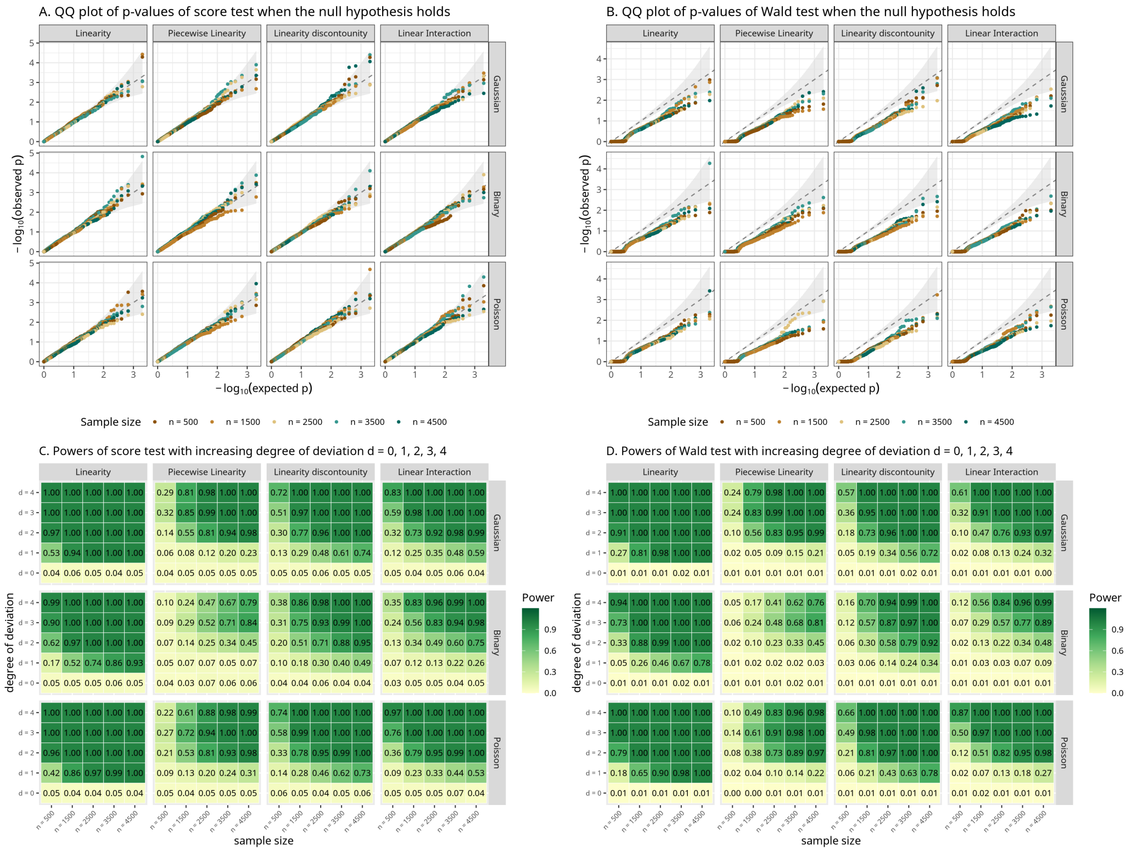

Figure S6: Performance of the Wald and score tests under the null and alternative hypotheses. **A**. QQ plots of $p$-values under the null hypothesis for the score test, and **B**. for the Wald test, across four structure types (columns) and three outcome distributions (rows), with sample sizes ranging from $n = 500$ to $n = 4500$. Each point compares the empirical distribution of $p$-values against the expected uniform distribution (gray dashed line), with the 95% confidence band constructed from the pointwise Beta distribution. **C**. Heatmaps of power under the alternative hypothesis for the score test, and **D**. for the Wald test, with increasing deviation from the null controlled by parameter $d$ (rows) and sample size (columns). Both color and overlaid numeric values indicate rejection frequencies (power).

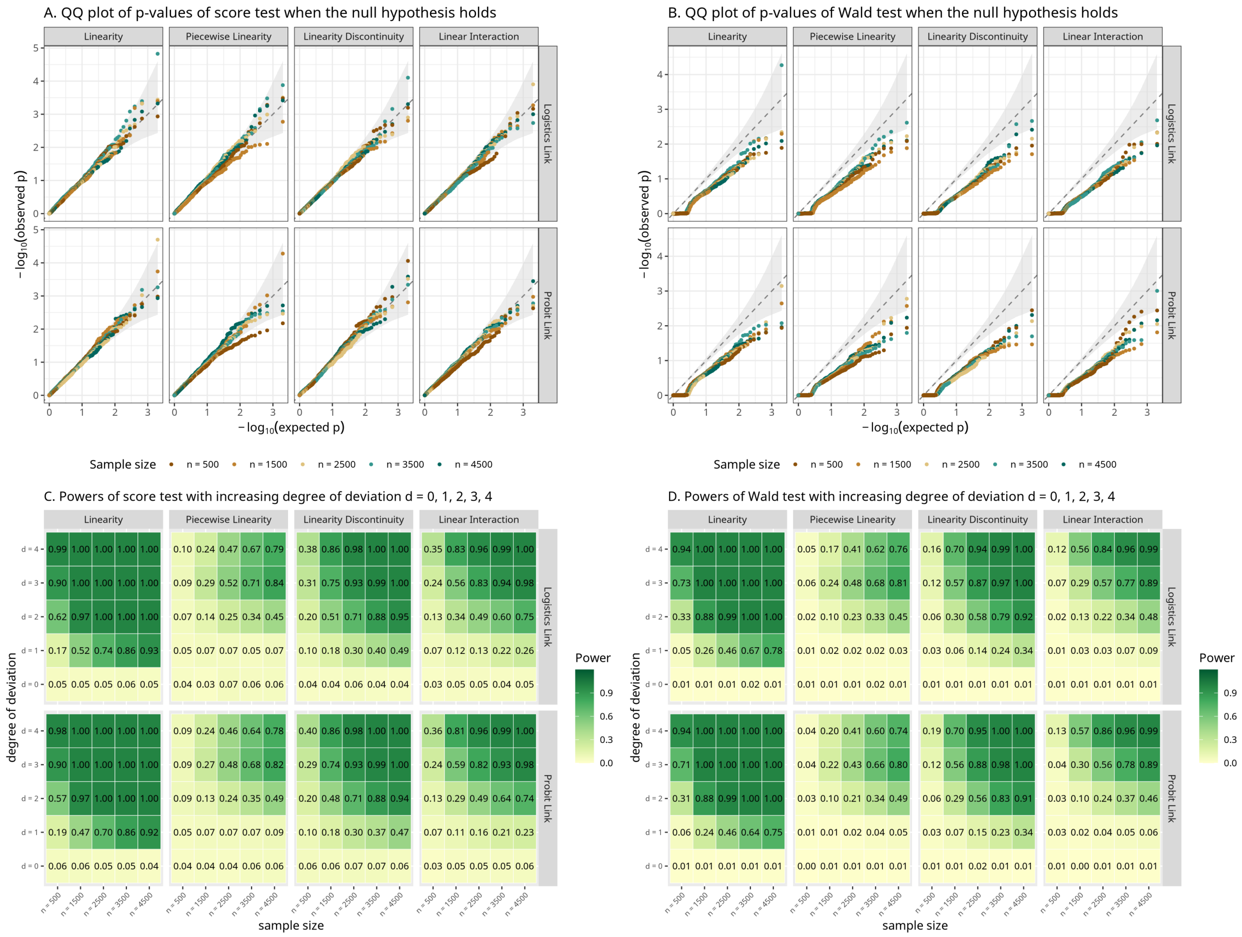

Figure S7: Performance of the Wald and score tests for testing linearity under logistic and probit link functions. **A**. QQ plots of $p$-values from the score test under the null hypothesis, across four parametric structures: linearity, piecewise linearity, linearity discontinuity, and linear interaction. **B**. QQ plots of $p$-values from the Wald test under the same settings. Each point compares the empirical distribution of $p$-values against the expected uniform distribution (gray dashed line), with the 95% confidence band constructed from the pointwise Beta distribution, across sample sizes $n = 500$ to $n = 4500$. **C**. Power heatmaps for the score test as the degree of deviation $d$ increases from 0 to 4. **D**. Corresponding power results for the Wald test. Each cell shows the rejection frequency over 1,000 simulations under logistic and probit links. Across all scenarios, the performances of the score test and Wald test for logistics regression and probit regression are almost the same.

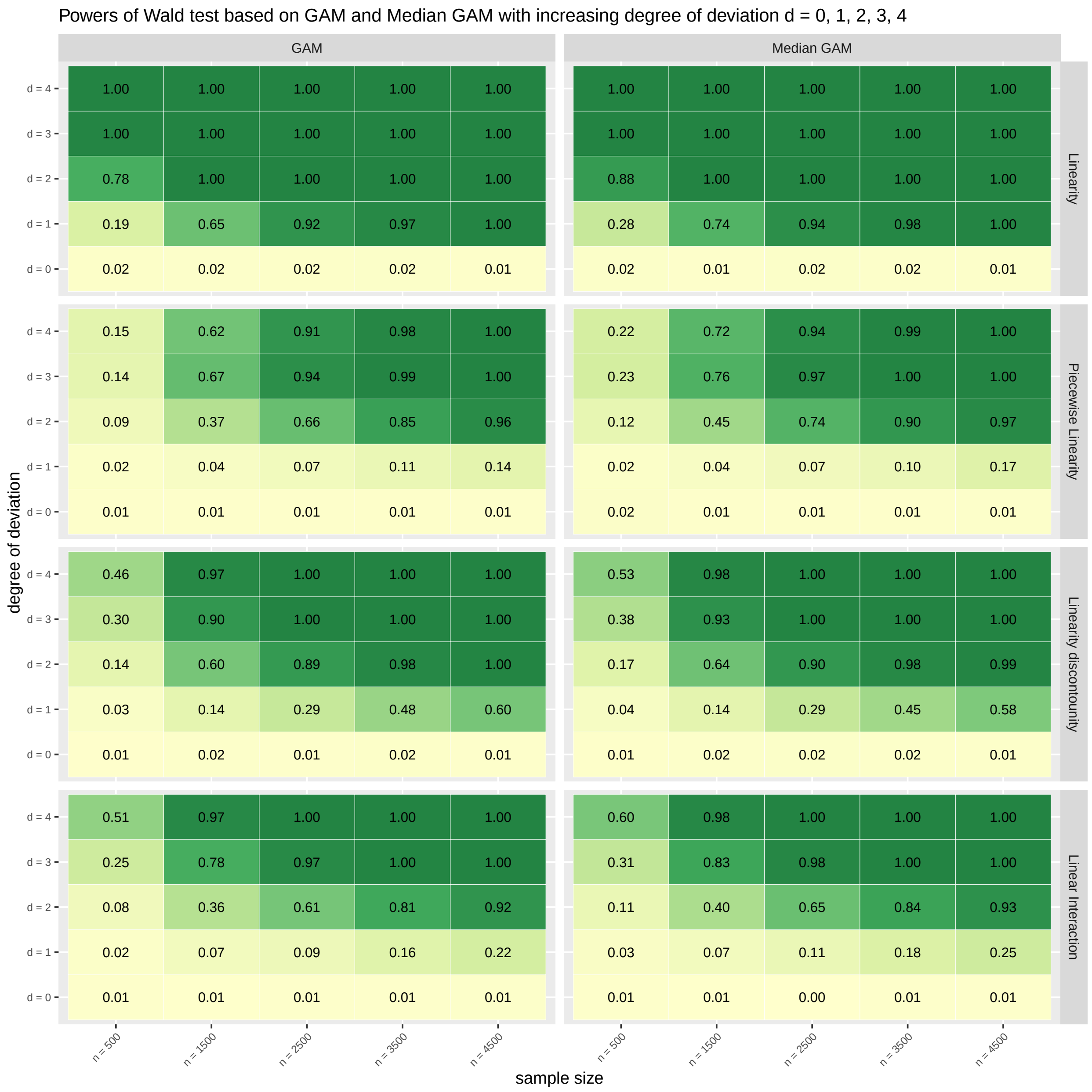

Figure S8: Comparison of Wald test power between GAM and Median GAM in the presence of outliers. The figure shows rejection frequencies across four structural hypotheses: linearity, piecewise linearity, linearity with discontinuity, and linear interaction. Each row corresponds to increasing deviation levels ($d = 0, 1, 2, 3, 4$), and each column represents increasing sample sizes ($n = 500, 1500, 2500, 3500, 4500$). **Left block**: Power of the Wald test under standard GAM fitting. **Right block**: Power under Median GAM, a robust alternative that mitigates the influence of outliers. Across all scenarios, Median GAM demonstrates improved robustness and higher power when outliers are present, particularly in small to moderate sample sizes.

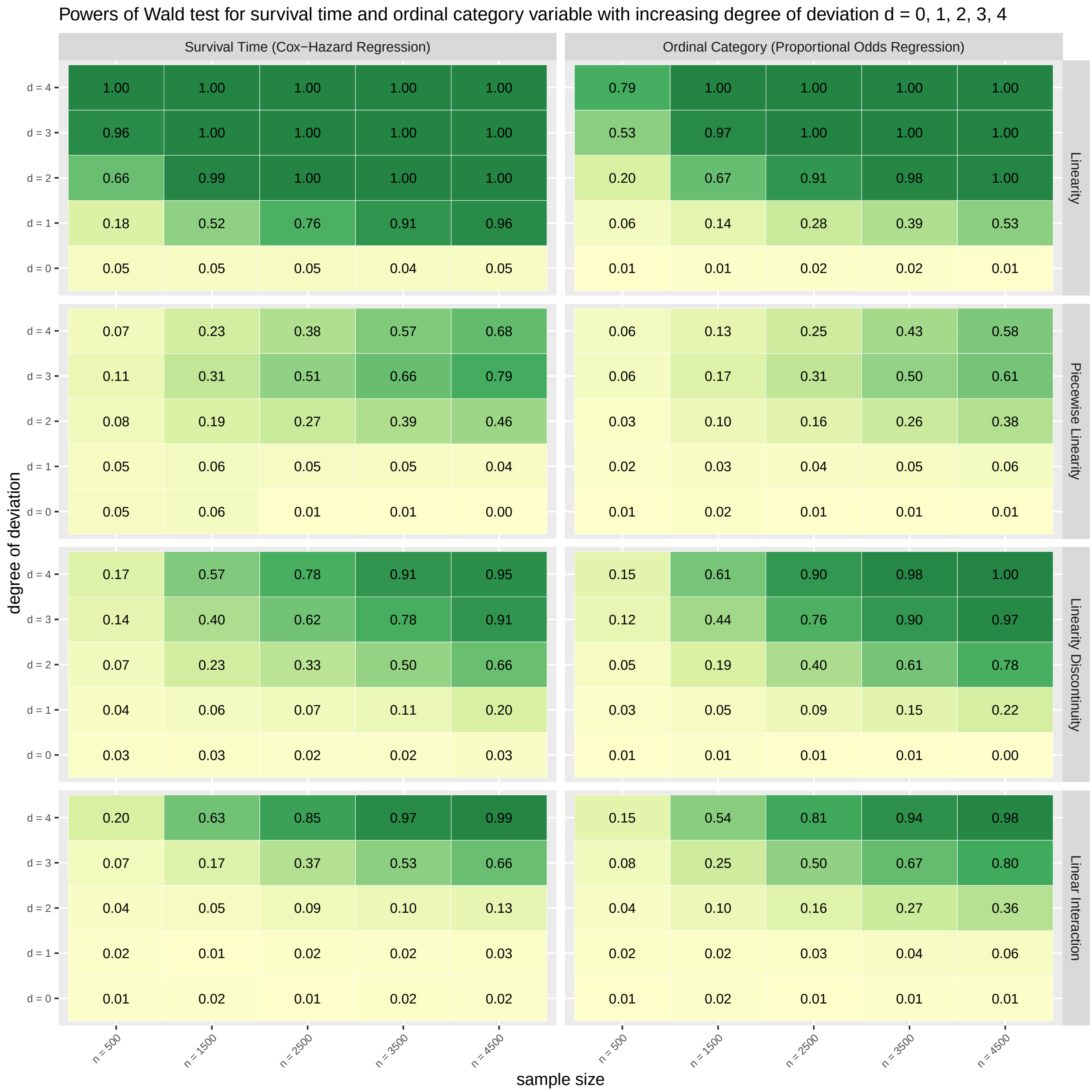

Figure S9: Power of the Wald test for survival time and ordinal outcome variables under increasing deviation from the null. The left block reports results from Cox proportional hazards regression (survival outcome), and the right block corresponds to proportional odds regression (ordinal outcome). Each row represents an increasing level of deviation from the null structure ($d = 0$ to $d = 4$), while each column corresponds to increasing sample sizes ($n = 500$ to $n = 4500$). For each of the four parametric structures—linearity, piecewise linearity, linearity with discontinuity, and linear interaction—the Wald test shows increasing power as deviation and sample size grow. However, the power is overall lower than for Gaussian outcomes, highlighting the greater difficulty of detecting nonlinear structure under non-Gaussian outcome types.

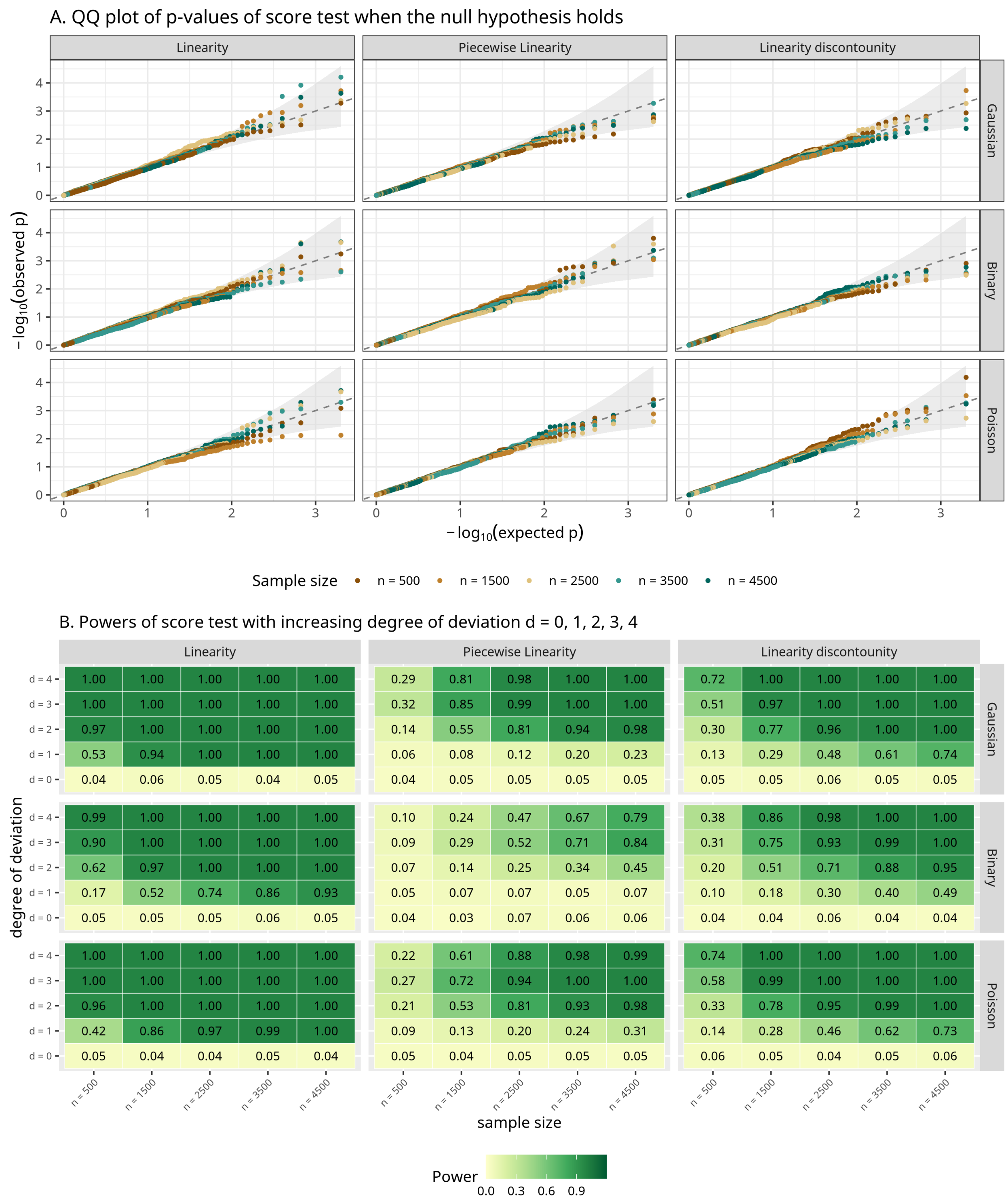

Figure S10: Performance of the score tests under the null and alternative hypotheses, for varying-coefficient model. **A**. QQ plots of $p$-values under the null hypothesis for the score test, across four structure types (columns) and three outcome distributions (rows), with sample sizes ranging from $n = 500$ to $n = 4500$. Each point compares the empirical distribution of $p$-values against the expected uniform distribution (gray dashed line), with the 95% confidence band constructed from the pointwise Beta distribution. **B**. Heatmaps of power under the alternative hypothesis for the score test. Both color and overlaid numeric values indicate rejection frequencies (power).

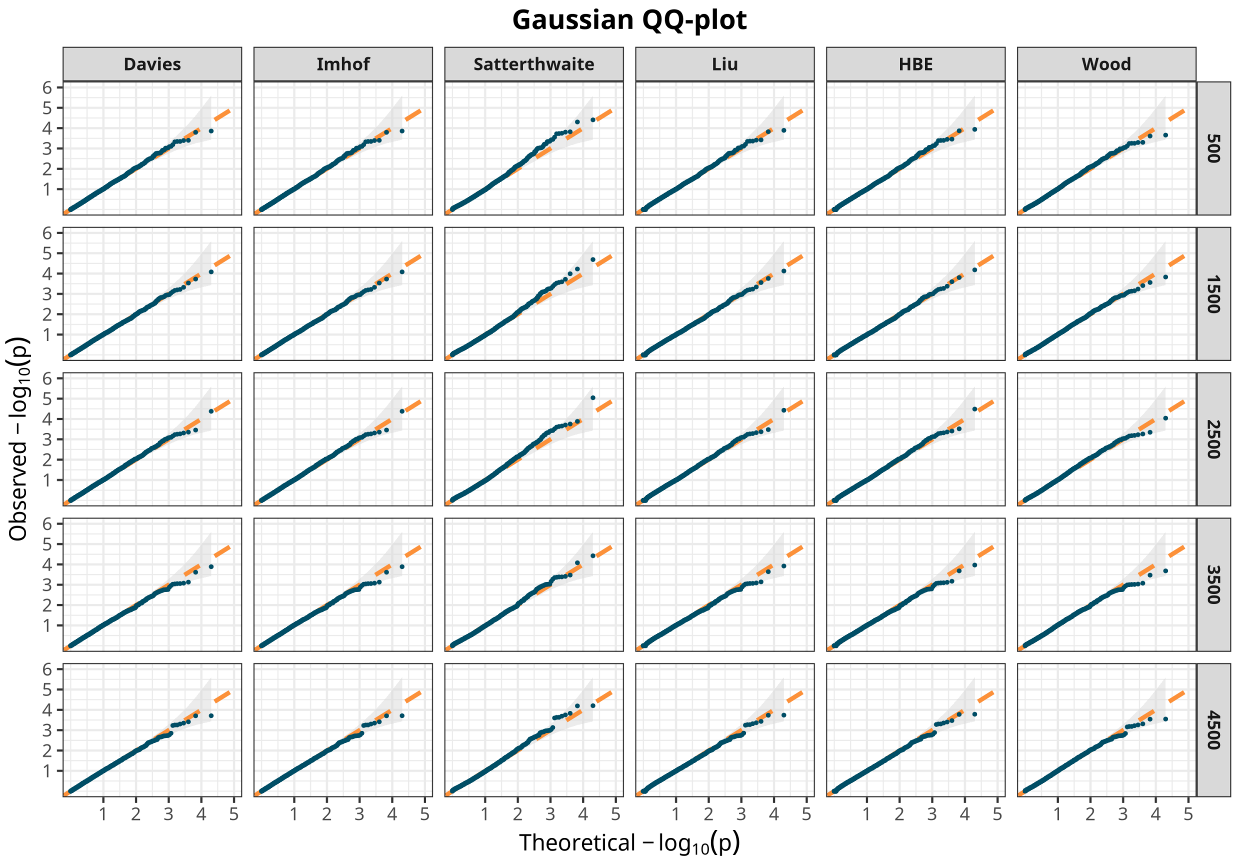

Figure S11: Quantile-quantile (QQ) plots for Type I error control under the null hypothesis of no non-linear interaction across six score test methods for Gaussian outcomes. Each column represents a different method: Davies, Imhof, Satterthwaite, Liu, Hall-Buckley-Eagleson (HBE), and Wood. Each row corresponds to a different sample size (500, 1500, 2500, 3500, 4500). The x-axis shows theoretical $-\log_{10}(p)$ values under the null distribution, and the y-axis shows observed $-\log_{10}(p)$ values from simulations. The orange dashed line represents perfect calibration (y = x). Adherence of the observed values (dark blue curves) to the diagonal line indicates proper Type I error control.

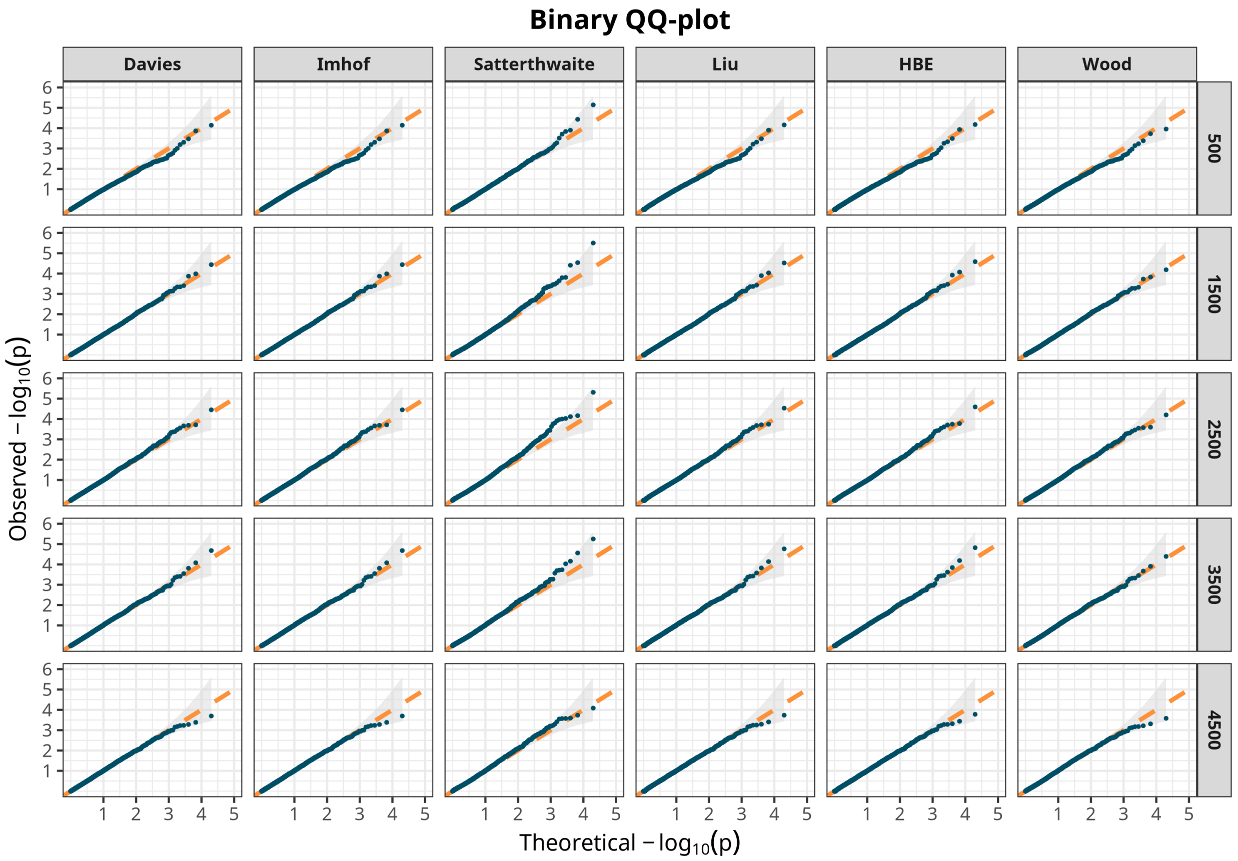

Figure S12: Quantile-quantile (QQ) plots for Type I error control under the null hypothesis of no non-linear interaction across six score test methods for binary outcomes. Each column represents a different method: Davies, Imhof, Satterthwaite, Liu, Hall-Buckley-Eagleson (HBE), and Wood. Each row corresponds to a different sample size (500, 1500, 2500, 3500, 4500). The x-axis shows theoretical $-\log_{10}(p)$ values under the null distribution, and the y-axis shows observed $-\log_{10}(p)$ values from simulations. The orange dashed line represents perfect calibration (y = x). Adherence of the observed values (dark blue curves) to the diagonal line indicates proper Type I error control.

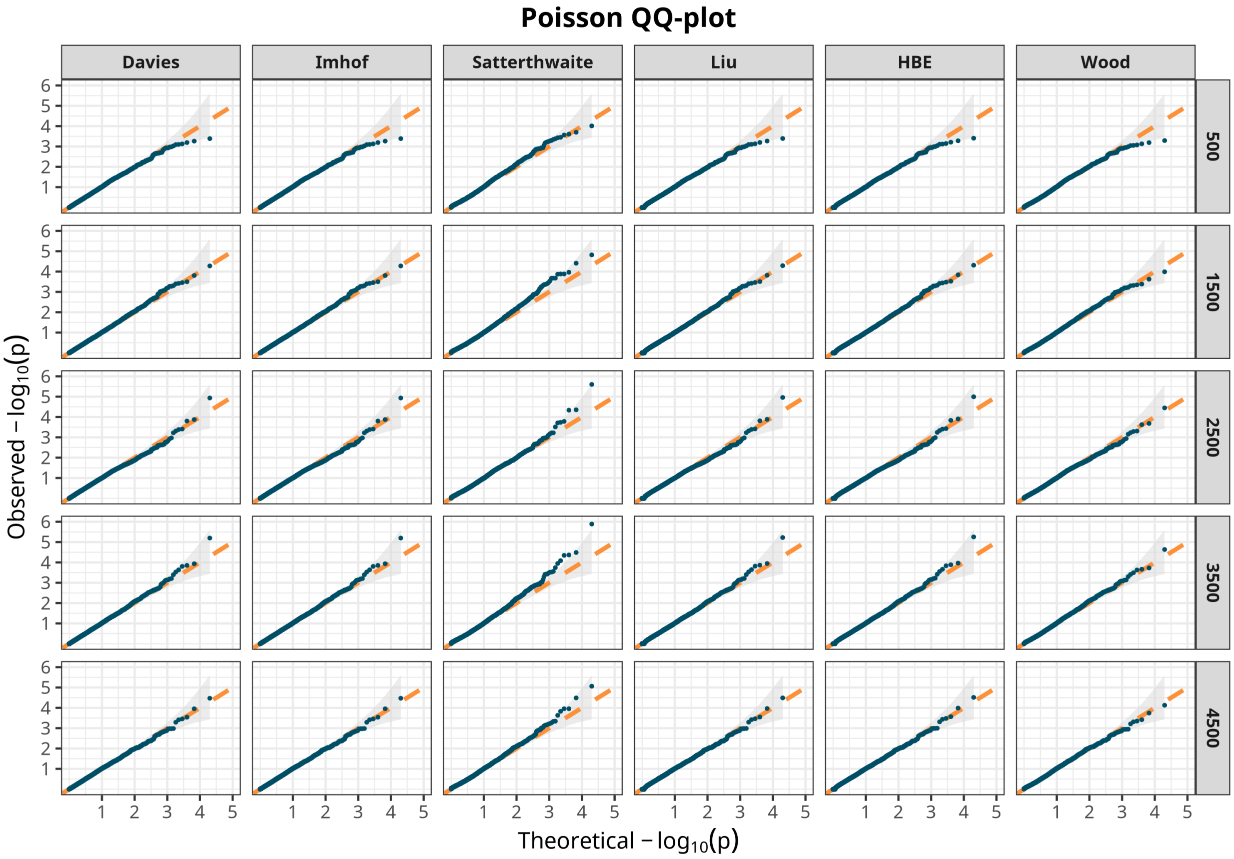

Figure S13: Quantile-quantile (QQ) plots for Type I error control under the null hypothesis of no non-linear interaction across six score test methods for poisson outcomes. Each column represents a different method: Davies, Imhof, Satterthwaite, Liu, Hall-Buckley-Eagleson (HBE), and Wood. Each row corresponds to a different sample size (500, 1500, 2500, 3500, 4500). The x-axis shows theoretical $-\log_{10}(p)$ values under the null distribution, and the y-axis shows observed $-\log_{10}(p)$ values from simulations. The orange dashed line represents perfect calibration (y = x). Adherence of the observed values (dark blue curves) to the diagonal line indicates proper Type I error control.

#### 4 Supplementary Real Data Analysis (with codes)

We provide additional examples to help users understand how to use `mgcv.taps` to define various parametric structures to be tested, along with the corresponding usage patterns within the `mgcv` framework.

##### 4.1 `mcycle` data in `MASS`

The first dataset we analyze is `mcycle`, the motorcycle acceleration data, which records head acceleration during a simulated crash. This dataset is included in the R package `MASS`. We first investigate the pattern of this data:

```
library(MASS)
library(mgcViz)
data("mcycle")
head(mcycle)
```

```
##   times accel
## 1   2.4   0.0
## 2   2.6  -1.3
## 3   3.2  -2.7
## 4   3.6   0.0
## 5   4.0  -2.7
## 6   6.2  -2.7
```

```
fit0 = gam(accel ~ s(times, bs="gp"), data=mcycle, method="REML")
b0 = getViz(fit0)
plot(sm(b0, 1)) +
  l_ciPoly(mul = 5, fill = "#60c5ba", alpha = 0.25) +
  l_rug(mapping = aes(x=x), alpha = 0.25, color="#60c5ba") +
  l_fitLine(colour = "black", size=2) +
  theme_get() +
  xlab("time") + ylab("f(time)") +
  ggtitle("Non-linear effect of time fitted by mgcv") +
  theme(panel.border = element_rect(colour = "black", fill=NA, size=1),
        panel.background = element_rect(fill = "#f4f4f4"),
        plot.title = element_text(size=18))
```

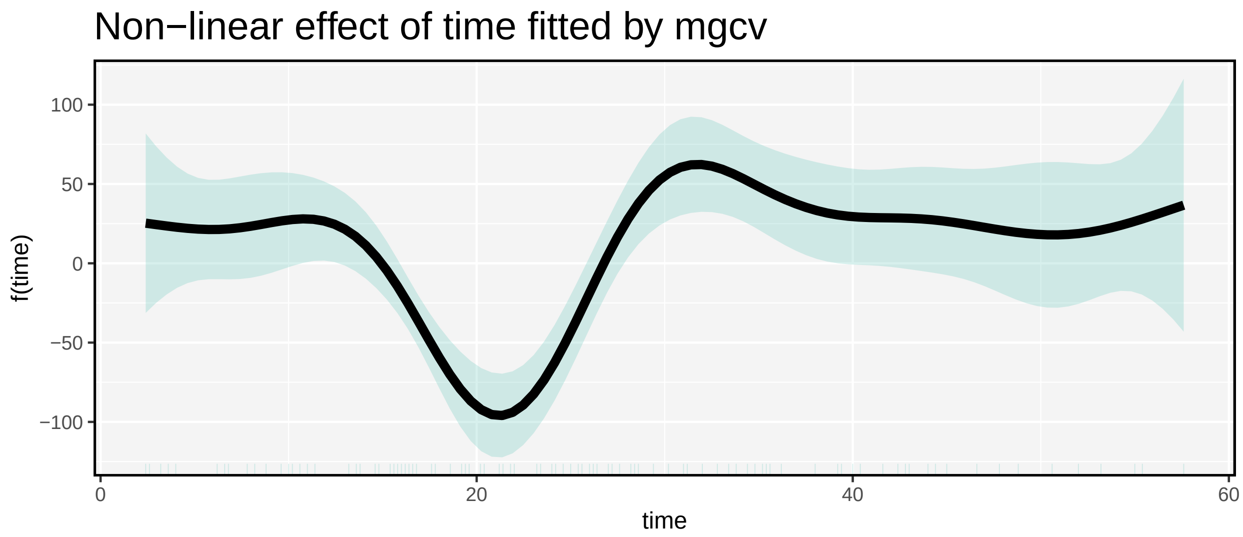

It is easy to see that there may exist four stages of the process of response: a stable stage, a shifting-down stage, a shifting-up stage, and a re-stable stage but with larger variation than the first stable stage. Hence, we consider a hypothesis on whether or not the acceleration is indeed a linear function with three change-points. The null hypothesis then become

$$\mathbf{H}_0: \ \boldsymbol{f}(\boldsymbol{t}) = \alpha_0 + \alpha_1 \boldsymbol{t} + \alpha_2(\boldsymbol{t} - t_{20})_+ + \alpha_3(\boldsymbol{t} - t_{62})_+ + \alpha_4(\boldsymbol{t} - t_{91})_+.$$

We fit use `AMatern` in `s(,bs="smooth.term")` to construct a mixed-effects representation with the fixed effect following the above parametric form:

```
fit1=gam(accel~s(times, bs="AMatern",
                 xt=list(getA=piecewise_linearity, para=c(13.6, 21.2, 30.2))),
           data=mcycle)
b1 = getViz(fit1)
plot(sm(b1, 1), n = 1000) +
  l_ciPoly(mul = 5, fill = "#60c5ba", alpha = 0.25) +
  l_rug(mapping = aes(x=x), alpha = 0.25, color="#60c5ba") +
  l_fitLine(colour = "black", size=2) +
  theme_get() +
  xlab("time") + ylab("f(time)") +
  ggtitle("Mixed effect of time fitted by mgcv.taps with three changepoints") +
  geom_vline(xintercept=13.6, linetype=2, size=0.75, color="grey30") +
  geom_vline(xintercept=21.2, linetype=2, size=0.75, color="grey30") +
  geom_vline(xintercept=30.2, linetype=2, size=0.75, color="grey30") +
  theme(panel.border = element_rect(colour = "black", fill=NA, size=1),
        panel.background = element_rect(fill = "#f4f4f4"),
        plot.title = element_text(size=18))
```

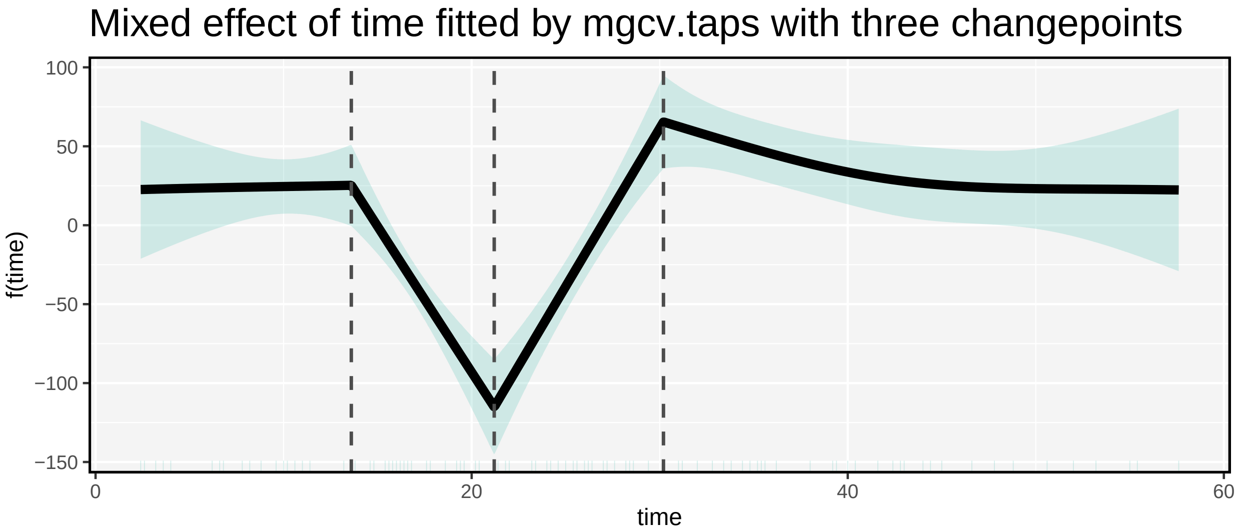

```
taps_score_test(fit1)
```

```
##    smooth.term smooth.pvalue method
##         <char>         <num> <char>
## 1:    s(times)    0.00966903 davies
```

```
taps_wald_test(fit1)
```

```
##    mixed.term fix.df fix.chisq    fix.pvalue fix.indices smooth.df smooth.chisq
##        <char>  <num>     <num>         <num>      <char>     <num>        <num>
## 1:   s(times)      4  467.8091 6.129671e-100    reported  1.481266     4.910157
##    smooth.pvalue
```

```
##             <num>
## 1:    0.05350179
```

It is obvious that the underlying curve in the first three stages is indeed piece-wise linear, but the curve in the fourth stage is likely not a straight line. The score test finds the $p$-value is 0.0086, while the Wald test finds the $p$-vale is 0.0535.

In the section of Supplementary Simulation, we did not cover one important detail: by default, the `"AMatern"` basis in `mgcv.taps` assumes a linear parametric structure. When users wish to test other parametric forms, they must explicitly specify the structure via the `xt` argument inside the `s()` function. For example, to test whether a function follows a piecewise linear form, we use the built-in function `piecewise_linearity` provided in the `mgcv.taps` package:

```
piecewise_linearity <- function(x, para){
  soft_thresholding <- function(x,b){
    y <- x - b
    y[y < 0] <- 0
    return(y)
  }
  knot <- para
  p <- length(knot)
  G <- matrix(0, length(x), p)
  for(i in 1:p){
    G[,i] <- soft_thresholding(x, knot[i])
  }
  basis_matrix <- cbind(1, x, G)
  colnames(basis_matrix) <- c("Intercept", "x", paste0("Smooth_",  seq_len(p)))

  return(basis_matrix)
}
```

This function must take exactly two arguments: the covariate `x` and the user-defined parameter `para`. Both must be passed via the `xt` argument in the form:

```
s(x, bs = "AMatern", xt = list(getA = piecewise_linearity, para = para))
```

Importantly, the shorthand `xt = list(piecewise_linearity, para)` is not recognized by `mgcv.taps`, because it fails to explicitly specify the `getA` and `para` components.

#### 4.2 aids data in R package jmcm

We also consider the CD4 data that has been investigated by a number of researchers [18],[8]. The CD4 cell counts (CD4) of 369 HIV-infected men are included as the response in this dataset, along with six covariates: time since seroconversion (time), age relative to arbitrary origin (age), packs of cigarettes smoked per day (pack), recreational drug use (drug), number of sexual partners (sex), and mental illness score (mental). There are a total of $n = 2376$ CD4 cell count measurements, with several repeated measurements done for everyone at different times over an eight-and-a-half-year span.

The model of the data is given by

$$\log(\mathrm{E}(\mathrm{CD4}_{ij})) = \boldsymbol{Z}_i^\top \boldsymbol{\gamma} + f(\mathrm{time}_{ij}) + \epsilon_{ij},$$

where $\boldsymbol{Z}_i = (1, \mathrm{age}_i, \mathrm{pack}_i, \mathrm{drug}_i, \mathrm{sex}_i, \mathrm{mental}_i)^\top$ and $\{\epsilon_{ij}\}$ are the IID Gaussian noises. We first explore the pattern of time effect by using `mgcv`:

```
library(jmcm)
data("aids")
head(aids)
```

```
##        time cd4  age packs drugs sex cesd    id
## 1 -0.741958 548 6.57     0     0   5    8 10002
## 2 -0.246407 893 6.57     0     1   5    2 10002
## 3  0.243669 657 6.57     0     1   5   -1 10002
## 4 -2.729637 464 6.95     0     1   5    4 10005
## 5 -2.250513 845 6.95     0     1   5   -4 10005
## 6 -0.221766 752 6.95     0     1   5   -5 10005
```

```r
fit0 = gam(cd4~s(time,bs="gp")+s(age,bs="gp")+s(cesd,bs="gp")+
             packs+drugs+sex, data=aids, family=quasipoisson(), method="REML")
## We use quais-Poisson family to fit the CD4+
summary(fit0)
```

```
##
## Family: quasipoisson
## Link function: log
##
## Formula:
## cd4 ~ s(time, bs = "gp") + s(age, bs = "gp") + s(cesd, bs = "gp") +
##     packs + drugs + sex
##
## Parametric coefficients:
##              Estimate Std. Error t value Pr(>|t|)
## (Intercept)  6.481174   0.020312 319.076  < 2e-16 ***
## packs        0.074353   0.006154  12.083  < 2e-16 ***
## drugs        0.065284   0.022894   2.852  0.00439 **
## sex         -0.005378   0.002707  -1.987  0.04704 *
## ---
## Signif. codes:  0 '***' 0.001 '**' 0.01 '*' 0.05 '.' 0.1 ' ' 1
##
## Approximate significance of smooth terms:
##           edf Ref.df      F p-value
## s(time) 7.017  8.094 74.240  <2e-16 ***
## s(age)  1.003  1.006  1.078  0.2992
## s(cesd) 1.003  1.006  6.557  0.0104 *
## ---
## Signif. codes:  0 '***' 0.001 '**' 0.01 '*' 0.05 '.' 0.1 ' ' 1
##
## R-sq.(adj) =  0.273   Deviance explained = 28.2%
## -REML = 7113.8  Scale est. = 150.32    n = 2376
```

```r
b0 = getViz(fit0)
plot(sm(b0, 1)) +
  l_ciPoly(mul = 5, fill = "#60c5ba", alpha = 0.25) +
  l_rug(mapping = aes(x=x), alpha = 0.25, color="#60c5ba") +
  l_fitLine(colour = "black", size=2) +
  theme_get() +
  xlab("time") + ylab("f(time)") +
  ggtitle("Non-linear effect of time fitted by mgcv") +
  theme(panel.border = element_rect(colour = "black", fill=NA, size=1),
        panel.background = element_rect(fill = "#f4f4f4"))
```

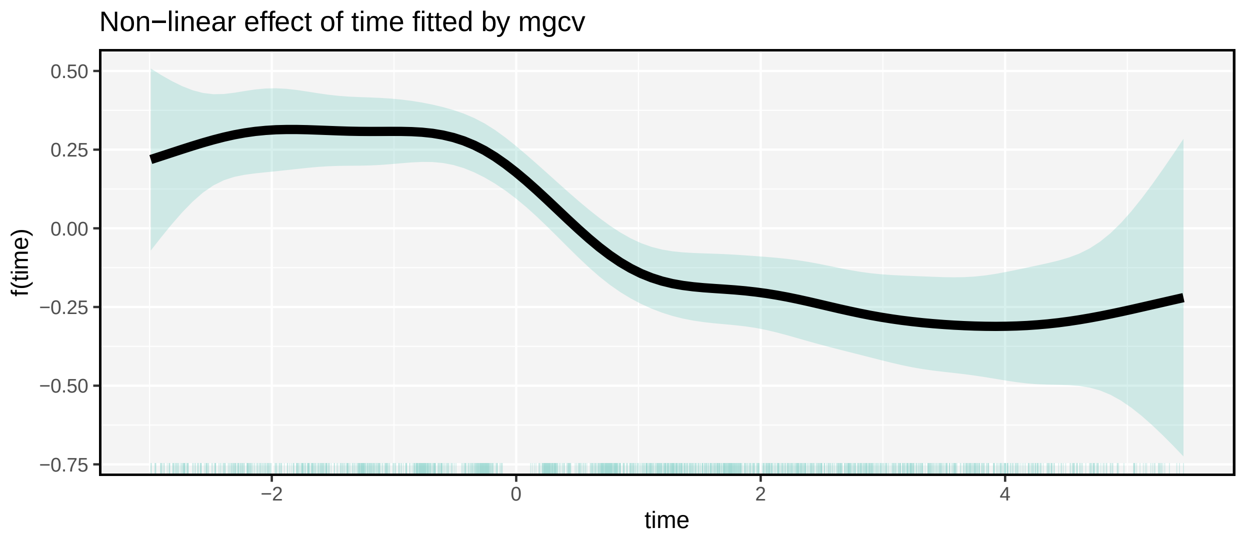

According the predicted patten of time effect, we wonder if $f(\text{time})$ is a combination of the linear segmented function and the linear discontinuous function:

$$f(\text{time}) = \alpha_1 \mathrm{I}(\text{time} > 0) + \alpha_2 \text{time} + \alpha_3 \{\mathrm{I}(\text{time} > 0)\text{time}\} + \alpha_4 (\text{time} - 2)_+,$$

which has a breakpoint at $\nu_0 = 0$ and a changepoint at $\nu_1 = 2$. The medical meaning of this structure is: with the increase of time, the counts of CD4 in patients will suddenly decrease at time $= 0$ (the changing trend of CD4 counts cannot increase); the slope will change at time $= 2$, indicating that the disease progress is more rapid or relatively slower:

```
fit1=gam(cd4~s(time,bs="AMatern",
     xt=list(getA=linearity_piecewise_discontinuity,para=list(para1=1.5,para=0)))
     +s(age,bs="gp")+s(cesd,bs="gp")+packs+drugs+sex, data=aids,
     family=quasipoisson(), method="REML")
summary(fit1)
```

```
##
## Family: quasipoisson
## Link function: log
##
## Formula:
## cd4 ~ s(time, bs = "AMatern", xt = list(getA = linearity_piecewise_discontinuity,
##     para = list(para1 = 1.5, para = 0))) + s(age, bs = "gp") +
##     s(cesd, bs = "gp") + packs + drugs + sex
##
## Parametric coefficients:
##              Estimate Std. Error t value Pr(>|t|)
## (Intercept)  6.482378   0.020299 319.350  < 2e-16 ***
## packs        0.074233   0.006149  12.073  < 2e-16 ***
## drugs        0.063477   0.022879   2.775  0.00557 **
## sex         -0.005452   0.002705  -2.015  0.04398 *
## ---
## Signif. codes:  0 '***' 0.001 '**' 0.01 '*' 0.05 '.' 0.1 ' ' 1
##
## Approximate significance of smooth terms:
##           edf Ref.df       F p-value
## s(time) 4.264  4.496 134.778  <2e-16 ***
## s(age)  1.001  1.003   1.002  0.3170
```

```
## s(cesd) 1.002  1.004   6.410  0.0113 *
## ---
## Signif. codes:  0 '***' 0.001 '**' 0.01 '*' 0.05 '.' 0.1 ' ' 1
##
## R-sq.(adj) =  0.274   Deviance explained = 28.1%
## -REML =   7110  Scale est. = 150.32    n = 2376
```

```
b1 = getViz(fit1)
plot(sm(b1, 1)) +
  l_ciPoly(mul = 5, fill = "#60c5ba", alpha = 0.25) +
  l_rug(mapping = aes(x=x), alpha = 0.25, color="#60c5ba") +
  l_fitLine(colour = "black", size=2) +
  theme_get() +
  xlab("time") + ylab("f(time)") +
  ggtitle("Mixed effect of time fitted by mgcv.taps with one breakpoint
          and one changepoint") +
  geom_vline(xintercept=0, linetype=2, size=0.75, color="grey30") +
  geom_vline(xintercept=1.5, linetype=2, size=0.75, color="grey30") +
  theme(panel.border = element_rect(colour = "black", fill=NA, size=1),
        panel.background = element_rect(fill = "#f4f4f4"))
```

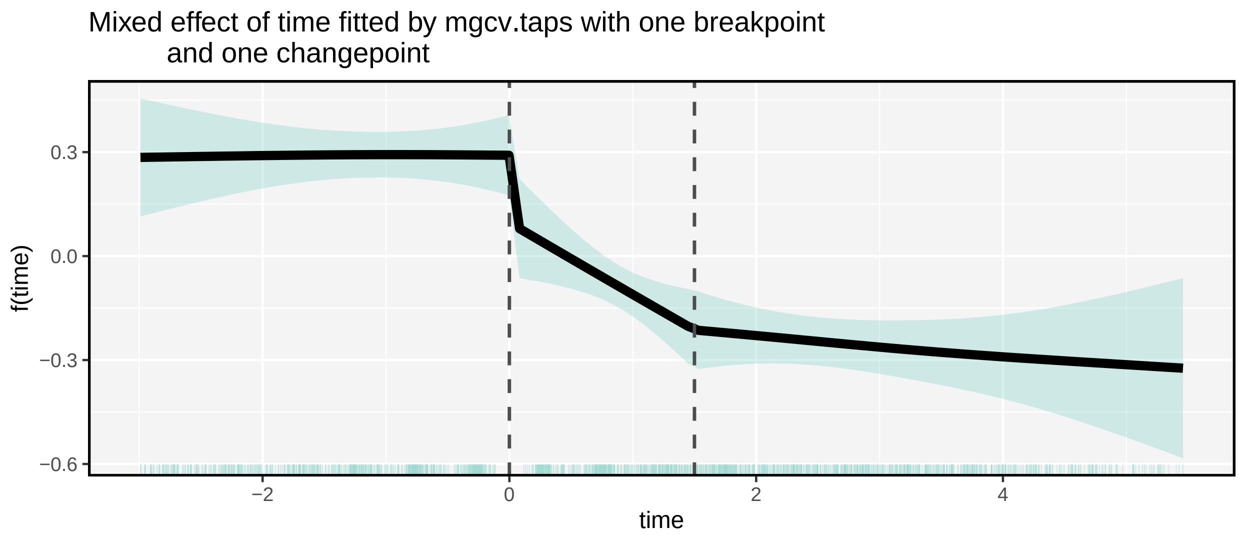

```
taps_score_test(fit1)
```

```
##    smooth.term smooth.pvalue method
##         <char>         <num> <char>
## 1:     s(time)     0.3524746 davies
```

```
# score test supports quasi exponential family of distributions
taps_wald_test(fit1)
```

```
##    mixed.term fix.df fix.chisq   fix.pvalue fix.indices smooth.df smooth.chisq
##        <char>  <num>     <num>        <num>      <char>     <num>        <num>
## 1:    s(time)      4  605.8852 8.25077e-130    reported 0.4960755    0.1112696
##    smooth.pvalue
##            <num>
## 1:     0.7387334
```

According to the result, $f(\text{time})$ is quite likely to follow the assigned structure. It shows that an HIV-infected man's CD4 cell counts remain unchanged at time=0, drop suddenly at time=0, continue to fall rapidly when $0 \leq$time$< 2$, and fall at a slower rate when time$\geq 2$. In addition, the $p$-values obtained by the score test and Wald test are 0.3652 and 0.7387, respectively, confirming our hypothesis.

In this example, we define a more complex case: the function $f$(time) involves both changepoints and breakpoints. To test this structure, we implement a custom parametric basis function `linearity_piecewise_discontinuity`, which is defined as follows:

```
linearity_piecewise_discontinuity <- function(x, para) {
changepoints <- para[[1]]
breakpoints <- para[[2]]
n <- length(x)
p1 <- length(changepoints)
p2 <- length(breakpoints)
B <- matrix(0, n, 2 + p1 + 2 * p2)
B[,1] <- 1  # Intercept
B[,2] <- x  # Linear term
if (p1 > 0) {
for (i in 1:p1) {
B[, 2 + i] <- pmax(x - changepoints[i], 0)
}
}
if (p2 > 0) {
for (i in 1:p2) {
B[, 2 + p1 + 2*i - 1]=as.numeric(x > breakpoints[i])
B[, 2 + p1 + 2*i]=(x - breakpoints[i]) * (x > breakpoints[i])
}
}
colnames(B) <- c("Intercept", "x",
                 paste0("Change_", changepoints),
                 as.vector(t(outer(breakpoints, c("_jump", "_slope"), paste0))))
return(B)
}
```

This example illustrates that when the parameter para becomes more complex (e.g., involving multiple elements such as changepoints and breakpoints), we require users to provide it as a list. Specifically, the xt argument in the s() function should always follow the format:

```
xt = list(getA = linearity_piecewise_discontinuity,
          para = list(changepoints = c(...), breakpoints = c(...)))
```

This ensures a consistent interface with exactly two named inputs: `getA` (the structure-generating function) and `para` (a list of parameters).

#### 4.3 mack data in R package gamair

The `mack` data in R package `gamair` relate to the distribution of mackerel eggs and were collected as part of the 1992 mackerel survey aimed at assessing the mackerel spawning stock biomass using the daily egg production method.

We use this data to introduce the usage of `"A2Matern"` that specifies a mixed-effects representation of a bivariate function. We first explore the data using `mgcv`:

```
library(gamair)
data("mack")
head(mack)
```

```
##   egg.count egg.dens b.depth   lat   lon  time salinity flow s.depth temp.surf
```

```
## 1         0     0.00    4342 44.57 -4.65  8.23    35.72  417     104       15.0
## 2         0     0.00    4334 44.57 -4.48  9.68    35.74  405      98       15.4
## 3         0     0.00    4286 44.57 -4.30 10.90    35.74  377     101       15.9
## 4         1    18.26    1438 44.02 -2.87 19.33    35.68  420      98       16.6
## 5         4    90.16     166 44.02 -2.07  8.78       NA  354     101       16.7
## 6         3    62.93     460 44.02 -2.13  9.42    35.17  373     100       16.6
##   temp.20m net.area country vessel vessel.haul     c.dist
## 1     15.0    0.242      SP   COSA          22 0.83951412
## 2     15.4    0.242      SP   COSA          23 0.85919263
## 3     15.9    0.242      SP   COSA          24 0.89301529
## 4     16.6    0.242      SP   COSA          93 0.39564087
## 5     16.7    0.242      SP   COSA         178 0.04000882
## 6     16.6    0.242      SP   COSA         179 0.09742345
```

```
fit0=gam(egg.count~s(lat,lon,bs="gp",k=30)+s(log(b.depth),bs="cr")+
           s(time,bs="cr")+s(log(flow),bs="cr")+s(temp.surf,bs="cr")+net.area,
           data=mack,method="REML",family=quasipoisson())
summary(fit0)
```

```
##
## Family: quasipoisson
## Link function: log
##
## Formula:
## egg.count ~ s(lat, lon, bs = "gp", k = 30) + s(log(b.depth),
##     bs = "cr") + s(time, bs = "cr") + s(log(flow), bs = "cr") +
##     s(temp.surf, bs = "cr") + net.area
##
## Parametric coefficients:
##             Estimate Std. Error t value Pr(>|t|)
## (Intercept)   0.5681     0.7951   0.715    0.475
## net.area      2.5499     3.6834   0.692    0.489
##
## Approximate significance of smooth terms:
##                    edf Ref.df      F p-value
## s(lat,lon)      21.276 23.701  7.322 < 2e-16 ***
## s(log(b.depth))  4.473  5.536 11.339 < 2e-16 ***
## s(time)          3.002  3.763  4.126 0.00389 **
## s(log(flow))     3.330  4.014 26.858 < 2e-16 ***
## s(temp.surf)     1.005  1.010  5.187 0.02367 *
## ---
## Signif. codes:  0 '***' 0.001 '**' 0.01 '*' 0.05 '.' 0.1 ' ' 1
##
## R-sq.(adj) =  0.721   Deviance explained = 81.6%
## -REML = 915.59  Scale est. = 6.5724    n = 634
```

```
fit_viz0 <- getViz(fit0)
plot(sm(fit_viz0, 1)) +
  l_fitRaster() +
  l_fitContour() +
  l_rug() +
  theme_classic() +
  labs(title = "Smooth surface of s(Latitude, Longitude)",
       x = "Latitude", y = "Longitude")
```

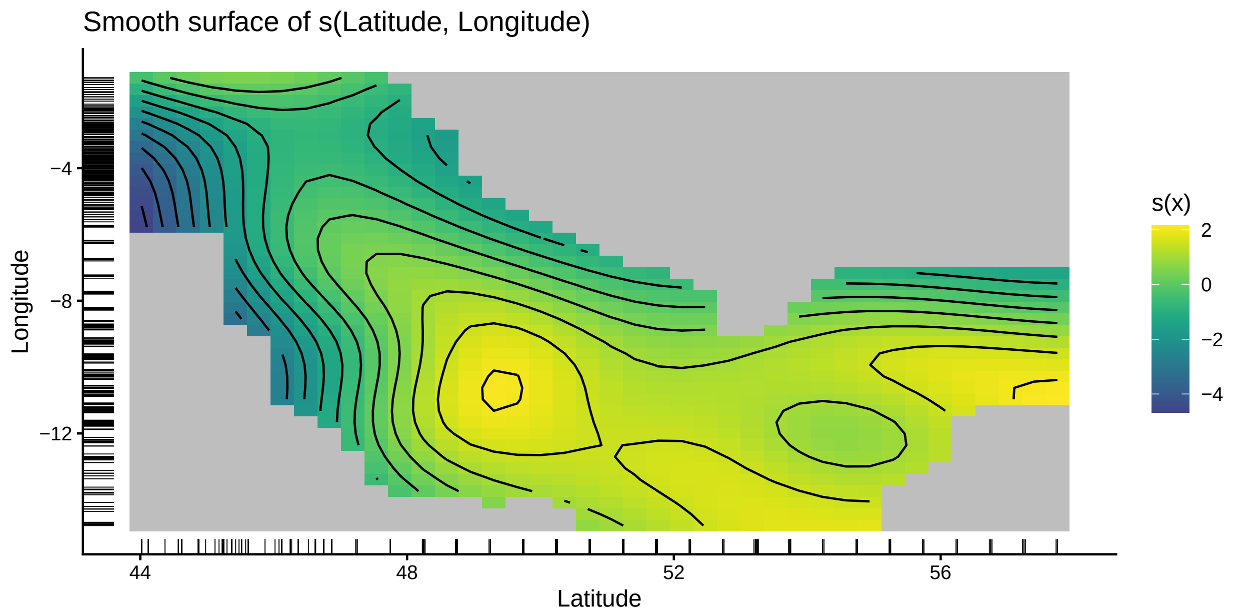

It is evident that the smooth surface s(Latitude, Longitude) is highly complex and cannot be adequately represented by a simple linear interaction. Nevertheless, we use this dataset as an illustrative example to demonstrate the usage of `mgcv.taps`:

```
fit1=gam(egg.count~s(lat,lon,bs="A2Matern",m=c(300,10), k=30,
    xt=list(getA=function(x1,x2,para){cbind(1,x1,x2,x1*x2)},para=NULL))+
           s(log(b.depth),bs="cr")+
           s(time,bs="cr")+s(log(flow),bs="cr")+s(temp.surf,bs="cr")+net.area,
           data=mack,method="REML",family=quasipoisson())
summary(fit1)
```

```
##
## Family: quasipoisson
## Link function: log
##
## Formula:
## egg.count ~ s(lat, lon, bs = "A2Matern", m = c(300, 10), k = 30,
##     xt = list(getA = function(x1, x2, para) {
##         cbind(1, x1, x2, x1 * x2)
##     }, para = NULL)) + s(log(b.depth), bs = "cr") + s(time, bs = "cr") +
##     s(log(flow), bs = "cr") + s(temp.surf, bs = "cr") + net.area
##
## Parametric coefficients:
##             Estimate Std. Error t value Pr(>|t|)
## (Intercept)   0.6792     0.8098   0.839    0.402
## net.area      2.0028     3.7537   0.534    0.594
##
## Approximate significance of smooth terms:
##                    edf Ref.df      F p-value
## s(lat,lon)      22.320 25.054  7.200  <2e-16 ***
## s(log(b.depth))  4.428  5.484 11.120  <2e-16 ***
## s(time)          2.982  3.739  3.870  0.0061 **
## s(log(flow))     3.399  4.087 27.442  <2e-16 ***
## s(temp.surf)     1.005  1.009  4.558  0.0338 *
```

```
## ---
## Signif. codes:  0 '***' 0.001 '**' 0.01 '*' 0.05 '.' 0.1 ' ' 1
##
## R-sq.(adj) =  0.725   Deviance explained = 81.9%
## -REML =  908.6  Scale est. = 6.5063    n = 634
```

```
fit_viz1 <- getViz(fit1)
plot(sm(fit_viz1, 1)) +
  l_fitRaster() +
  l_fitContour() +
  l_rug() +
  theme_classic() +
  labs(title = "Smooth surface of s(Latitude, Longitude) with parametric structure
       being linear interaction",
       x = "Latitude", y = "Longitude")
```

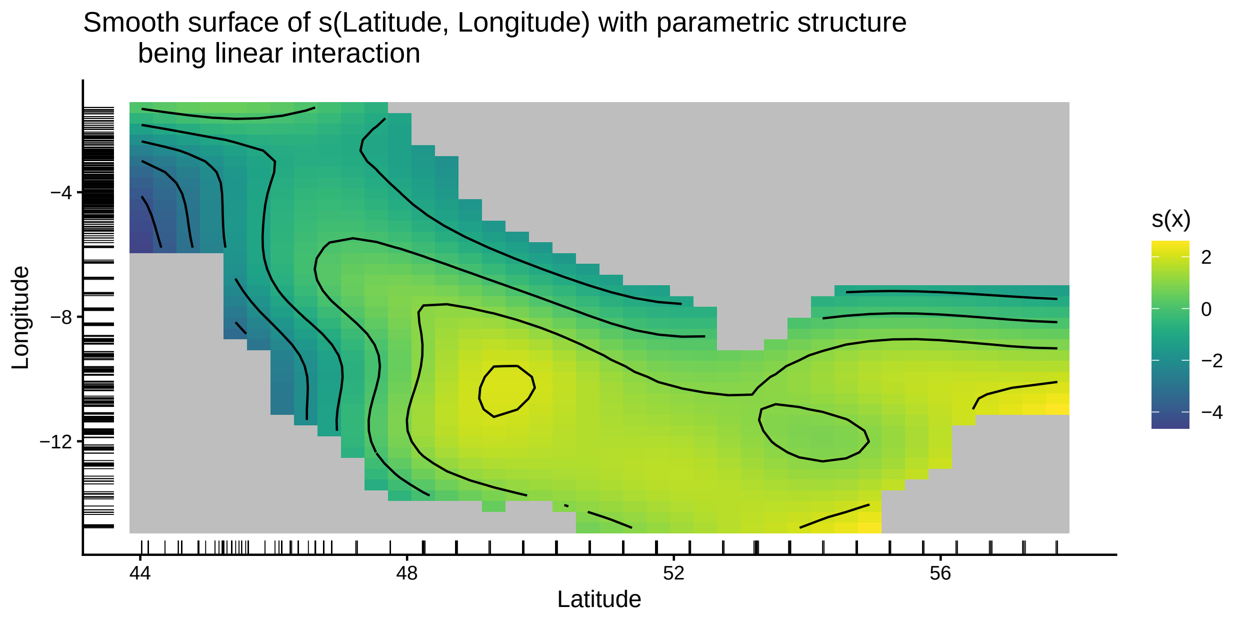

```
taps_score_test(fit1)
```

```
##    smooth.term smooth.pvalue method
##         <char>         <num> <char>
## 1:  s(lat,lon)    0.00375281 davies
```

```
taps_wald_test(fit1)
```

```
##    mixed.term fix.df fix.chisq   fix.pvalue fix.indices smooth.df smooth.chisq
##        <char>  <num>     <num>        <num>      <char>     <num>        <num>
## 1: s(lat,lon)      3  39.73968 1.209852e-08    reported  22.05409      103.935
##    smooth.pvalue
##            <num>
## 1:             0
```

The "A2Matern" basis is used for bivariate smooths. Compared to the univariate case, the only difference is that the getA function must explicitly accept x1, x2, and para as arguments. When the structure of para is complex, it must be provided as a named list.

[10] David Ruppert, Matt P Wand, and Raymond J Carroll. Semiparametric Regression. Cambridge University Press, 2003.

[11] Grace Wahba. Spline Models for Observational Data. SIAM, 1990.

[12] Grace Wahba and James Wendelberger. "Some new mathematical methods for variational objective analysis using splines and cross validation". In: Monthly Weather Review 108.8 (1980), pp. 1122–1143.

[13] Martin J Wainwright. High-Dimensional Statistics: A Non-Asymptotic Viewpoint. Cambridge University Press, 2019.

[14] Simon N Wood. Generalized Additive Models: An Introduction with R. CRC Press, 2017.
